# Factors associated with hospitalization of children aged 6-35 months presenting with diarrhea in six low- and middle-income countries: a cohort analysis in the Enterics for Global Health (EFGH) study, 2022 – 2024

**DOI:** 10.64898/2025.12.18.25342637

**Authors:** Mahzabeen Ireen, Sonia I. Rao, Beth A Tippett Barr, Brian O. Onyando, Billy Ogwel, Donnie Mategula, Faisal Ahmmed, Firdausi Qadri, Farah Naz Qamar, Hannah E. Atlas, Md Nazmul Hasan Rajib, Md Ismail Hossen, Musa Jallow, Maureen Ndalama, M. Jahangir Hossain, Patricia B Pavlinac, Pablo Penataro Yori, Paul Garcia Bardales, Saba Siddiqui, Farhana Khanam, Md Taufiqul Islam

## Abstract

**Background:** Around 1.7 billion cases of diarrhea are reported globally, with the highest burden in Africa and South-East Asia, where higher hospitalization rates are observed among children under five years of age. Hospitalized children with diarrhea often experience serious complications and a higher risk of death. Although various known risk factors contribute to these hospitalizations, they may vary by country. The purpose of this analysis was to identify host, clinical, and sociodemographic factors associated with hospitalization among children presenting with diarrhea in the Enterics for Global Health (EFGH) *Shigella* surveillance study, conducted from June 2022 to October 2024.

**Methodology:** We conducted a secondary data analysis of longitudinal cohorts from six EFGH study sites. Hospitalization was defined as the child being admitted from at least 12 am to 6 am or self-discharged before that timeframe. Information on risk factors, clinical episodes, and hospital management was collected. We used descriptive statistics to summarize frequencies and proportions, and robust Poisson regression to estimate risk ratios (RRs) for hospitalization, adjusting for age and study site, to examine the associations with risk factors.

**Results:** Overall, 5% of children experienced diarrhea-related hospitalizations at enrollment or during the three months of follow-up. Several risk factors were significantly associated with higher likelihood of diarrhea-related hospitalization, including vomiting (adjusted RR [aRR], 2.53; 95% CI, 1.98- 3.22; p<0.001), severe dehydration (aRR, 17.40; 95% CI, 12.74-23.78; p<0.001), severe wasting (aRR, 2.62; 95% CI, 1.78-3.87; p<0.001) and severe diarrhea (aRR, 4.79; 95% CI, 3.69-6.20; p<0.001). Older age was associated with a lower risk of hospitalization, with children 24-35 months experiencing 57% lower risk compared to 6-11 months (aRR 0.43; 95% CI, 0.29-0.64); p<0.001).

**Conclusion:** These findings highlight the need for integrated approaches to reduce hospitalization rates for diarrhea in young children and to develop policies and interventions to mitigate the impact of childhood diarrhea.

## Introduction

In 2019 Global Burden of Disease Study identified childhood diarrheal diseases as a continuing and significant global health challenge [1]. Each year, around 1.7 billion cases of diarrhea are reported globally, making it the third leading cause of death among children aged 1-59 months [2]. In 2021, diarrhea accounted for 9% of all deaths among children under five, with an estimated 1,200 deaths globally per day [3]. The highest disease burden was reported from Africa and South-East Asia [4–6].

The substantial global burden underscores the urgent need for targeted interventions, especially in regions with the highest incidence of childhood diarrheal disease. This health challenge is particularly significant in low- and middle-income countries (LMICs), where young children face disproportionately higher hospitalization rates due to diarrhea, compared to those in high-income countries [7, 8]. For example, in Bangladesh, the hospitalization rate was 9.6 per 1,000 person-years from 2011 to 2013 [9], and in Kenya, diarrhea accounted for 11.2% of hospitalizations for all age groups, among which 16.9% were seen in children aged <5 years between 2001 and 2003 [10].

Hospitalized children with diarrhea cause severe complications and increased morbidity and mortality, including dehydration, malnutrition, and potentially life-threatening conditions like sepsis, especially if bacteremia is present, or if they have pre-existing conditions like severe stunting or wasting [11–13]. This situation also worsens the economic hardship for affected families [14–16]. The Global Enteric Multicenter study (GEMS) found that out-of-pocket costs per episode of diarrhea range from $14.88 in Kenya and $17.87 in Mali [17]. These costs tend to be higher for more severe cases of diarrhea and younger children [18].

Several known risk factors contribute to diarrhea-related hospitalizations, including early age, delayed care, malnutrition, dehydration, lack of exclusive breastfeeding, low educational level, inadequate hygiene, and immune deficiency [18–26]. While previous studies have identified risk factors for diarrheal hospitalizations in children, there is a lack of evidence on regional variations and age-specific vulnerabilities. Children aged 6–35 months experienced unique biological and environmental risks that may differ across settings, though few studies have systematically examined these factors. To address this gap, we conducted an analysis to identify host, clinical, and sociodemographic factors associated with hospitalization among children aged 6-35 months presenting with diarrhea at enrollment, or within three months of follow-up, in the Enterics for Global Health (EFGH) health-facility based *Shigella* surveillance study.

## Materials and methods

### Study design, area, and population

We conducted a secondary analysis using data from the EFGH *Shigella* surveillance study. The study design was a cross-sectional and longitudinal cohort started in June 2022 in seven countries, including Bangladesh, Kenya, Malawi, Mali, Pakistan, Peru, and The Gambia [27]. Facility-based surveillance was conducted for 24 months to estimate the incidence rates and consequences of *Shigella-*attributable diarrhea among children aged 6-35 months presenting with diarrhea who were enrolled in the study. Each EFGH health facility had a dedicated study team that worked five or six days a week. Children presenting with diarrhea, defined as having three or more abnormally loose or watery stools in the past 24 hours, were screened using a standardized screening questionnaire after obtaining verbal consent from their caregivers.

To be eligible for the main study, children had to meet specific criteria, which included children whose primary caregiver provided informed written consent, planned to reside in the study area for the next four months, and were willing to have the child participate in follow-up visits. Enrollment was completed within four hours of the child’s presentation at the facility. Children were excluded if their diarrheal episode had lasted more than seven days at the time of enrollment, if they were referred to a non-EFGH facility for care, if their caregiver was unwilling or unable to provide consent or withdrew consent, or if the site-specific enrollment cap had been reached. Each enrolled child was identified with a specific participant identification number. At enrollment, a clinical examination, anthropometry measurement, medical record abstraction, and caregiver interviews were carried out. Socio-demographics, water, sanitation, and hygiene (WASH) behaviour, breastfeeding, vaccination history, and other related information were collected during the interview. Rectal swab, stool, and dried blood spot samples were collected. At week 4 and month 3 follow-ups, data were collected on the child’s condition, health history since the last visit, recovery status from the index diarrheal episode, new diarrheal episodes, associated care-seeking behavior, recent hospitalization, and other illnesses. Information on the updated anthropometric measurements was also collected. The full study protocol and detailed enrollment procedures were published elsewhere [27–34].

### Definitions used in the analysis

**<u>Diarrhea</u>** was defined as the passage of three or more abnormally loose or watery stools in the past 24 hours **Index diarrheal episode** was defined as the episode that prompted a child’s enrollment in EFGH.

**<u>Hospitalization</u>** was defined as hospitalization during the index diarrhea episode or within 3 months of follow-up after the index diarrheal episode had been resolved. Only new hospitalizations that were not a continuation of management from a previous hospitalization were included. To standardize across sites where intake and discharge processes may vary, hospitalization was defined as an overnight stay, i.e., the child was in the ward from at least 12 am to 6 am or self-discharged prior to that timeframe.

**<u>Exclusive breastfeeding</u>** was defined by the age in months at which the child first consumed non-breastmilk fluids or foods. The child is categorized as exclusively breastfed if this age is greater than 6 months.

**Receipt of age-appropriate vaccines** was defined by whether the child had received all age-appropriate vaccines (BCG, polio, DPT, hepatitis B, Hib, rotavirus, MMR, pneumococcal pneumonia) according to each country’snational immunization schedule, allowing for a 1-month delay.

**Blood in stool** was defined by the caregiver’s report of blood in stool during the child’s index diarrhea episode or a clinician’s diagnosis during the enrollment procedures.

**Vomiting** was defined as whether the child had any vomiting during the index diarrhea episode (vomiting duration greater than 0 days or maximum number of vomits per day greater than 0).

**Fever** was defined as whether the child had any fever during the index diarrhea episode, with fever duration greater than 0 days or maximum axillary temperature greater than or equal to 37.2°C.

**<u>Dehydration</u>** was defined using the WHO dehydration categories as per the Integrated Management of Childhood Illness (IMCI) guidelines [2]. Categorized as “none”, “some”, or “severe”, calculated from the clinician’s physical assessment of the child at enrollment.

**Acute comorbidity** was defined as when the child had any of the following diagnoses at enrollment: anemia, urinary tract infection, malaria, suspected sepsis, fever of unknown origin/febrile illness, upper respiratory tract infection, pneumonia, other lower respiratory tract infections, meningitis, acutely unwell - unknown cause, poisoning/herbal intoxication, asthma, or soft tissue/skin infection.

**Chronic comorbidity** was defined as when the child had any of the following at enrollment: HIV, sickle cell anemia, tuberculosis, or caregiver reports the child had ever been diagnosed with any of the following: rheumatic heart disease, leukemia, sickle cell anemia, tuberculosis, HIV, or other chronic condition.

**<u>Diarrhea severity</u>** was based on the Modified Vesikari diarrhea severity score (MVS). MVS was calculated as follows (Table 1):

**Table 1:** Modified Vesikari Clinical Severity scoring system parameters and scores [35, 36]

| Parameter | Score |  |  |
| --- | --- | --- | --- |
|  | 1 | 2 | 3 |
| Diarrhea |  |  |  |
| Maximum number of stools per day | 1-3 | 4-5 | ≥6 |
| Diarrhea duration (days) | 1-4 | 5 | ≥6 |
| Vomiting |  |  |  |
| Maximum number of vomiting episodes per day | 1 | 2-4 | ≥5 |
| Vomiting duration (days) | 1 | 2 | ≥3 |
| Temperature (axillary) | 36.6-37.9°C | 38.0-38.4°C | ≥38.5°C |
| Dehydration | N/A | 1-5% | ≥6% |
| Treatment | Rehydration | Hospitalization | N/A |
| **Severity Category** |  |  |  |
| Maximum score (20) | Mild (<7) | Moderate (7-10) | Severe (≥11) |

\*Table adapted from rotavirus clinical trials utilizing the Vesikari clinical severity scoring system (Clark et al., 2004; Ruuska & Vesikari, 1990).

**<u>Underweight</u>** was defined by weight-for-age z-score (WAZ) at enrollment. WAZ < -3 was classified as severe, -3 ≤ WAZ < -2 as moderate, and WAZ ≥ -2 as none.

**<u>Wasting</u>** was defined by weight-for-height (WHZ) and mid-upper arm circumference (MUAC) at enrollment. WHZ < -3 or MUAC <11.5 was classified as severe, -3 ≤ WHZ < -2 or 11.5 ≤ MUAC >12.5 as moderate, and WHZ ≥ -2 and MUAC≥12.5 as none.

**<u>Stunting</u>** was defined by length/height-for-age z-score (LAZ/HAZ) at enrollment. LAZ/HAZ < -2 was classified as stunted, and LAZ/HAZ ≥ -2 as not stunted.

**Time taken to seek care** was calculated based on the difference in days between when the caregiver reported the child’s diarrhea started and the date the child was enrolled.

**Improved household drinking water source** was defined as water piped into dwelling, piped into compound/yard/plot, piped to a neighbor’s house, public tap/standpipe, tube well or borehole, protected well, water from spring (protected), rainwater, tanker-truck, cart with small tank/drum, water kiosk (filtered), bottled water, or sachet water [37].

**Improved sanitation** was defined as flushed or pour flush to piped sewer system, flushed or pour flush to septic tank, ventilated improved pit latrine, pit latrine with slab floor, or composting toilet [38].

**<u>Wealth index quintile</u>** was defined as five categories of the wealth index that align with either urban or national wealth quintiles, depending on whether the caregiver and child’s catchment area was located in an urban or rural/mixed urban-rural setting. The first quintile indicates less wealth, and the fifth quintile indicates more wealth [39].

### Sample size

A total of 9,476 children were enrolled in the EFGH study at seven sites from June 2022 to August 2024, and follow-up of participants continued until December 2024, including a window period for 3 months follow-up. We excluded Mali from the analysis due to only one hospitalization case. We excluded children with missing data for predictor, covariate, or outcome variables, resulting in a final sample size of 6,052. (Fig 1).

**Fig 1:**
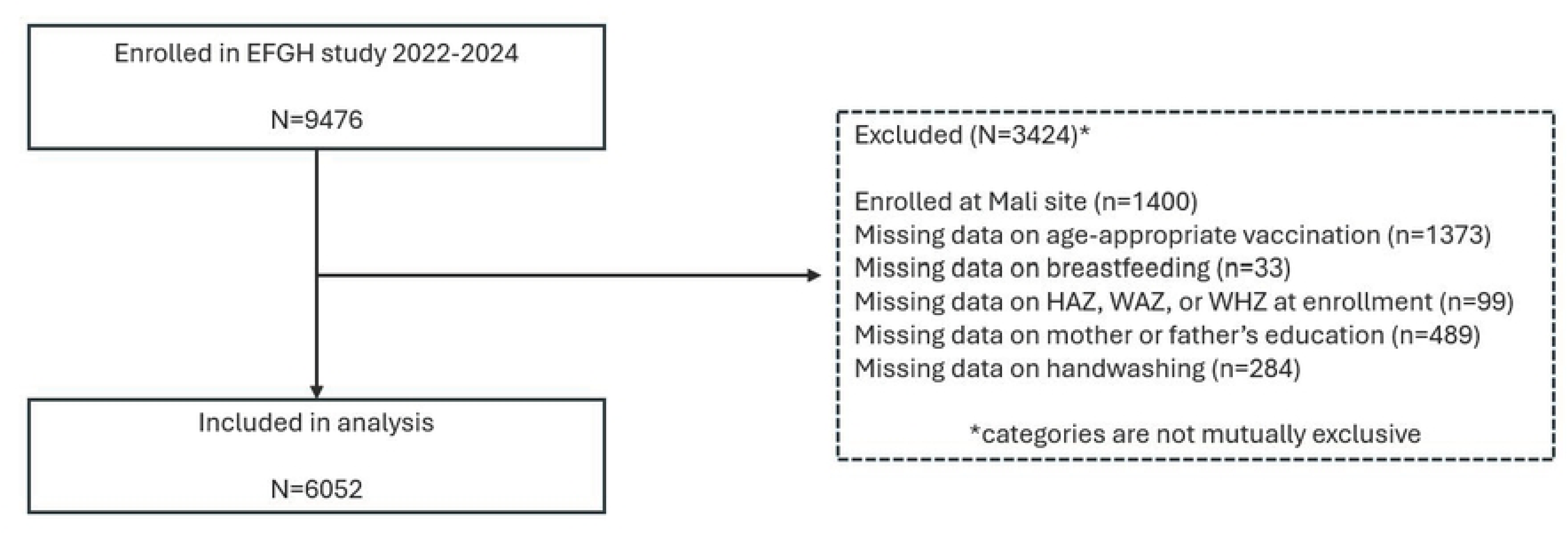
**Study participants included in final analysis**

### Statistical analysis

Descriptive statistics were presented using frequency and proportions. To describe the association between host, clinical, and sociodemographic risk factors and a binary outcome of hospitalization, crude and adjusted risk ratios (aRR) with 95% confidence intervals (CI) were estimated using bivariate Poisson regression with robust standard errors. We adjusted for age and study site. We used a generalized estimating equations model to account for children who were enrolled multiple times over the study period. For site-specific analyses, we ran chi-square or Fisher’s exact test to compare hospitalized and non-hospitalized groups. All analysis was conducted in R Studio (version 4.3.1).

### Ethical clearance

This secondary analysis did not require additional ethical clearance beyond the approvals of the parent study at each country site, details of which can be found elsewhere [27].

## Results

Across the six EFGH sites, 305 children (5%) were hospitalized out of 6,052 children, and Bangladesh had the highest proportion (8.7%), where Malawi (1.1%) reported the lowest proportion of hospitalization (Table 2).

**Table 2.**
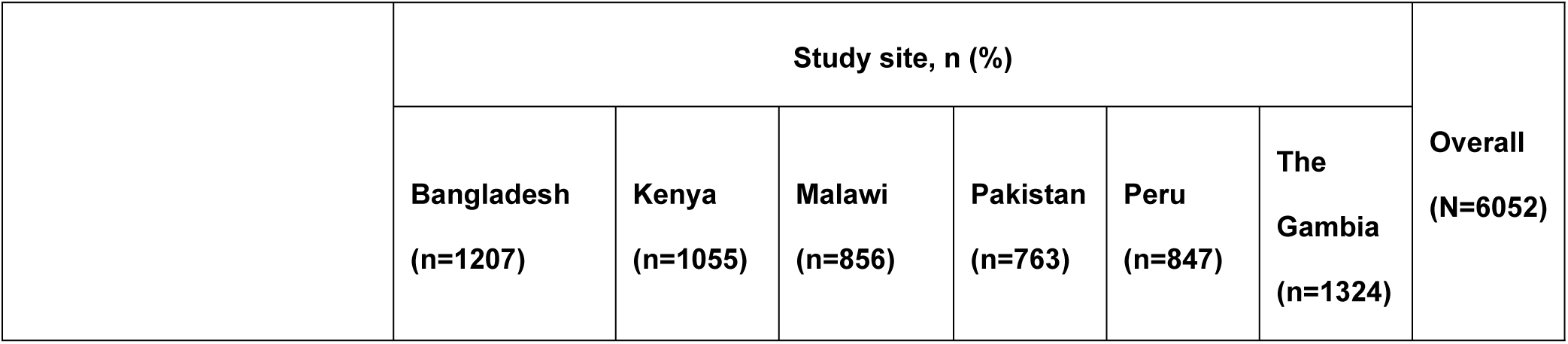
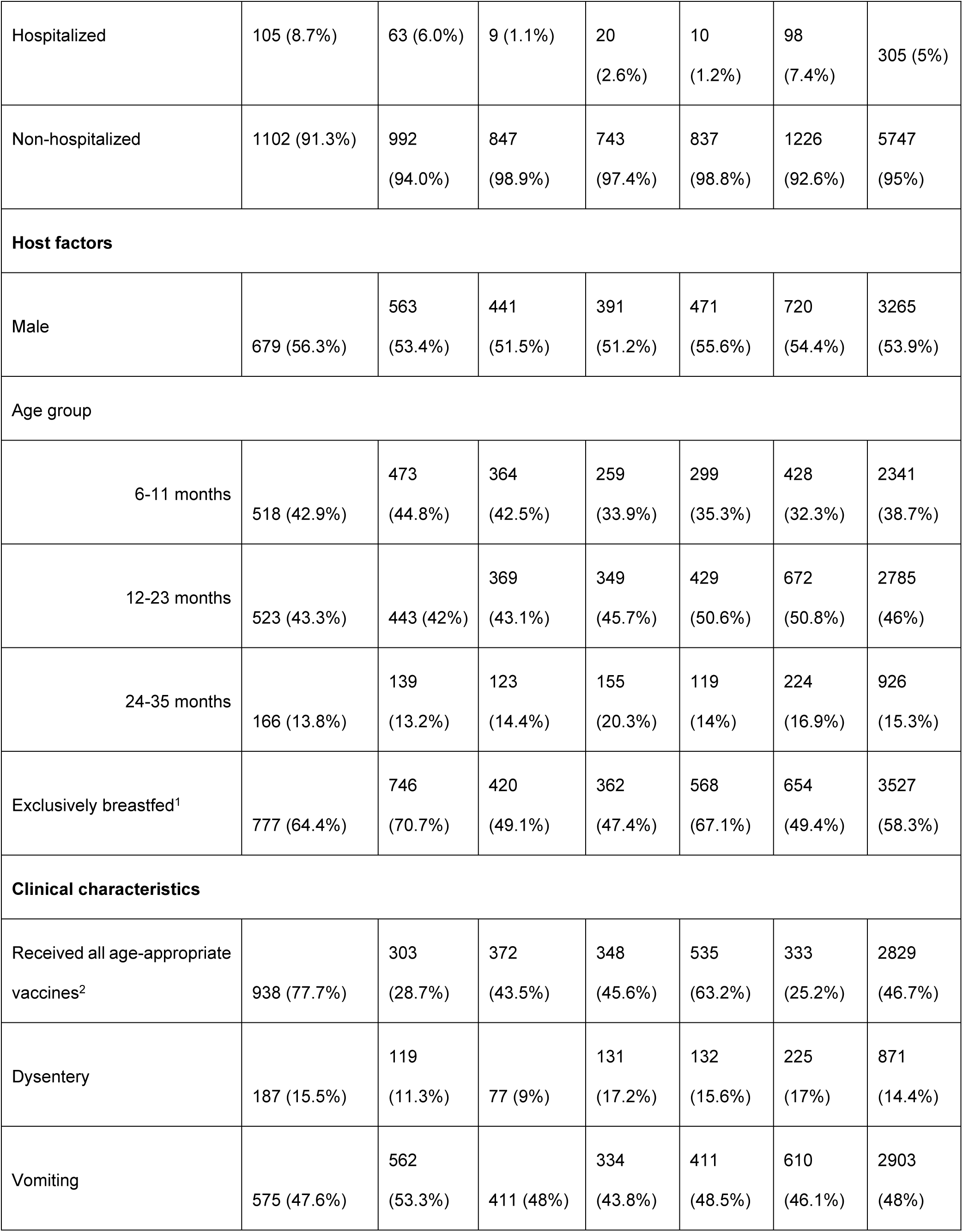
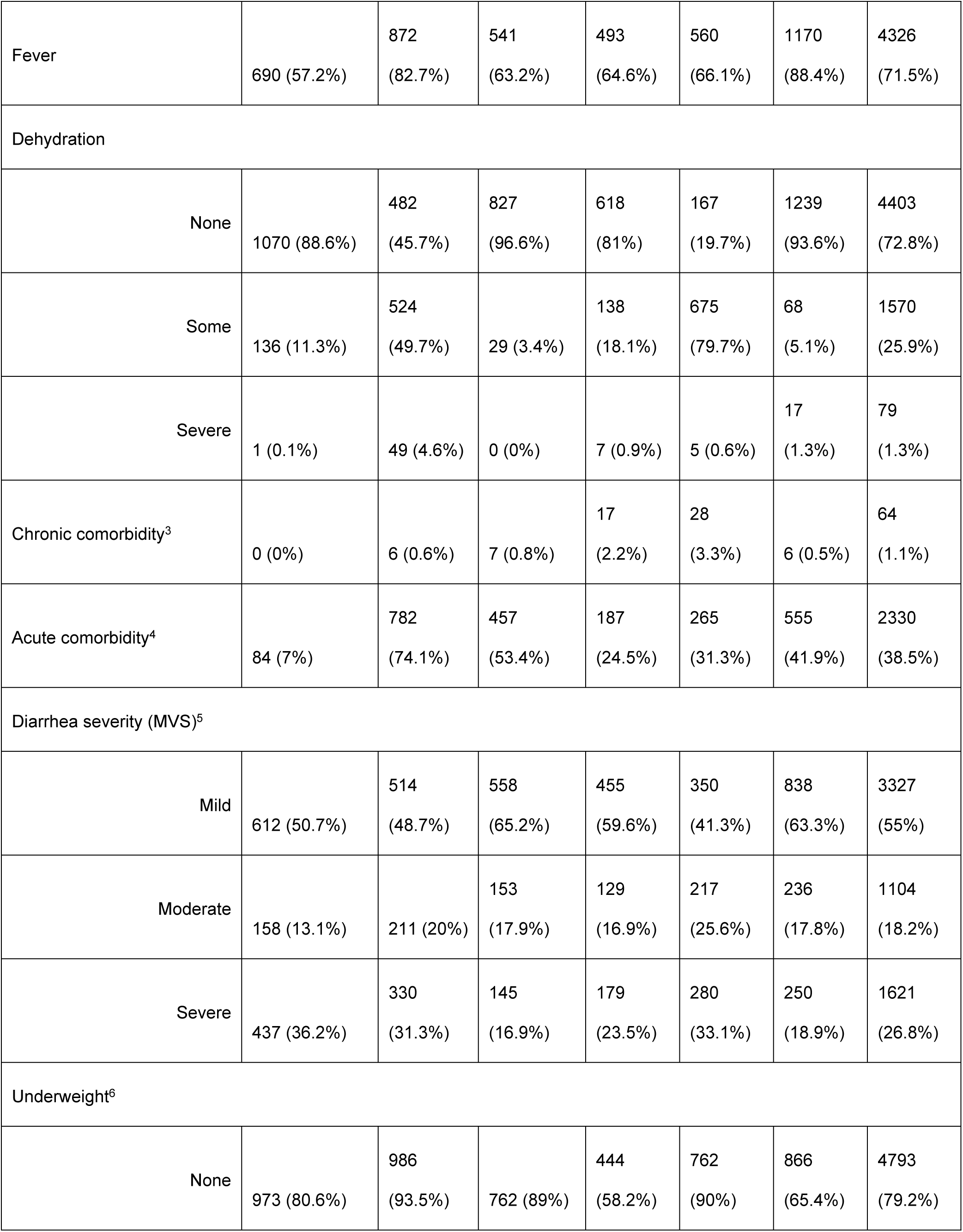
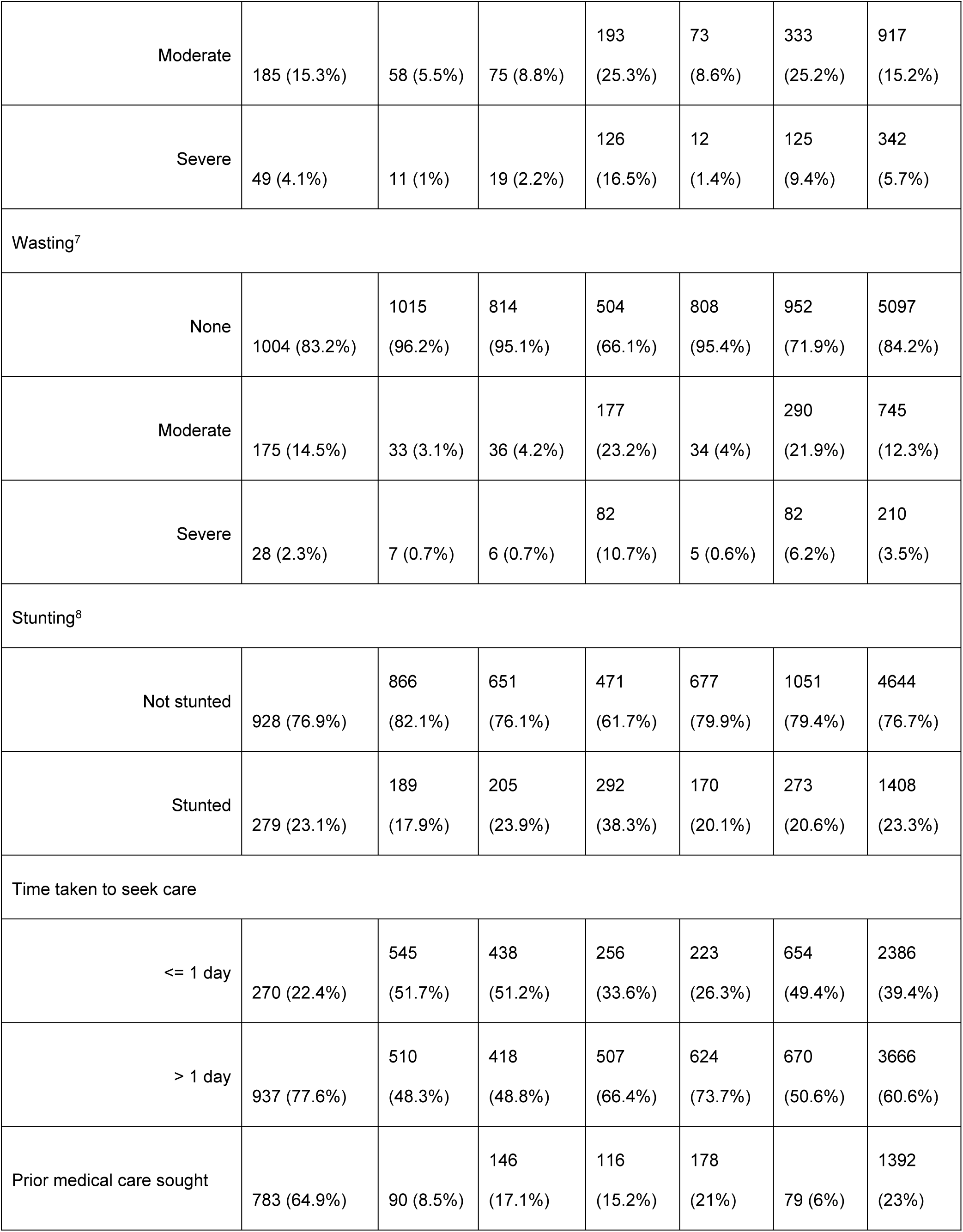
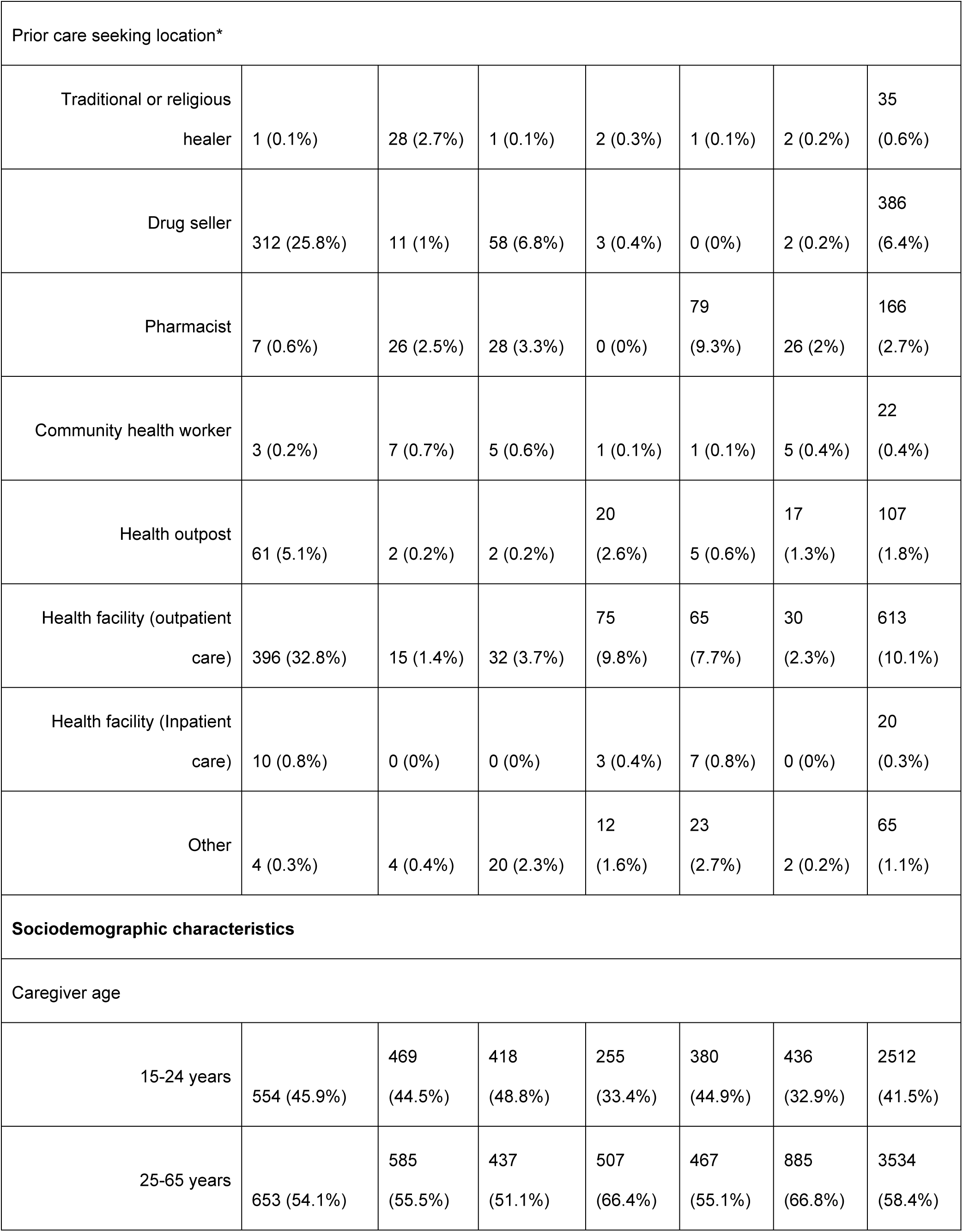
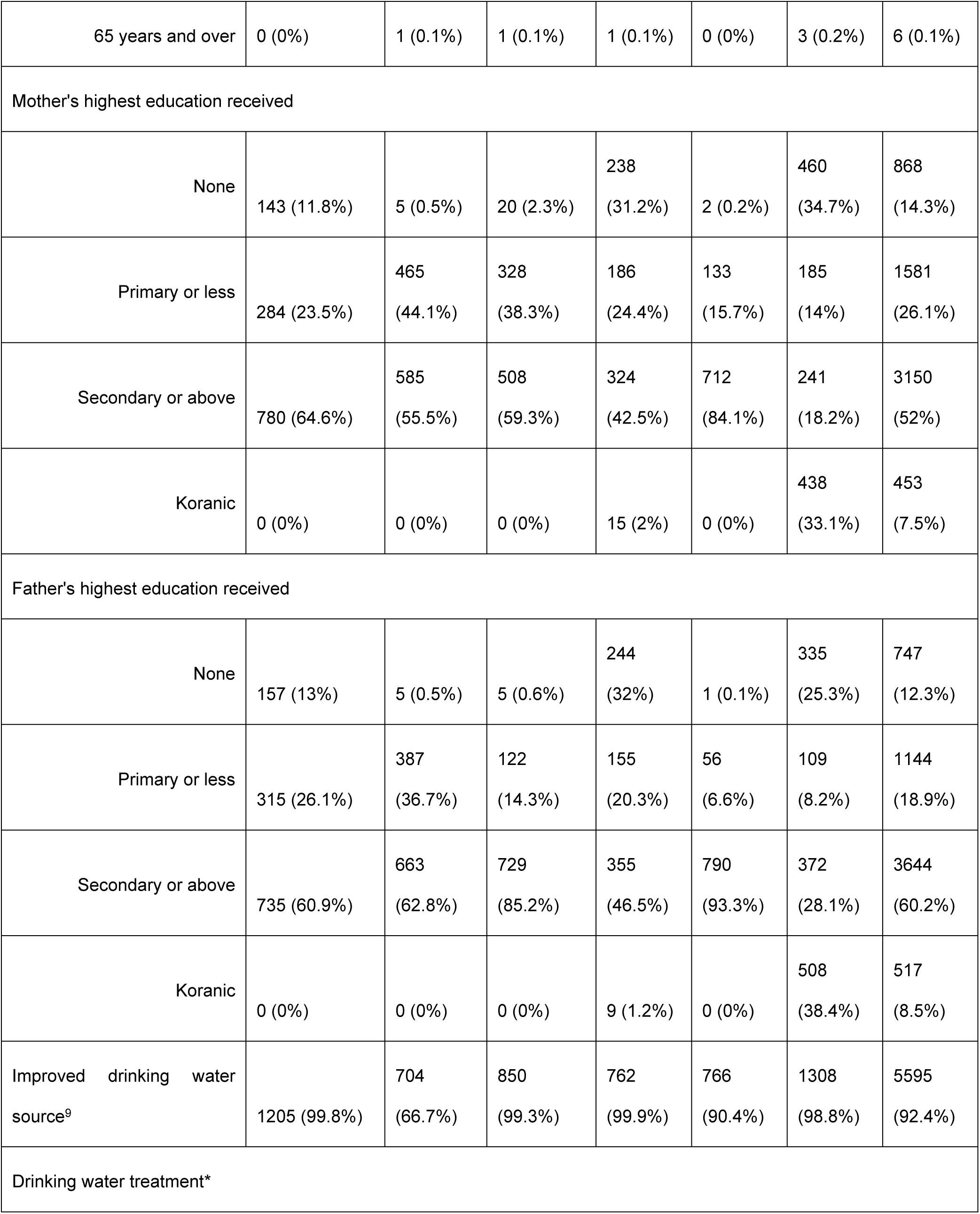
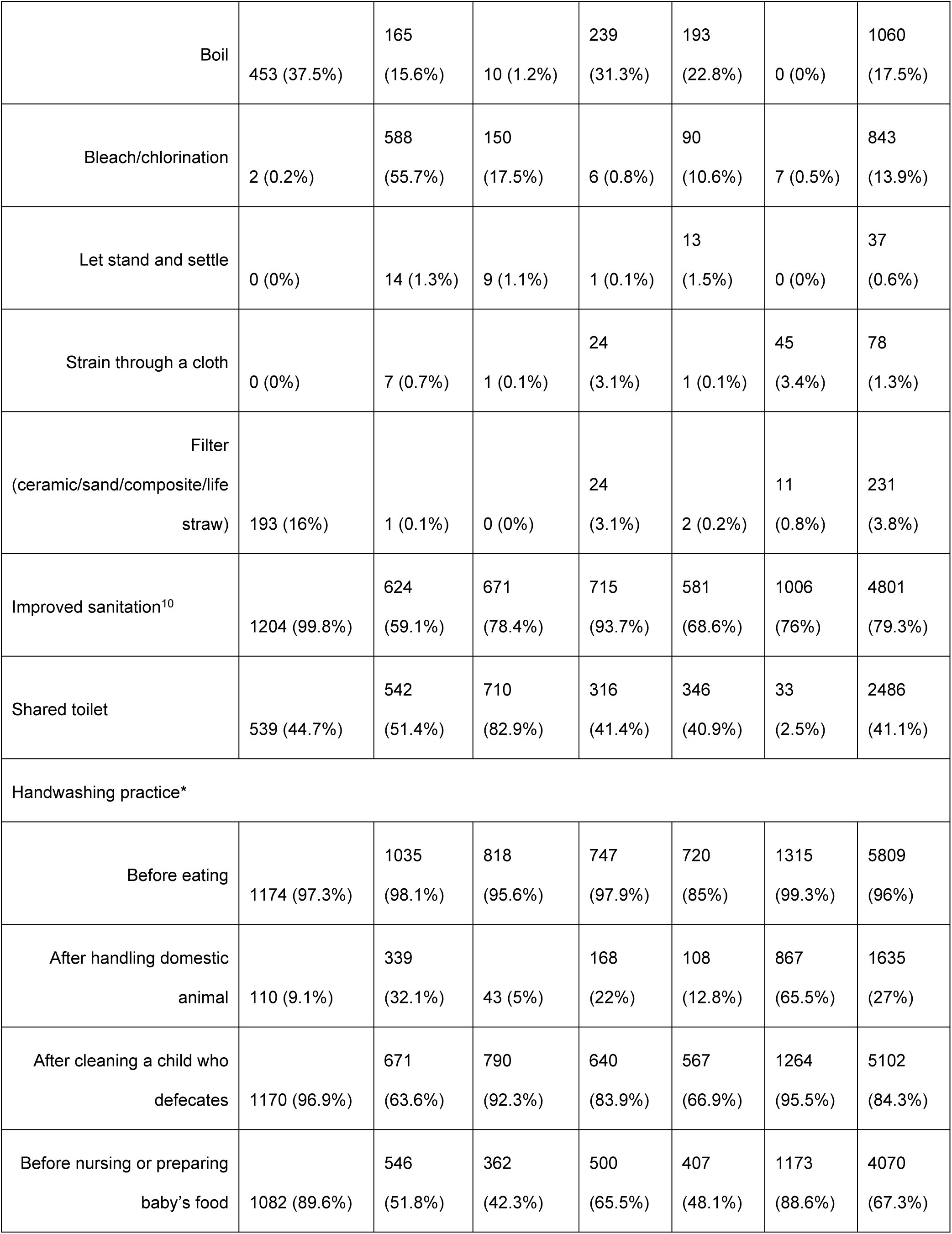
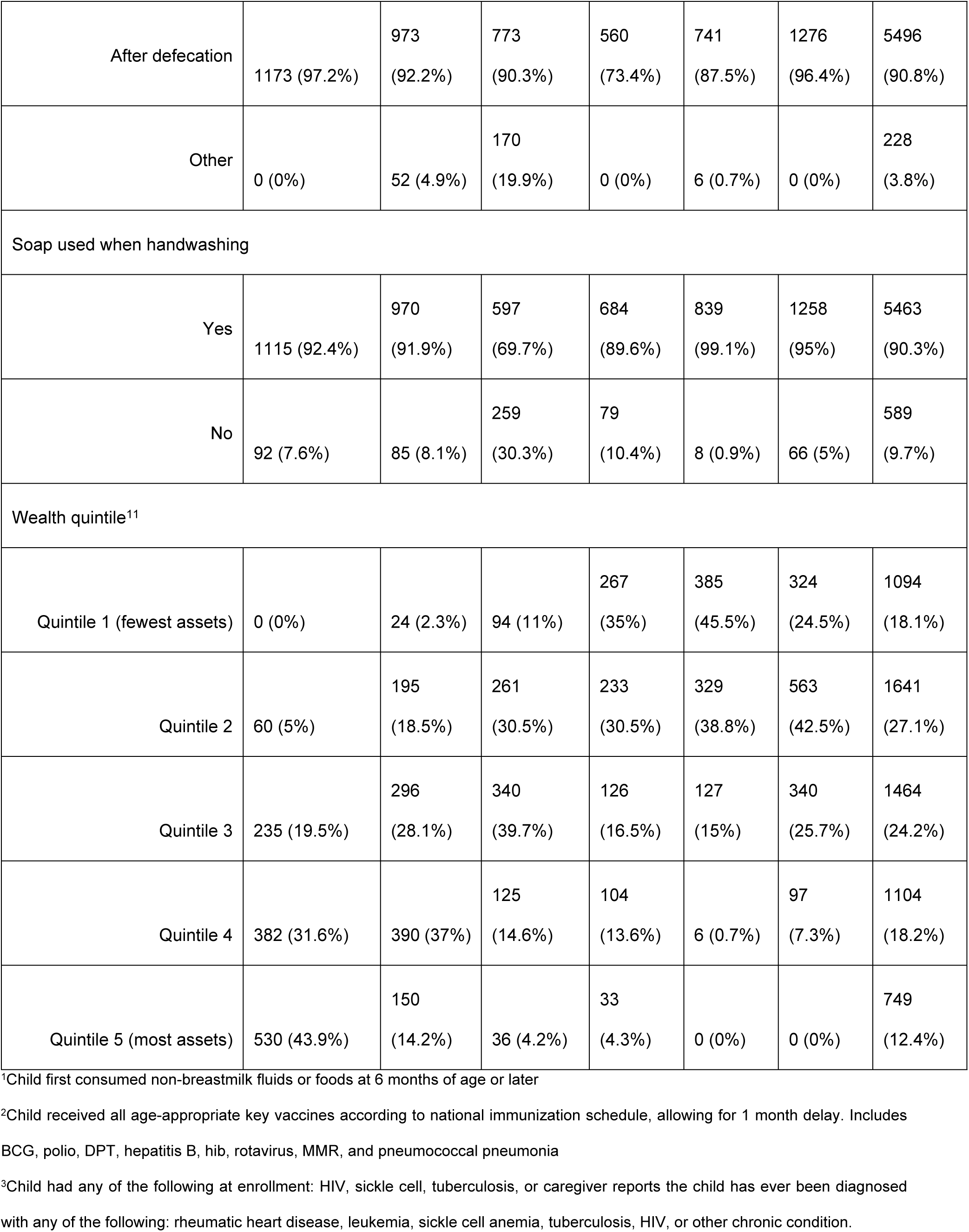
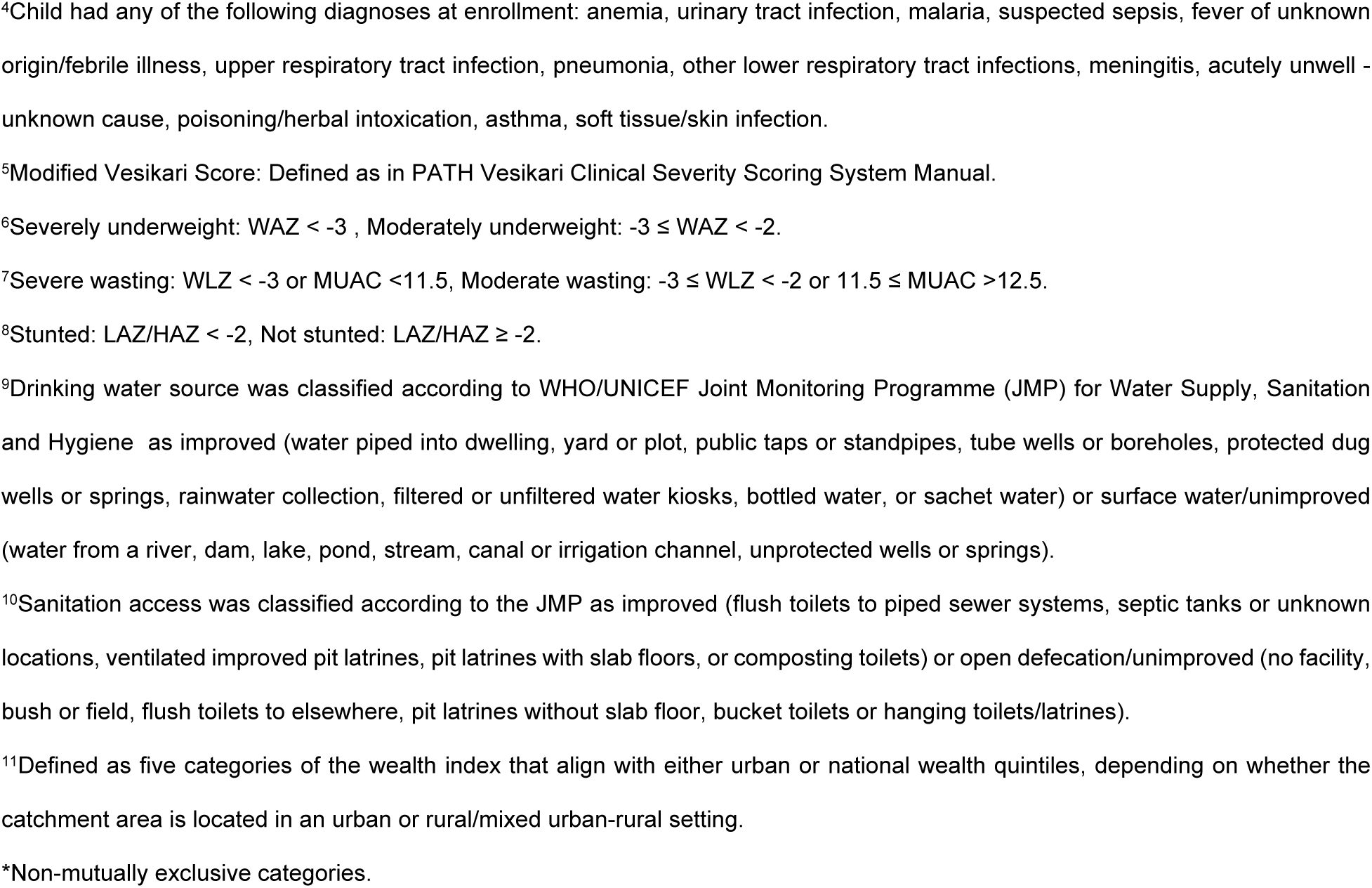
Characteristics of children aged 6-35 months enrolled in the EFGH study (N=6052)

|  | Study site, n (%) |  |  |  |  |  | Overall<br>(N=6052) |
| --- | --- | --- | --- | --- | --- | --- | --- |
|  | Bangladesh<br>(n=1207) | Kenya<br>(n=1055) | Malawi<br>(n=856) | Pakistan<br>(n=763) | Peru<br>(n=847) | The<br>Gambia<br>(n=1324) |  |
| Hospitalized | 105 (8.7%) | 63 (6.0%) | 9 (1.1%) | 20<br>(2.6%) | 10<br>(1.2%) | 98<br>(7.4%) | 305 (5%) |
| Non-hospitalized | 1102 (91.3%) | 992<br>(94.0%) | 847<br>(98.9%) | 743<br>(97.4%) | 837<br>(98.8%) | 1226<br>(92.6%) | 5747<br>(95%) |
| **Host factors** |  |  |  |  |  |  |  |
| Male | 679 (56.3%) | 563<br>(53.4%) | 441<br>(51.5%) | 391<br>(51.2%) | 471<br>(55.6%) | 720<br>(54.4%) | 3265<br>(53.9%) |
| **Age group** |  |  |  |  |  |  |  |
| 6-11 months | 518 (42.9%) | 473<br>(44.8%) | 364<br>(42.5%) | 259<br>(33.9%) | 299<br>(35.3%) | 428<br>(32.3%) | 2341<br>(38.7%) |
| 12-23 months | 523 (43.3%) | 443 (42%) | 369<br>(43.1%) | 349<br>(45.7%) | 429<br>(50.6%) | 672<br>(50.8%) | 2785<br>(46%) |
| 24-35 months | 166 (13.8%) | 139<br>(13.2%) | 123<br>(14.4%) | 155<br>(20.3%) | 119<br>(14%) | 224<br>(16.9%) | 926<br>(15.3%) |
| Exclusively breastfed<sup>1</sup> | 777 (64.4%) | 746<br>(70.7%) | 420<br>(49.1%) | 362<br>(47.4%) | 568<br>(67.1%) | 654<br>(49.4%) | 3527<br>(58.3%) |
| **Clinical characteristics** |  |  |  |  |  |  |  |
| Received all age-appropriate vaccines<sup>2</sup> | 938 (77.7%) | 303<br>(28.7%) | 372<br>(43.5%) | 348<br>(45.6%) | 535<br>(63.2%) | 333<br>(25.2%) | 2829<br>(46.7%) |
| Dysentery | 187 (15.5%) | 119<br>(11.3%) | 77 (9%) | 131<br>(17.2%) | 132<br>(15.6%) | 225<br>(17%) | 871<br>(14.4%) |
| Vomiting | 575 (47.6%) | 562<br>(53.3%) | 411 (48%) | 334<br>(43.8%) | 411<br>(48.5%) | 610<br>(46.1%) | 2903<br>(48%) |
| Fever | 690 (57.2%) | 872 (82.7%) | 541 (63.2%) | 493 (64.6%) | 560 (66.1%) | 1170 (88.4%) | 4326 (71.5%) |
| Dehydration |  |  |  |  |  |  |  |
| None | 1070 (88.6%) | 482 (45.7%) | 827 (96.6%) | 618 (81%) | 167 (19.7%) | 1239 (93.6%) | 4403 (72.8%) |
| Some | 136 (11.3%) | 524 (49.7%) | 29 (3.4%) | 138 (18.1%) | 675 (79.7%) | 68 (5.1%) | 1570 (25.9%) |
| Severe | 1 (0.1%) | 49 (4.6%) | 0 (0%) | 7 (0.9%) | 5 (0.6%) | 17 (1.3%) | 79 (1.3%) |
| Chronic comorbidity<sup>3</sup> | 0 (0%) | 6 (0.6%) | 7 (0.8%) | 17 (2.2%) | 28 (3.3%) | 6 (0.5%) | 64 (1.1%) |
| Acute comorbidity<sup>4</sup> | 84 (7%) | 782 (74.1%) | 457 (53.4%) | 187 (24.5%) | 265 (31.3%) | 555 (41.9%) | 2330 (38.5%) |
| Diarrhea severity (MVS)<sup>5</sup> |  |  |  |  |  |  |  |
| Mild | 612 (50.7%) | 514 (48.7%) | 558 (65.2%) | 455 (59.6%) | 350 (41.3%) | 838 (63.3%) | 3327 (55%) |
| Moderate | 158 (13.1%) | 211 (20%) | 153 (17.9%) | 129 (16.9%) | 217 (25.6%) | 236 (17.8%) | 1104 (18.2%) |
| Severe | 437 (36.2%) | 330 (31.3%) | 145 (16.9%) | 179 (23.5%) | 280 (33.1%) | 250 (18.9%) | 1621 (26.8%) |
| Underweight<sup>6</sup> |  |  |  |  |  |  |  |
| None | 973 (80.6%) | 986 (93.5%) | 762 (89%) | 444 (58.2%) | 762 (90%) | 866 (65.4%) | 4793 (79.2%) |
| Moderate | 185 (15.3%) | 58 (5.5%) | 75 (8.8%) | 193 (25.3%) | 73 (8.6%) | 333 (25.2%) | 917 (15.2%) |
| Severe | 49 (4.1%) | 11 (1%) | 19 (2.2%) | 126 (16.5%) | 12 (1.4%) | 125 (9.4%) | 342 (5.7%) |
| Wasting<sup>7</sup> |  |  |  |  |  |  |  |
| None | 1004 (83.2%) | 1015 (96.2%) | 814 (95.1%) | 504 (66.1%) | 808 (95.4%) | 952 (71.9%) | 5097 (84.2%) |
| Moderate | 175 (14.5%) | 33 (3.1%) | 36 (4.2%) | 177 (23.2%) | 34 (4%) | 290 (21.9%) | 745 (12.3%) |
| Severe | 28 (2.3%) | 7 (0.7%) | 6 (0.7%) | 82 (10.7%) | 5 (0.6%) | 82 (6.2%) | 210 (3.5%) |
| Stunting<sup>8</sup> |  |  |  |  |  |  |  |
| Not stunted | 928 (76.9%) | 866 (82.1%) | 651 (76.1%) | 471 (61.7%) | 677 (79.9%) | 1051 (79.4%) | 4644 (76.7%) |
| Stunted | 279 (23.1%) | 189 (17.9%) | 205 (23.9%) | 292 (38.3%) | 170 (20.1%) | 273 (20.6%) | 1408 (23.3%) |
| Time taken to seek care |  |  |  |  |  |  |  |
| <= 1 day | 270 (22.4%) | 545 (51.7%) | 438 (51.2%) | 256 (33.6%) | 223 (26.3%) | 654 (49.4%) | 2386 (39.4%) |
| > 1 day | 937 (77.6%) | 510 (48.3%) | 418 (48.8%) | 507 (66.4%) | 624 (73.7%) | 670 (50.6%) | 3666 (60.6%) |
| Prior medical care sought | 783 (64.9%) | 90 (8.5%) | 146 (17.1%) | 116 (15.2%) | 178 (21%) | 79 (6%) | 1392 (23%) |

| Prior care seeking location* |  |  |  |  |  |  |  |
| --- | --- | --- | --- | --- | --- | --- | --- |
| Traditional or religious healer | 1 (0.1%) | 28 (2.7%) | 1 (0.1%) | 2 (0.3%) | 1 (0.1%) | 2 (0.2%) | 35 (0.6%) |
| Drug seller | 312 (25.8%) | 11 (1%) | 58 (6.8%) | 3 (0.4%) | 0 (0%) | 2 (0.2%) | 386 (6.4%) |
| Pharmacist | 7 (0.6%) | 26 (2.5%) | 28 (3.3%) | 0 (0%) | 79 (9.3%) | 26 (2%) | 166 (2.7%) |
| Community health worker | 3 (0.2%) | 7 (0.7%) | 5 (0.6%) | 1 (0.1%) | 1 (0.1%) | 5 (0.4%) | 22 (0.4%) |
| Health outpost | 61 (5.1%) | 2 (0.2%) | 2 (0.2%) | 20 (2.6%) | 5 (0.6%) | 17 (1.3%) | 107 (1.8%) |
| Health facility (outpatient care) | 396 (32.8%) | 15 (1.4%) | 32 (3.7%) | 75 (9.8%) | 65 (7.7%) | 30 (2.3%) | 613 (10.1%) |
| Health facility (Inpatient care) | 10 (0.8%) | 0 (0%) | 0 (0%) | 3 (0.4%) | 7 (0.8%) | 0 (0%) | 20 (0.3%) |
| Other | 4 (0.3%) | 4 (0.4%) | 20 (2.3%) | 12 (1.6%) | 23 (2.7%) | 2 (0.2%) | 65 (1.1%) |
| Sociodemographic characteristics |  |  |  |  |  |  |  |
| Caregiver age |  |  |  |  |  |  |  |
| 15-24 years | 554 (45.9%) | 469 (44.5%) | 418 (48.8%) | 255 (33.4%) | 380 (44.9%) | 436 (32.9%) | 2512 (41.5%) |
| 25-65 years | 653 (54.1%) | 585 (55.5%) | 437 (51.1%) | 507 (66.4%) | 467 (55.1%) | 885 (66.8%) | 3534 (58.4%) |
| 65 years and over | 0 (0%) | 1 (0.1%) | 1 (0.1%) | 1 (0.1%) | 0 (0%) | 3 (0.2%) | 6 (0.1%) |
| Mother's highest education received |  |  |  |  |  |  |  |
| None | 143 (11.8%) | 5 (0.5%) | 20 (2.3%) | 238 (31.2%) | 2 (0.2%) | 460 (34.7%) | 868 (14.3%) |
| Primary or less | 284 (23.5%) | 465 (44.1%) | 328 (38.3%) | 186 (24.4%) | 133 (15.7%) | 185 (14%) | 1581 (26.1%) |
| Secondary or above | 780 (64.6%) | 585 (55.5%) | 508 (59.3%) | 324 (42.5%) | 712 (84.1%) | 241 (18.2%) | 3150 (52%) |
| Koranic | 0 (0%) | 0 (0%) | 0 (0%) | 15 (2%) | 0 (0%) | 438 (33.1%) | 453 (7.5%) |
| Father's highest education received |  |  |  |  |  |  |  |
| None | 157 (13%) | 5 (0.5%) | 5 (0.6%) | 244 (32%) | 1 (0.1%) | 335 (25.3%) | 747 (12.3%) |
| Primary or less | 315 (26.1%) | 387 (36.7%) | 122 (14.3%) | 155 (20.3%) | 56 (6.6%) | 109 (8.2%) | 1144 (18.9%) |
| Secondary or above | 735 (60.9%) | 663 (62.8%) | 729 (85.2%) | 355 (46.5%) | 790 (93.3%) | 372 (28.1%) | 3644 (60.2%) |
| Koranic | 0 (0%) | 0 (0%) | 0 (0%) | 9 (1.2%) | 0 (0%) | 508 (38.4%) | 517 (8.5%) |
| Improved drinking water source<sup>9</sup> | 1205 (99.8%) | 704 (66.7%) | 850 (99.3%) | 762 (99.9%) | 766 (90.4%) | 1308 (98.8%) | 5595 (92.4%) |
| Drinking water treatment* |  |  |  |  |  |  |  |
| Boil | 453 (37.5%) | 165<br>(15.6%) | 10 (1.2%) | 239<br>(31.3%) | 193<br>(22.8%) | 0 (0%) | 1060<br>(17.5%) |
| Bleach/chlorination | 2 (0.2%) | 588<br>(55.7%) | 150<br>(17.5%) | 6 (0.8%) | 90<br>(10.6%) | 7 (0.5%) | 843<br>(13.9%) |
| Let stand and settle | 0 (0%) | 14 (1.3%) | 9 (1.1%) | 1 (0.1%) | 13<br>(1.5%) | 0 (0%) | 37<br>(0.6%) |
| Strain through a cloth | 0 (0%) | 7 (0.7%) | 1 (0.1%) | 24<br>(3.1%) | 1 (0.1%) | 45<br>(3.4%) | 78<br>(1.3%) |
| Filter<br>(ceramic/sand/composite/life<br>straw) | 193 (16%) | 1 (0.1%) | 0 (0%) | 24<br>(3.1%) | 2 (0.2%) | 11<br>(0.8%) | 231<br>(3.8%) |
| Improved sanitation<sup>10</sup> | 1204 (99.8%) | 624<br>(59.1%) | 671<br>(78.4%) | 715<br>(93.7%) | 581<br>(68.6%) | 1006<br>(76%) | 4801<br>(79.3%) |
| Shared toilet | 539 (44.7%) | 542<br>(51.4%) | 710<br>(82.9%) | 316<br>(41.4%) | 346<br>(40.9%) | 33<br>(2.5%) | 2486<br>(41.1%) |
| Handwashing practice* |  |  |  |  |  |  |  |
| Before eating | 1174 (97.3%) | 1035<br>(98.1%) | 818<br>(95.6%) | 747<br>(97.9%) | 720<br>(85%) | 1315<br>(99.3%) | 5809<br>(96%) |
| After handling domestic<br>animal | 110 (9.1%) | 339<br>(32.1%) | 43 (5%) | 168<br>(22%) | 108<br>(12.8%) | 867<br>(65.5%) | 1635<br>(27%) |
| After cleaning a child who<br>defecates | 1170 (96.9%) | 671<br>(63.6%) | 790<br>(92.3%) | 640<br>(83.9%) | 567<br>(66.9%) | 1264<br>(95.5%) | 5102<br>(84.3%) |
| Before nursing or preparing<br>baby's food | 1082 (89.6%) | 546<br>(51.8%) | 362<br>(42.3%) | 500<br>(65.5%) | 407<br>(48.1%) | 1173<br>(88.6%) | 4070<br>(67.3%) |
| After defecation | 1173 (97.2%) | 973 (92.2%) | 773 (90.3%) | 560 (73.4%) | 741 (87.5%) | 1276 (96.4%) | 5496 (90.8%) |
| Other | 0 (0%) | 52 (4.9%) | 170 (19.9%) | 0 (0%) | 6 (0.7%) | 0 (0%) | 228 (3.8%) |
| Soap used when handwashing |  |  |  |  |  |  |  |
| Yes | 1115 (92.4%) | 970 (91.9%) | 597 (69.7%) | 684 (89.6%) | 839 (99.1%) | 1258 (95%) | 5463 (90.3%) |
| No | 92 (7.6%) | 85 (8.1%) | 259 (30.3%) | 79 (10.4%) | 8 (0.9%) | 66 (5%) | 589 (9.7%) |
| Wealth quintile<sup>11</sup> |  |  |  |  |  |  |  |
| Quintile 1 (fewest assets) | 0 (0%) | 24 (2.3%) | 94 (11%) | 267 (35%) | 385 (45.5%) | 324 (24.5%) | 1094 (18.1%) |
| Quintile 2 | 60 (5%) | 195 (18.5%) | 261 (30.5%) | 233 (30.5%) | 329 (38.8%) | 563 (42.5%) | 1641 (27.1%) |
| Quintile 3 | 235 (19.5%) | 296 (28.1%) | 340 (39.7%) | 126 (16.5%) | 127 (15%) | 340 (25.7%) | 1464 (24.2%) |
| Quintile 4 | 382 (31.6%) | 390 (37%) | 125 (14.6%) | 104 (13.6%) | 6 (0.7%) | 97 (7.3%) | 1104 (18.2%) |
| Quintile 5 (most assets) | 530 (43.9%) | 150 (14.2%) | 36 (4.2%) | 33 (4.3%) | 0 (0%) | 0 (0%) | 749 (12.4%) |

<sup>1</sup>Child first consumed non-breastmilk fluids or foods at 6 months of age or later
<sup>2</sup>Child received all age-appropriate key vaccines according to national immunization schedule, allowing for 1 month delay. Includes BCG, polio, DPT, hepatitis B, hib, rotavirus, MMR, and pneumococcal pneumonia
<sup>3</sup>Child had any of the following at enrollment: HIV, sickle cell, tuberculosis, or caregiver reports the child has ever been diagnosed with any of the following: rheumatic heart disease, leukemia, sickle cell anemia, tuberculosis, HIV, or other chronic condition.
<sup>4</sup>Child had any of the following diagnoses at enrollment: anemia, urinary tract infection, malaria, suspected sepsis, fever of unknown origin/febrile illness, upper respiratory tract infection, pneumonia, other lower respiratory tract infections, meningitis, acutely unwell -unknown cause, poisoning/herbal intoxication, asthma, soft tissue/skin infection.
<sup>5</sup>Modified Vesikari Score: Defined as in PATH Vesikari Clinical Severity Scoring System Manual.
<sup>6</sup>Severely underweight: WAZ < -3 , Moderately underweight: $-3 \leq \text{WAZ} < -2$ .
<sup>7</sup>Severe wasting: WLZ < -3 or MUAC <11.5, Moderate wasting: $-3 \leq \text{WLZ} < -2$ or $11.5 \leq \text{MUAC} < 12.5$ .
<sup>8</sup>Stunted: LAZ/HAZ < -2, Not stunted: LAZ/HAZ $\geq -2$ .
<sup>9</sup>Drinking water source was classified according to WHO/UNICEF Joint Monitoring Programme (JMP) for Water Supply, Sanitation and Hygiene as improved (water piped into dwelling, yard or plot, public taps or standpipes, tube wells or boreholes, protected dug wells or springs, rainwater collection, filtered or unfiltered water kiosks, bottled water, or sachet water) or surface water/unimproved (water from a river, dam, lake, pond, stream, canal or irrigation channel, unprotected wells or springs).
<sup>10</sup>Sanitation access was classified according to the JMP as improved (flush toilets to piped sewer systems, septic tanks or unknown locations, ventilated improved pit latrines, pit latrines with slab floors, or composting toilets) or open defecation/unimproved (no facility, bush or field, flush toilets to elsewhere, pit latrines without slab floor, bucket toilets or hanging toilets/latrines).
<sup>11</sup>Defined as five categories of the wealth index that align with either urban or national wealth quintiles, depending on whether the catchment area is located in an urban or rural/mixed urban-rural setting.
\*Non-mutually exclusive categories.

Among the 6,052 participants, 53.9% were male, and 46% were aged 12-23 months. A total of 3,527 (58.3%) children were reported to be exclusively breastfed, and 2,829 (46.7%) received all age-appropriate vaccinations, with the highest coverage (77.7%) reported from Bangladesh. Nearly one in six children (14.4%) exhibited blood in their stool, approximately half of them (48%) experienced vomiting, and almost three-quarters (71.5%) had fever. Overall, 72.8% of the children experienced no signs of dehydration, 25.9% experienced some dehydration, and 1.3% presented with severe dehydration. The highest and lowest proportions of some dehydration were reported from Peru (79.7%) and Malawi (3.4%), respectively. Severe dehydration was observed in 1.3% of children. Acute comorbidities were identified in 38.5% of the children, where Kenya (74.1%) reported the highest rate. According to the modified Vesikari score, 55%, 18.2%, and 26.8% of children presented with mild, moderate, and severe diarrhea, respectively. Nutritional assessments indicated that 5.7% of children were severely underweight, 3.5% were severely wasted, and 23.3% were stunted. Regarding healthcare-seeking behavior, overall, 60.6% of caregivers sought care more than one day after symptom development, and 23% of caregivers had a history of seeking prior medical care before coming to the EFGH health facilities. The highest education received by mothers was secondary or above for 52% of individuals. Overall, 92.4% households had access to improved water source and 79.3% use improved sanitation. Despite high overall access to improved water sources and sanitation, Kenya reported a lower figure, with 66.7% for water sources and 59.1% for sanitation. Overall, the handwashing behaviour showed 84.3% of caregivers wash their hands after cleaning a child who defecated, and 67.3% of caregivers practice hand washing before nursing or preparing the baby’s food (Table 2).

In this analysis, various host, clinical, and sociodemographic factors were assessed to identify the association with diarrhea-related hospitalization (Table 3). The risk of hospitalization was 31% higher for male children than females (aRR, 1.31; 95% CI, 1.04-1.64; p=0.019). The risk of hospitalization was 57% lower among those aged 24-35 months (aRR, 0.43; 95% CI, 0.29-0.64; p<0.001) compared to 6-11 months. Vomiting at enrollment was associated with a 2.53 times higher risk of hospitalization (aRR, 2.53; 95% CI, 1.98-3.22; p<0.001), and for fever, the hospitalization risk was 2.24 times higher (aRR, 2.24; 95% CI, 1.62-3.10; p<0.001) compared to children who did not exhibit these symptoms. In the case of some dehydration, the risk of hospitalization was 4.38 times higher (aRR, 4.38; 95% CI, 3.39-5.65; p<0.001) and for severe dehydration, the risk was 17.4 times higher (aRR, 17.40; 95% CI, 12.74-23.78; p<0.001) compared to the children with no signs of dehydration. Chronic comorbidity was also a significant risk factor for hospitalization; the risk was 2.61 times higher (aRR, 2.61; 95% CI, 1.02-6.69; p=0.044) compared to children with no comorbidity. The risk of hospitalization increased with diarrhea severity; a moderate severity score was associated with a 95% higher risk (aRR, 1.95; 95% CI, 1.36-2.80; p<0.001), and a severe score carried a 4.79 times higher risk (aRR, 4.79; 95% CI, 3.69-6.20; p<0.001) compared to mild diarrhea. The risk of hospitalization was 35% higher (aRR, 1.35; 95% CI, 1-1.81; p=0.048) for moderately underweight children and 96% higher (aRR, 1.96; 95% CI, 1.32-2.89; p<0.001) for those severely underweight compared to children who were not underweight. For severe wasting, the risk of hospitalization was 2.62 times higher (aRR, 2.62; 95% CI, 1.78-3.87; p<0.001) than the children with no wasting. The risk of hospitalization was 49% higher (aRR, 1.49; 95%CI, 1.11-2.01; p=0.007) for children whose caregivers sought medical care before attending the EFGH facility than those who did not. A higher risk of hospitalization was observed for those who used a filter (aRR, 1.51; 95% CI, 1.01-2.26; p=0.046) and strain of cloth (aRR, 1.98; 95% CI, 1.01-3.85; p=0.045) for treating drinking water in comparison to those who did not treat water using these methods. Children of caregivers who washed their hands after cleaning a child who defecated (aRR, 1.76; 95% CI, 1.14-2.74; p=0.011) had 76% higher risk of hospitalization compared to children of caregivers who did not report washing their hands. Also, the children of caregivers who washed their hands before nursing or preparing the baby’s food had 60% higher (aRR, 1.60; 95% CI, 1.16-2.21; p=0.004) risk of hospitalization in comparison to those who did not report washing their hands (Table 3).

**Table 3.**
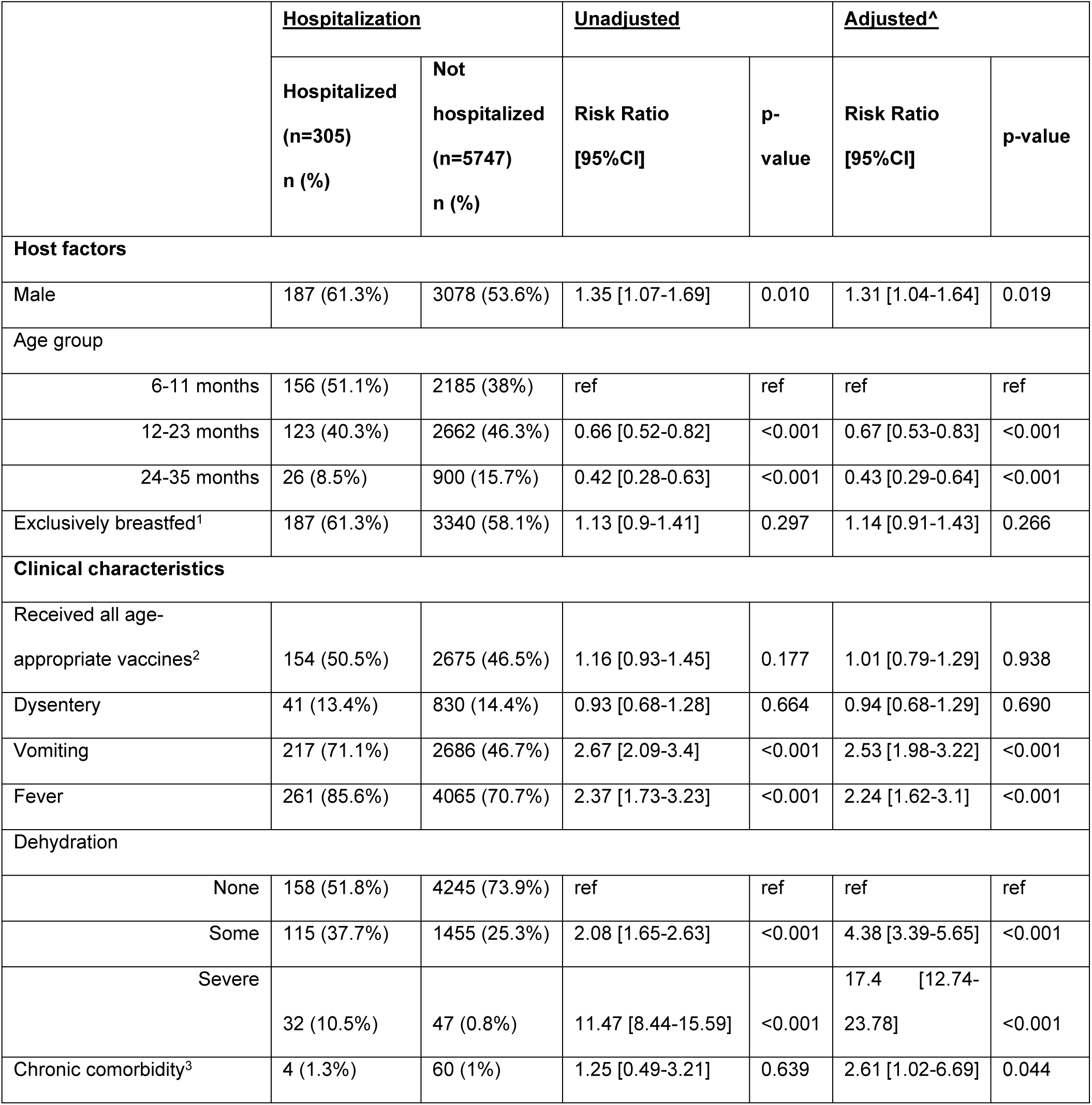
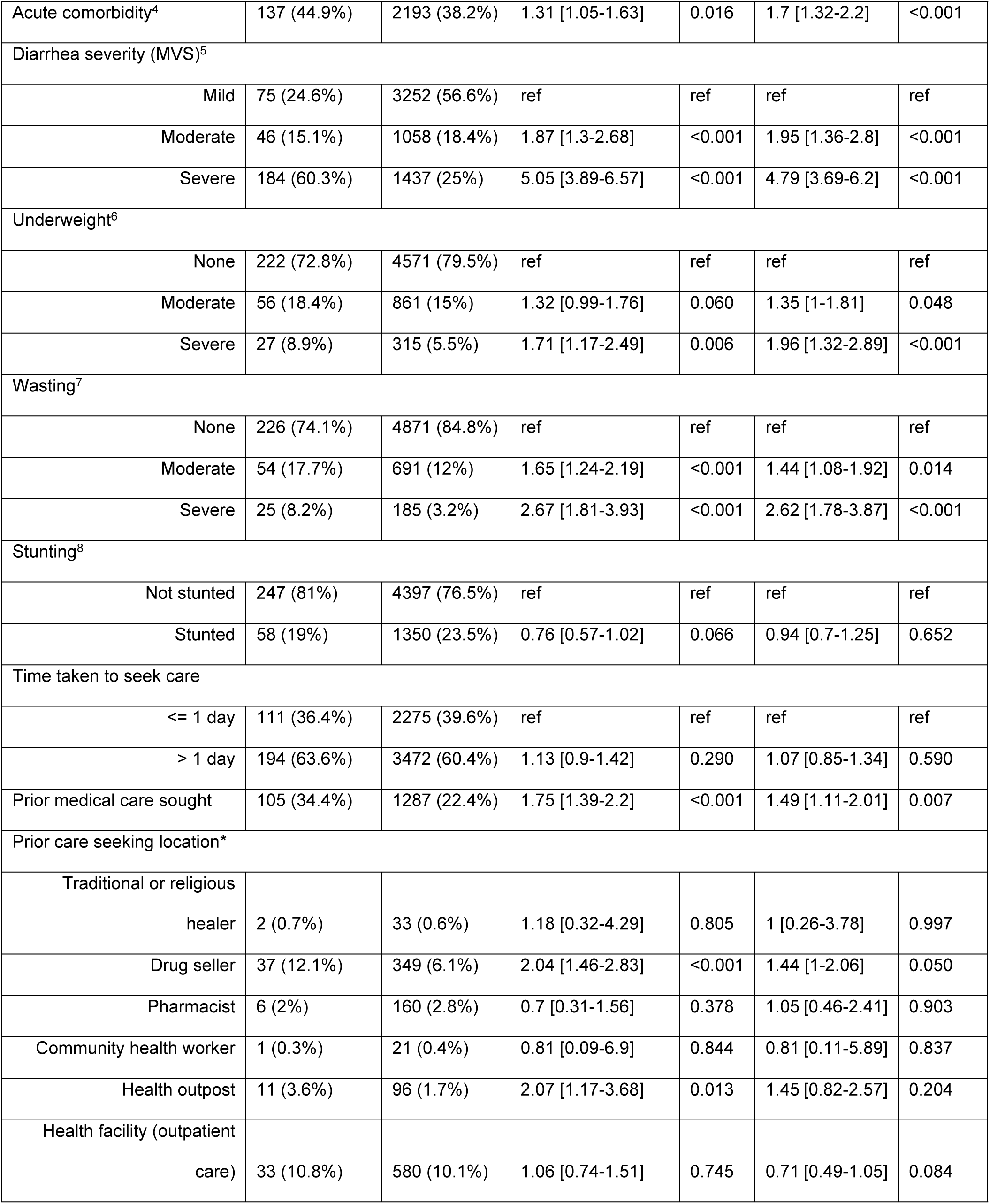
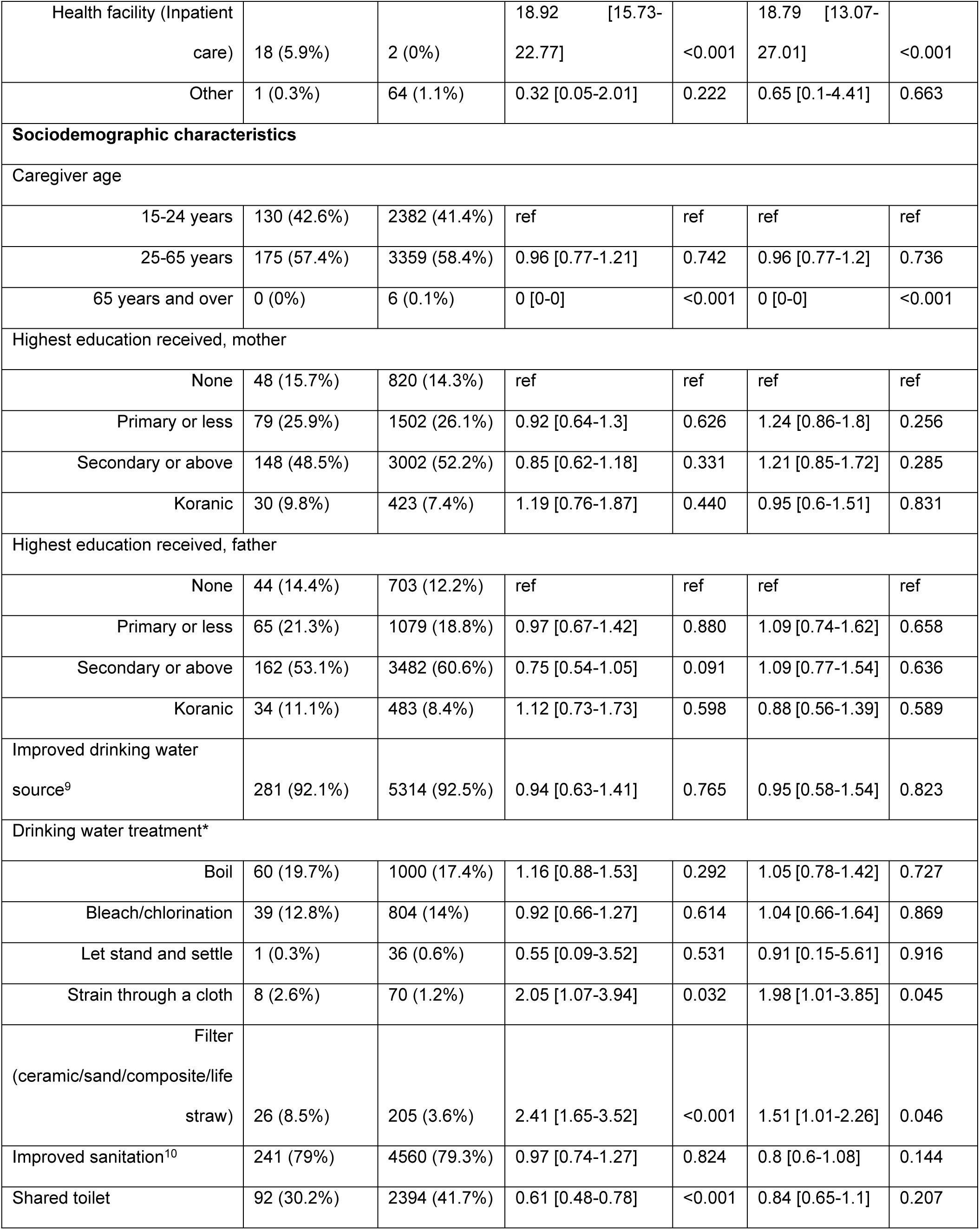
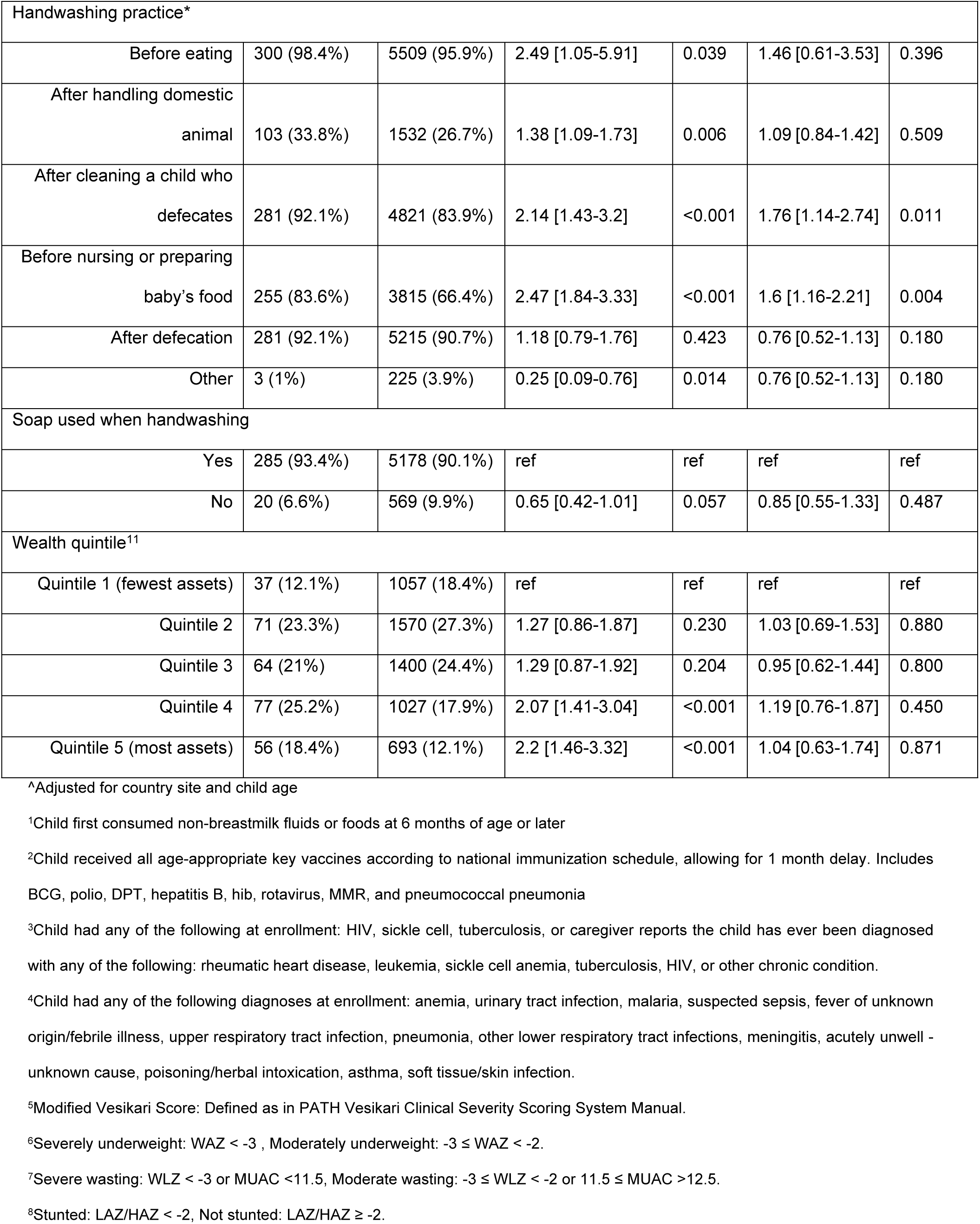
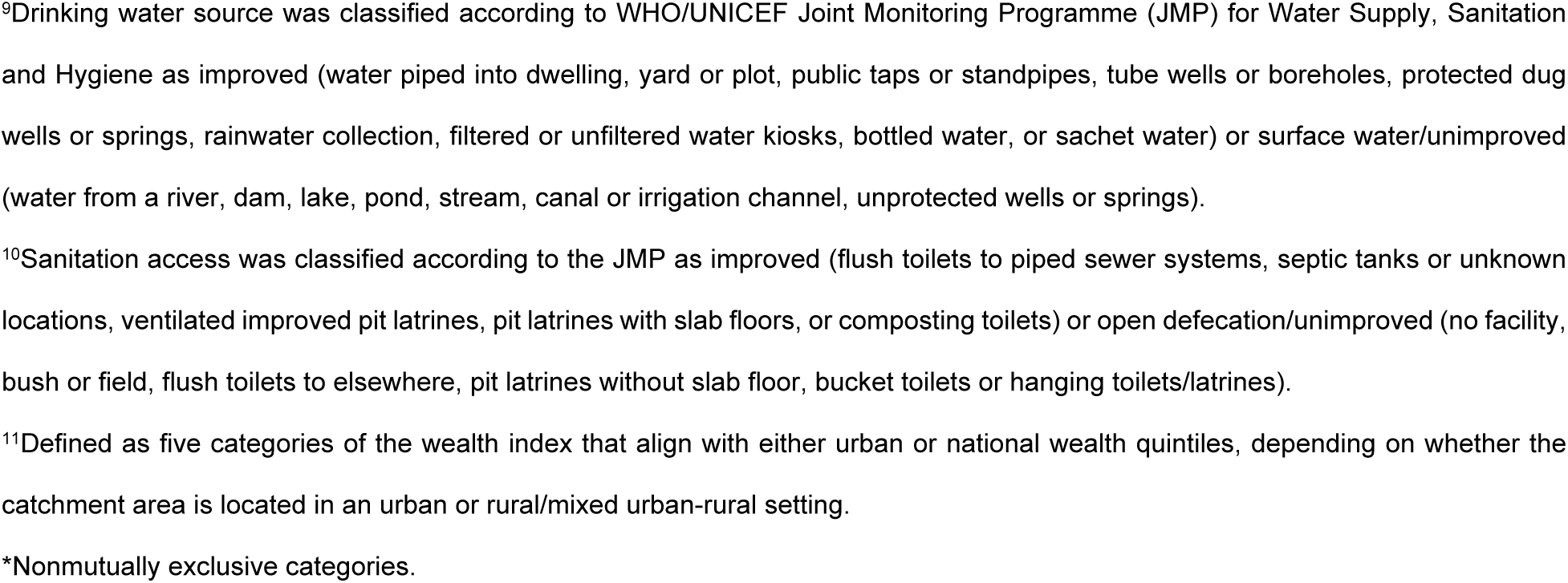
Host, clinical and sociodemographic factors of hospitalization among children aged 6-35 months

|  | <u>Hospitalization</u> |  | <u>Unadjusted</u> |  | <u>Adjusted<sup>^</sup></u> |  |
| --- | --- | --- | --- | --- | --- | --- |
|  | Hospitalized<br>(n=305)<br>n (%) | Not<br>hospitalized<br>(n=5747)<br>n (%) | Risk Ratio<br>[95%CI] | p-value | Risk Ratio<br>[95%CI] | p-value |
| **Host factors** |  |  |  |  |  |  |
| Male | 187 (61.3%) | 3078 (53.6%) | 1.35 [1.07-1.69] | 0.010 | 1.31 [1.04-1.64] | 0.019 |
| Age group |  |  |  |  |  |  |
| 6-11 months | 156 (51.1%) | 2185 (38%) | ref | ref | ref | ref |
| 12-23 months | 123 (40.3%) | 2662 (46.3%) | 0.66 [0.52-0.82] | <0.001 | 0.67 [0.53-0.83] | <0.001 |
| 24-35 months | 26 (8.5%) | 900 (15.7%) | 0.42 [0.28-0.63] | <0.001 | 0.43 [0.29-0.64] | <0.001 |
| Exclusively breastfed<sup>1</sup> | 187 (61.3%) | 3340 (58.1%) | 1.13 [0.9-1.41] | 0.297 | 1.14 [0.91-1.43] | 0.266 |
| **Clinical characteristics** |  |  |  |  |  |  |
| Received all age-appropriate vaccines<sup>2</sup> | 154 (50.5%) | 2675 (46.5%) | 1.16 [0.93-1.45] | 0.177 | 1.01 [0.79-1.29] | 0.938 |
| Dysentery | 41 (13.4%) | 830 (14.4%) | 0.93 [0.68-1.28] | 0.664 | 0.94 [0.68-1.29] | 0.690 |
| Vomiting | 217 (71.1%) | 2686 (46.7%) | 2.67 [2.09-3.4] | <0.001 | 2.53 [1.98-3.22] | <0.001 |
| Fever | 261 (85.6%) | 4065 (70.7%) | 2.37 [1.73-3.23] | <0.001 | 2.24 [1.62-3.1] | <0.001 |
| Dehydration |  |  |  |  |  |  |
| None | 158 (51.8%) | 4245 (73.9%) | ref | ref | ref | ref |
| Some | 115 (37.7%) | 1455 (25.3%) | 2.08 [1.65-2.63] | <0.001 | 4.38 [3.39-5.65] | <0.001 |
| Severe | 32 (10.5%) | 47 (0.8%) | 11.47 [8.44-15.59] | <0.001 | 17.4 [12.74-23.78] | <0.001 |
| Chronic comorbidity<sup>3</sup> | 4 (1.3%) | 60 (1%) | 1.25 [0.49-3.21] | 0.639 | 2.61 [1.02-6.69] | 0.044 |
| Acute comorbidity<sup>4</sup> | 137 (44.9%) | 2193 (38.2%) | 1.31 [1.05-1.63] | 0.016 | 1.7 [1.32-2.2] | <0.001 |
| Diarrhea severity (MVS)<sup>5</sup> |  |  |  |  |  |  |
| Mild | 75 (24.6%) | 3252 (56.6%) | ref | ref | ref | ref |
| Moderate | 46 (15.1%) | 1058 (18.4%) | 1.87 [1.3-2.68] | <0.001 | 1.95 [1.36-2.8] | <0.001 |
| Severe | 184 (60.3%) | 1437 (25%) | 5.05 [3.89-6.57] | <0.001 | 4.79 [3.69-6.2] | <0.001 |
| Underweight<sup>6</sup> |  |  |  |  |  |  |
| None | 222 (72.8%) | 4571 (79.5%) | ref | ref | ref | ref |
| Moderate | 56 (18.4%) | 861 (15%) | 1.32 [0.99-1.76] | 0.060 | 1.35 [1-1.81] | 0.048 |
| Severe | 27 (8.9%) | 315 (5.5%) | 1.71 [1.17-2.49] | 0.006 | 1.96 [1.32-2.89] | <0.001 |
| Wasting<sup>7</sup> |  |  |  |  |  |  |
| None | 226 (74.1%) | 4871 (84.8%) | ref | ref | ref | ref |
| Moderate | 54 (17.7%) | 691 (12%) | 1.65 [1.24-2.19] | <0.001 | 1.44 [1.08-1.92] | 0.014 |
| Severe | 25 (8.2%) | 185 (3.2%) | 2.67 [1.81-3.93] | <0.001 | 2.62 [1.78-3.87] | <0.001 |
| Stunting<sup>8</sup> |  |  |  |  |  |  |
| Not stunted | 247 (81%) | 4397 (76.5%) | ref | ref | ref | ref |
| Stunted | 58 (19%) | 1350 (23.5%) | 0.76 [0.57-1.02] | 0.066 | 0.94 [0.7-1.25] | 0.652 |
| Time taken to seek care |  |  |  |  |  |  |
| <= 1 day | 111 (36.4%) | 2275 (39.6%) | ref | ref | ref | ref |
| > 1 day | 194 (63.6%) | 3472 (60.4%) | 1.13 [0.9-1.42] | 0.290 | 1.07 [0.85-1.34] | 0.590 |
| Prior medical care sought | 105 (34.4%) | 1287 (22.4%) | 1.75 [1.39-2.2] | <0.001 | 1.49 [1.11-2.01] | 0.007 |
| Prior care seeking location* |  |  |  |  |  |  |
| Traditional or religious healer | 2 (0.7%) | 33 (0.6%) | 1.18 [0.32-4.29] | 0.805 | 1 [0.26-3.78] | 0.997 |
| Drug seller | 37 (12.1%) | 349 (6.1%) | 2.04 [1.46-2.83] | <0.001 | 1.44 [1-2.06] | 0.050 |
| Pharmacist | 6 (2%) | 160 (2.8%) | 0.7 [0.31-1.56] | 0.378 | 1.05 [0.46-2.41] | 0.903 |
| Community health worker | 1 (0.3%) | 21 (0.4%) | 0.81 [0.09-6.9] | 0.844 | 0.81 [0.11-5.89] | 0.837 |
| Health outpost | 11 (3.6%) | 96 (1.7%) | 2.07 [1.17-3.68] | 0.013 | 1.45 [0.82-2.57] | 0.204 |
| Health facility (outpatient care) | 33 (10.8%) | 580 (10.1%) | 1.06 [0.74-1.51] | 0.745 | 0.71 [0.49-1.05] | 0.084 |
| Health facility (Inpatient care) | 18 (5.9%) | 2 (0%) | 18.92 [15.73-22.77] | <0.001 | 18.79 [13.07-27.01] | <0.001 |
| Other | 1 (0.3%) | 64 (1.1%) | 0.32 [0.05-2.01] | 0.222 | 0.65 [0.1-4.41] | 0.663 |
| **Sociodemographic characteristics** |  |  |  |  |  |  |
| Caregiver age |  |  |  |  |  |  |
| 15-24 years | 130 (42.6%) | 2382 (41.4%) | ref | ref | ref | ref |
| 25-65 years | 175 (57.4%) | 3359 (58.4%) | 0.96 [0.77-1.21] | 0.742 | 0.96 [0.77-1.2] | 0.736 |
| 65 years and over | 0 (0%) | 6 (0.1%) | 0 [0-0] | <0.001 | 0 [0-0] | <0.001 |
| Highest education received, mother |  |  |  |  |  |  |
| None | 48 (15.7%) | 820 (14.3%) | ref | ref | ref | ref |
| Primary or less | 79 (25.9%) | 1502 (26.1%) | 0.92 [0.64-1.3] | 0.626 | 1.24 [0.86-1.8] | 0.256 |
| Secondary or above | 148 (48.5%) | 3002 (52.2%) | 0.85 [0.62-1.18] | 0.331 | 1.21 [0.85-1.72] | 0.285 |
| Koranic | 30 (9.8%) | 423 (7.4%) | 1.19 [0.76-1.87] | 0.440 | 0.95 [0.6-1.51] | 0.831 |
| Highest education received, father |  |  |  |  |  |  |
| None | 44 (14.4%) | 703 (12.2%) | ref | ref | ref | ref |
| Primary or less | 65 (21.3%) | 1079 (18.8%) | 0.97 [0.67-1.42] | 0.880 | 1.09 [0.74-1.62] | 0.658 |
| Secondary or above | 162 (53.1%) | 3482 (60.6%) | 0.75 [0.54-1.05] | 0.091 | 1.09 [0.77-1.54] | 0.636 |
| Koranic | 34 (11.1%) | 483 (8.4%) | 1.12 [0.73-1.73] | 0.598 | 0.88 [0.56-1.39] | 0.589 |
| Improved drinking water source<sup>9</sup> | 281 (92.1%) | 5314 (92.5%) | 0.94 [0.63-1.41] | 0.765 | 0.95 [0.58-1.54] | 0.823 |
| Drinking water treatment* |  |  |  |  |  |  |
| Boil | 60 (19.7%) | 1000 (17.4%) | 1.16 [0.88-1.53] | 0.292 | 1.05 [0.78-1.42] | 0.727 |
| Bleach/chlorination | 39 (12.8%) | 804 (14%) | 0.92 [0.66-1.27] | 0.614 | 1.04 [0.66-1.64] | 0.869 |
| Let stand and settle | 1 (0.3%) | 36 (0.6%) | 0.55 [0.09-3.52] | 0.531 | 0.91 [0.15-5.61] | 0.916 |
| Strain through a cloth | 8 (2.6%) | 70 (1.2%) | 2.05 [1.07-3.94] | 0.032 | 1.98 [1.01-3.85] | 0.045 |
| Filter (ceramic/sand/composite/life straw) | 26 (8.5%) | 205 (3.6%) | 2.41 [1.65-3.52] | <0.001 | 1.51 [1.01-2.26] | 0.046 |
| Improved sanitation<sup>10</sup> | 241 (79%) | 4560 (79.3%) | 0.97 [0.74-1.27] | 0.824 | 0.8 [0.6-1.08] | 0.144 |
| Shared toilet | 92 (30.2%) | 2394 (41.7%) | 0.61 [0.48-0.78] | <0.001 | 0.84 [0.65-1.1] | 0.207 |
| Handwashing practice* |  |  |  |  |  |  |
| Before eating | 300 (98.4%) | 5509 (95.9%) | 2.49 [1.05-5.91] | 0.039 | 1.46 [0.61-3.53] | 0.396 |
| After handling domestic animal | 103 (33.8%) | 1532 (26.7%) | 1.38 [1.09-1.73] | 0.006 | 1.09 [0.84-1.42] | 0.509 |
| After cleaning a child who defecates | 281 (92.1%) | 4821 (83.9%) | 2.14 [1.43-3.2] | <0.001 | 1.76 [1.14-2.74] | 0.011 |
| Before nursing or preparing baby's food | 255 (83.6%) | 3815 (66.4%) | 2.47 [1.84-3.33] | <0.001 | 1.6 [1.16-2.21] | 0.004 |
| After defecation | 281 (92.1%) | 5215 (90.7%) | 1.18 [0.79-1.76] | 0.423 | 0.76 [0.52-1.13] | 0.180 |
| Other | 3 (1%) | 225 (3.9%) | 0.25 [0.09-0.76] | 0.014 | 0.76 [0.52-1.13] | 0.180 |
| Soap used when handwashing |  |  |  |  |  |  |
| Yes | 285 (93.4%) | 5178 (90.1%) | ref | ref | ref | ref |
| No | 20 (6.6%) | 569 (9.9%) | 0.65 [0.42-1.01] | 0.057 | 0.85 [0.55-1.33] | 0.487 |
| Wealth quintile<sup>11</sup> |  |  |  |  |  |  |
| Quintile 1 (fewest assets) | 37 (12.1%) | 1057 (18.4%) | ref | ref | ref | ref |
| Quintile 2 | 71 (23.3%) | 1570 (27.3%) | 1.27 [0.86-1.87] | 0.230 | 1.03 [0.69-1.53] | 0.880 |
| Quintile 3 | 64 (21%) | 1400 (24.4%) | 1.29 [0.87-1.92] | 0.204 | 0.95 [0.62-1.44] | 0.800 |
| Quintile 4 | 77 (25.2%) | 1027 (17.9%) | 2.07 [1.41-3.04] | <0.001 | 1.19 [0.76-1.87] | 0.450 |
| Quintile 5 (most assets) | 56 (18.4%) | 693 (12.1%) | 2.2 [1.46-3.32] | <0.001 | 1.04 [0.63-1.74] | 0.871 |

<sup>11</sup>Adjusted for country site and child age
<sup>1</sup>Child first consumed non-breastmilk fluids or foods at 6 months of age or later
<sup>2</sup>Child received all age-appropriate key vaccines according to national immunization schedule, allowing for 1 month delay. Includes BCG, polio, DPT, hepatitis B, hib, rotavirus, MMR, and pneumococcal pneumonia
<sup>3</sup>Child had any of the following at enrollment: HIV, sickle cell, tuberculosis, or caregiver reports the child has ever been diagnosed with any of the following: rheumatic heart disease, leukemia, sickle cell anemia, tuberculosis, HIV, or other chronic condition.
<sup>4</sup>Child had any of the following diagnoses at enrollment: anemia, urinary tract infection, malaria, suspected sepsis, fever of unknown origin/febrile illness, upper respiratory tract infection, pneumonia, other lower respiratory tract infections, meningitis, acutely unwell - unknown cause, poisoning/herbal intoxication, asthma, soft tissue/skin infection.
<sup>5</sup>Modified Vesikari Score: Defined as in PATH Vesikari Clinical Severity Scoring System Manual.
<sup>6</sup>Severely underweight: WAZ < -3 , Moderately underweight: -3 ≤ WAZ < -2.
<sup>7</sup>Severe wasting: WLZ < -3 or MUAC <11.5, Moderate wasting: -3 ≤ WLZ < -2 or 11.5 ≤ MUAC >12.5.
<sup>8</sup>Stunted: LAZ/HAZ < -2, Not stunted: LAZ/HAZ ≥ -2.
<sup>9</sup>Drinking water source was classified according to WHO/UNICEF Joint Monitoring Programme (JMP) for Water Supply, Sanitation and Hygiene as improved (water piped into dwelling, yard or plot, public taps or standpipes, tube wells or boreholes, protected dug wells or springs, rainwater collection, filtered or unfiltered water kiosks, bottled water, or sachet water) or surface water/unimproved (water from a river, dam, lake, pond, stream, canal or irrigation channel, unprotected wells or springs).
<sup>10</sup>Sanitation access was classified according to the JMP as improved (flush toilets to piped sewer systems, septic tanks or unknown locations, ventilated improved pit latrines, pit latrines with slab floors, or composting toilets) or open defecation/unimproved (no facility, bush or field, flush toilets to elsewhere, pit latrines without slab floor, bucket toilets or hanging toilets/latrines).
<sup>11</sup>Defined as five categories of the wealth index that align with either urban or national wealth quintiles, depending on whether the catchment area is located in an urban or rural/mixed urban-rural setting.
\*Nonmutually exclusive categories.

In site-specific chi-square and Fisher’s exact test analyses, hospitalization outcomes differed significantly by diarrhea severity in Bangladesh (χ²(1, N = 1207) = 80.31, p<0.001), Kenya (χ²(2, N = 1055) = 47.08, p<0.001), Malawi (Fisher’s exact, p=0.035), Pakistan (Fisher’s exact, p<0.001), and The Gambia (χ²(2, N = 1324) = 43.24, p<0.001). The proportion of children hospitalized also differed by vomiting at enrollment in Bangladesh (χ²(1, N = 1207) = 31.58, p<0.001), Kenya (χ²(1, N = 1055) = 17.23, p<0.001), Pakistan (χ²(1, N = 763) = 12.51, p<0.001), and The Gambia (χ²(1, N = 1324) = 9.13, p=0.002), respectively. Similarly, hospitalization differed by dehydration at enrollment in Bangladesh (Fisher’s exact, p<0.001), Kenya (Fisher’s exact, p<0.001), Pakistan (Fisher’s exact, p=0.012) and The Gambia (Fisher’s exact, p<0.001). These three clinical factors, which had a strongly positive association with hospitalization in the all-site analysis, were not significantly associated in the Peru-specific analysis. Severe wasting was observed in 18.4% of children who were hospitalized at The Gambia site, where a significant association between wasting and hospitalization was observed (χ²(2, N = 1324) = 28.85, p<0.001). In other sites, the number of hospitalized children with severe wasting was low, and no significant association was found. Additional site-specific data are included in the supplementary tables (S1 Table - S6 Table).

## Discussion

Overall, diarrhea-related hospitalizations accounted for 5% across the EFGH study sites. The highest proportion of hospitalizations was reported in Bangladesh, whereas the lowest was seen in Malawi. We found several significant factors associated with an increased risk of diarrhea-related hospitalization, including male gender, younger age, vomiting, fever, some or severe dehydration, comorbidities at enrollment, higher diarrhea severity, moderately or severely underweight at baseline, moderate or severe wasting at baseline, prior seeking of medical care, drinking water treatment methods and handwashing practices (Fig 2).

**Fig 2:**
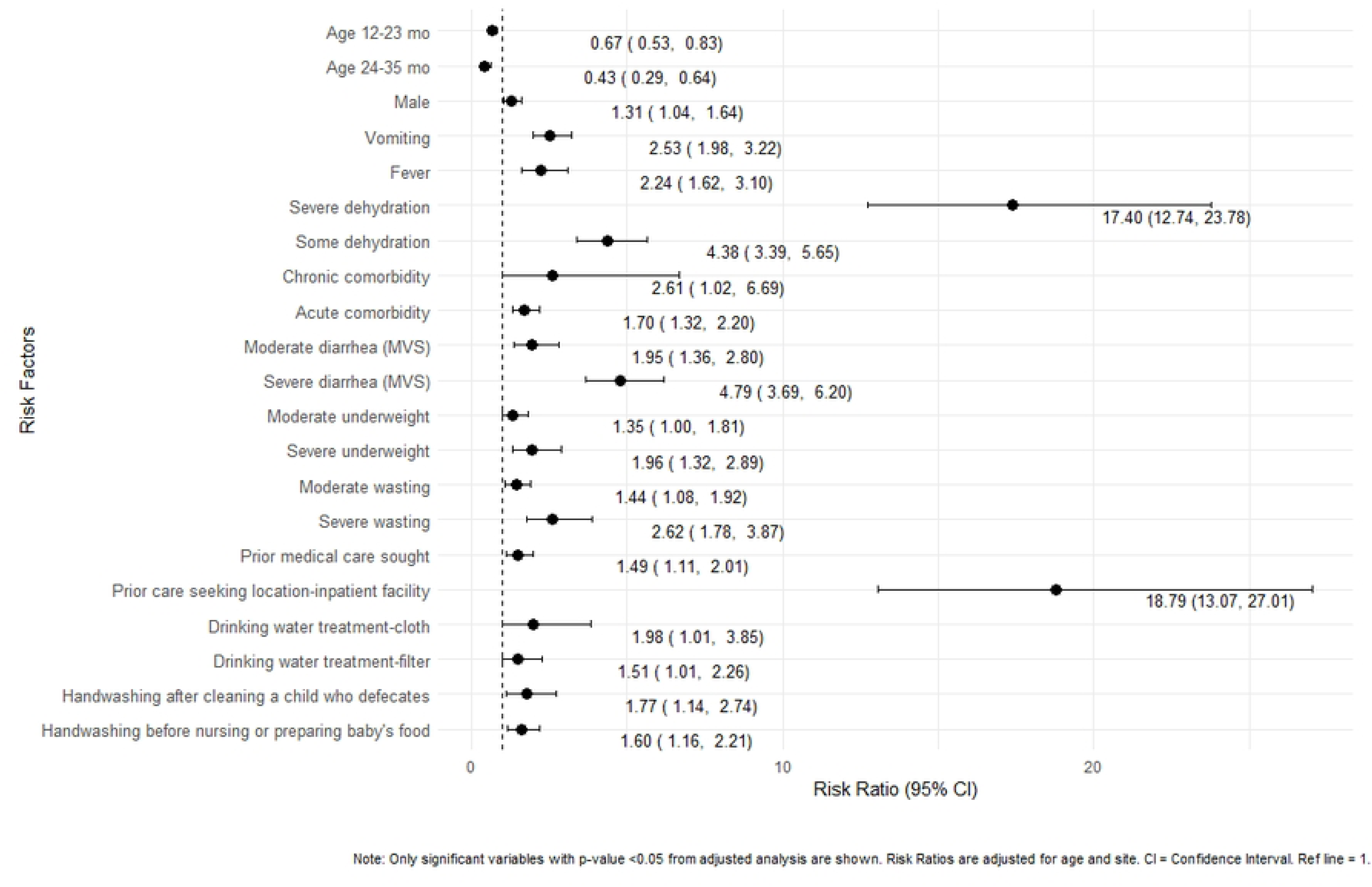
**Risk of hospitalization among children 6-35 months**

Previous studies have highlighted several risk factors for diarrhea-related hospitalization across diverse geographical settings. For instance, a study from Bangladesh emphasized the role of socioeconomic factors, such as education, household income, and environmental conditions, including unsafe water sources and home infrastructure [19]. Studies conducted in Brazil, Iran, and Turkey focused on clinical and demographic factors such as younger age, poor nutritional status, vomiting, dehydration, and a history of previous hospitalization, and identified these as significant risk factors for diarrheal hospitalization [20, 40, 41], which aligns with our findings. We have also observed a higher rate (34.4%) of hospitalization among children who had sought prior medical care before presenting at the EFGH health facilities, as their condition had not improved. The Global Enteric Multicenter Study (GEMS) also underscored the critical importance of timely medical intervention, particularly in averting the progression of dehydration to severe conditions necessitating hospitalization [5].

Nutritional status critically influences health outcomes in children with diarrheal diseases, as malnutrition exacerbates the severity and duration of diarrhea, thereby increasing the risk of hospitalization and mortality[42]. Our study findings reinforced these factors, as moderate and severe underweight and wasting were significantly associated with a higher risk of hospitalization.

Handwashing with soap can significantly reduce hospitalization due to diarrheal diseases[43]. However, we found a positive association between caregivers washing their hands after child defecation and before nursing or preparing the child’s food and the likelihood of hospitalization. This may be because caregivers who are more diligent about hand hygiene are also more proactive in seeking medical care, which could result in a higher rate of hospitalization for their children.

In Malawi and Peru, a higher proportion of children presented with stunting and vomiting but were not hospitalized. Although the prevalence of these symptoms at enrollment was comparable across the other four sites, hospitalization rates were low in both countries: 1.1% in Malawi and 1.2% in Peru. This may be due to sub-optimal healthcare-seeking behavior, inadequate access to quality healthcare, financial barriers, or the presence of chronic conditions. Previous studies have also reported poor quality and low utilization of sick child healthcare services in health facilities, despite the wide availability of curative care in Peru and Malawi [44, 45]. The findings of this study emphasized that improving the quality of these facilities could enhance healthcare utilization and help reduce preventable childhood morbidity and mortality.

Our analysis had several limitations. Firstly, we were limited by a strict definition of hospitalization that was used in the main study [27]. Secondly, the information obtained from caregivers’ interviews was based on memory, which might impose recall bias. Thirdly, the findings may not be generalizable to all children with diarrhea in the community, as this study included only children who sought care at the EFGH health facilities. Despite these limitations, the study had several strengths, including multi-country implementation, rigorous data collection methods, and a recent comprehensive surveillance strategy.

In conclusion, our analysis demonstrated that younger age, male sex, diarrhea severity, malnutrition, delayed care seeking, and WASH variables are the primary risk factors for hospitalization among young children with diarrhea in LMICs. These findings highlight the need for integrated strategies that combine clinical management, hygiene, and nutritional support to reduce hospitalization rates for diarrhea in young children living in resource-limited settings. Such insights can guide the development of effective policies and interventions aimed at improving child health outcomes and alleviating the burden of childhood diarrhea in LMICs.

## Acknowledgments

icddr,b is also grateful to the Governments of Bangladesh and Canada for providing core/unrestricted support and acknowledges with gratitude the commitment of the Gates Foundation to its research efforts. The authors are grateful to the governments of each study site for providing their unrestricted support. The authors thank the children who participated in this study and their families, and the dedicated physicians, nurses, scientists, and staff at each study site for their dedication and outstanding performance of clinical and laboratory study activities. The author gratefully acknowledges the continuous support provided by the EFGH Coordination and the Nyanja Health Research Institute for the EFGH Manuscript Writing Cohort Program.

## Author contribution

M.I., S.R., M.T.I., and F.K. were responsible for drafting this report. S.R. led the data analysis. M.I., S.R., M.T.I., and F.K. led the data interpretation and verified the data presented in the manuscript. M.I., S.R., M.T.I., F.K., B.T.B., P.B.P., H.E.A., B.O.O., B.O., M.J.H., M.J., D.M., M.N., P.P.Y., P.G.B., M.N.H.R., M.I.H., F.A., F.Q., F.N.Q., and S.S. revised the report critically. All authors have full access to all the data in the study. All authors have read and agreed to the published version of the manuscript.

## Potential conflicts of interest

We have no conflicts of interest to disclose.

## Funding

This research was funded by the Gates Foundation (award numbers INV-031791, INV-045988, INV-062665, INV-076498). The funders had no role in study design, data collection and analysis, decision to publish, or preparation of the manuscript.

## Data Availability

The deidentified and anonymized EFGH datasets, data dictionaries, statistical analysis plan, CRFs and study protocol were made publicly available on the Vivli repository in December 2025. Access to the data and supporting documents is available on request at <u>Vivli DOI - PR00011860</u> and requires the execution of a Data Use Agreement.

---

**S1 Table.**
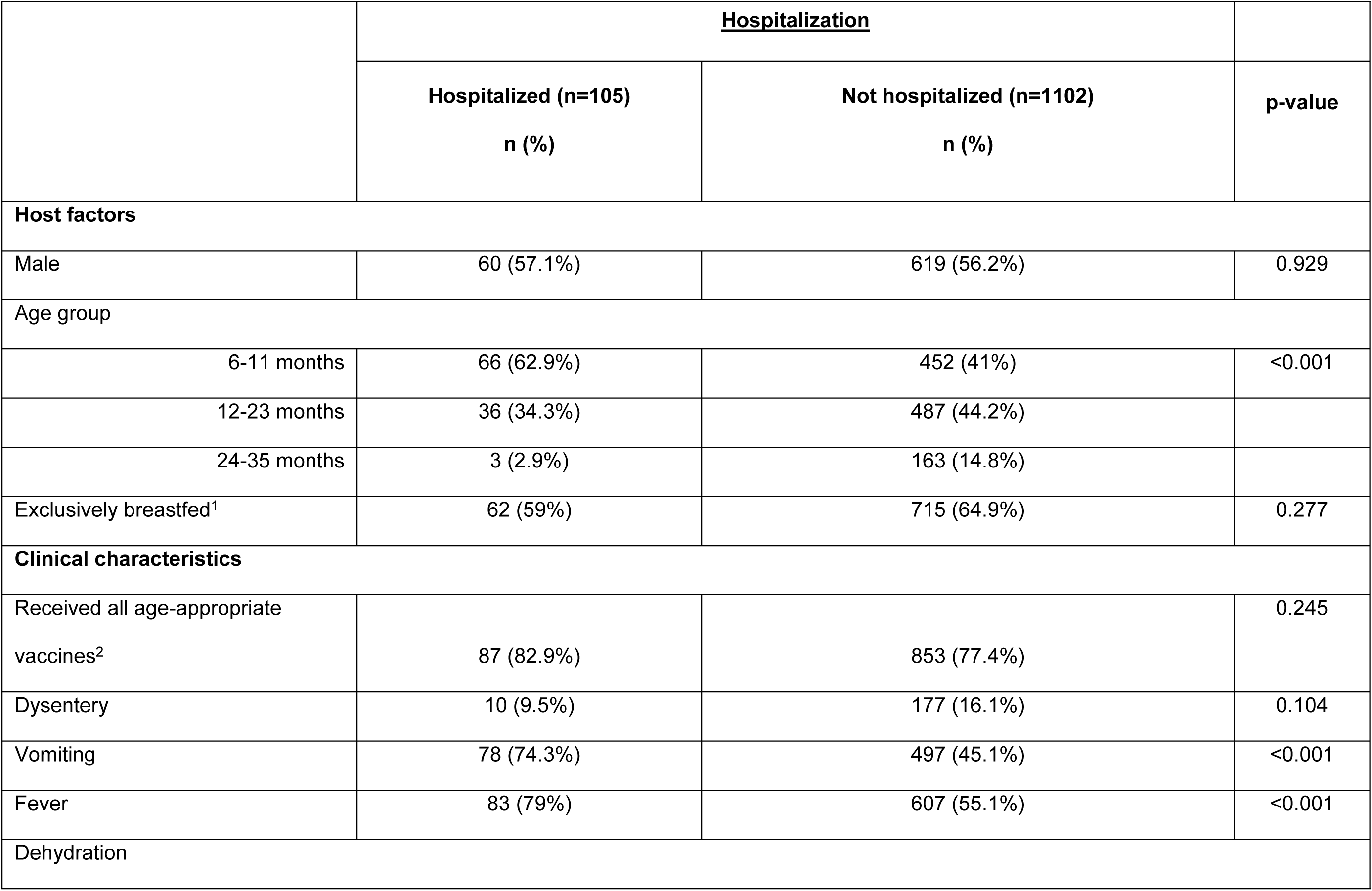
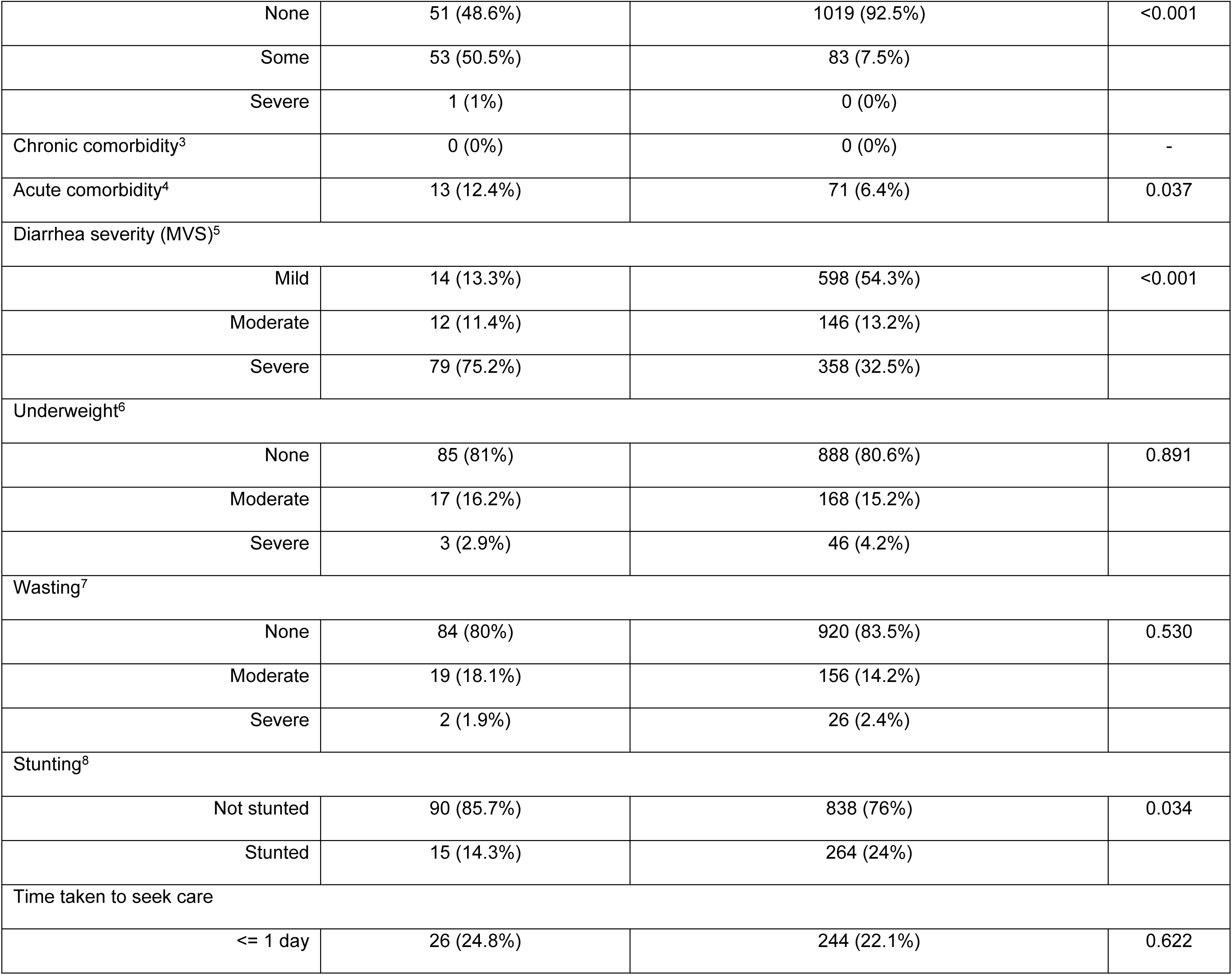
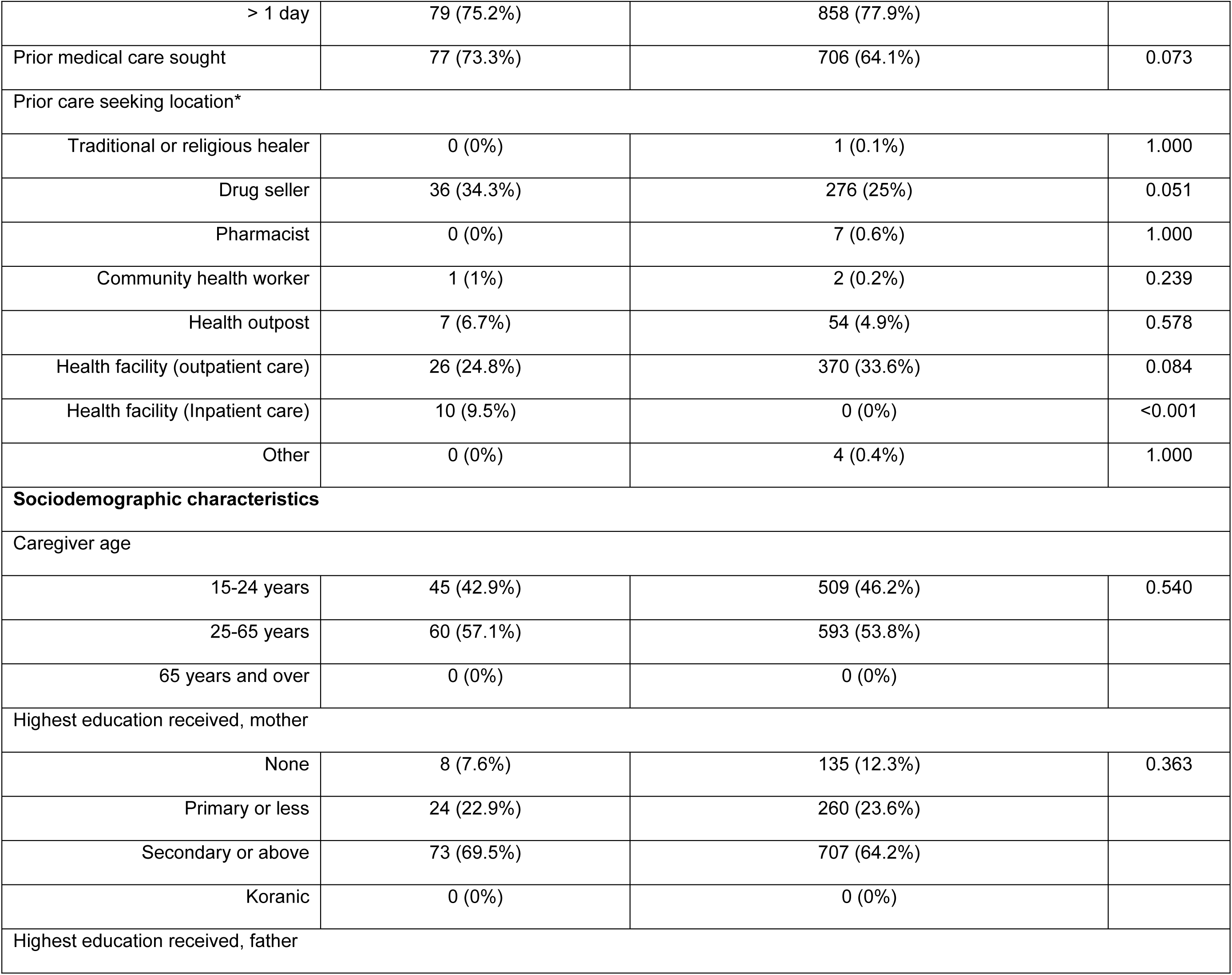
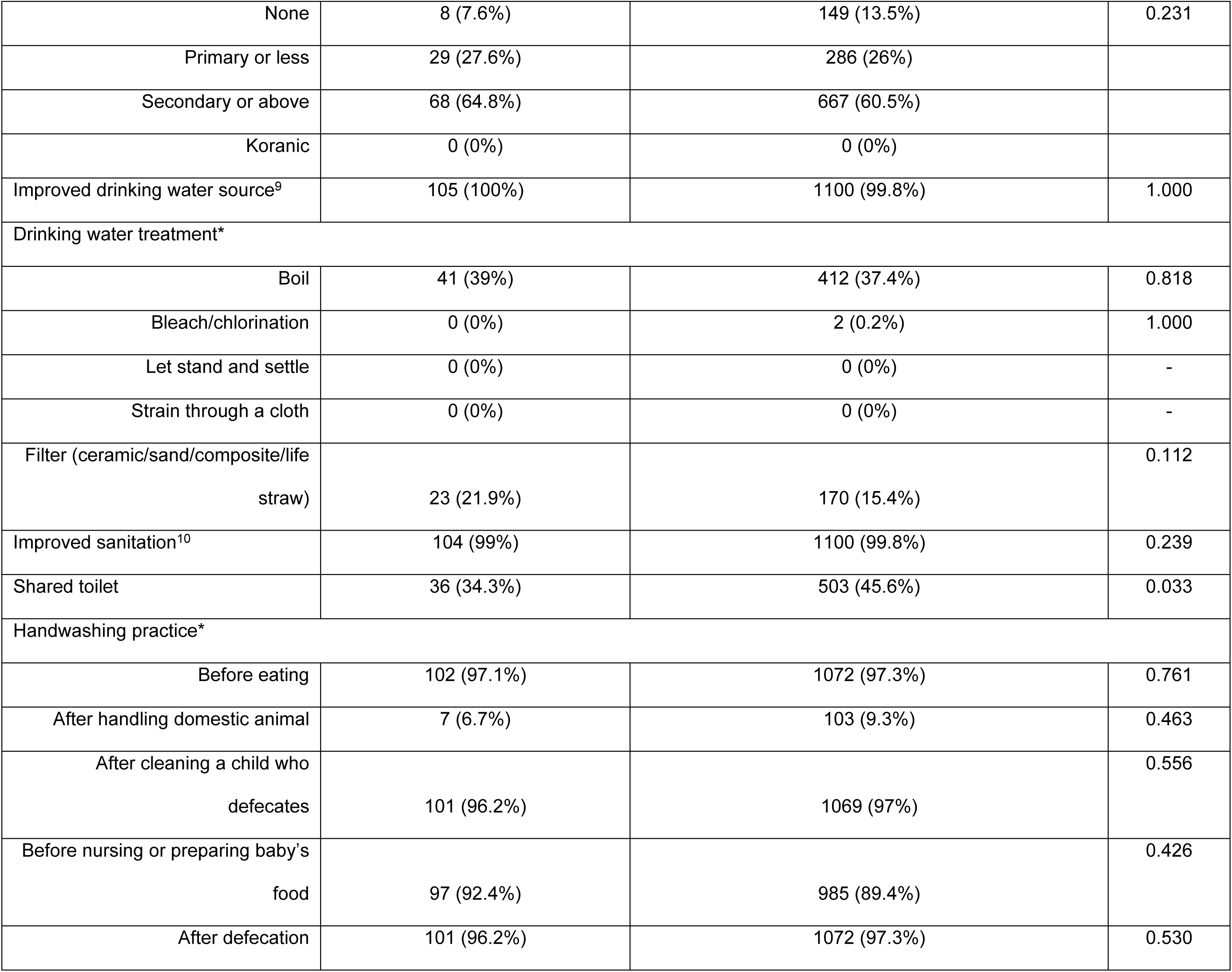
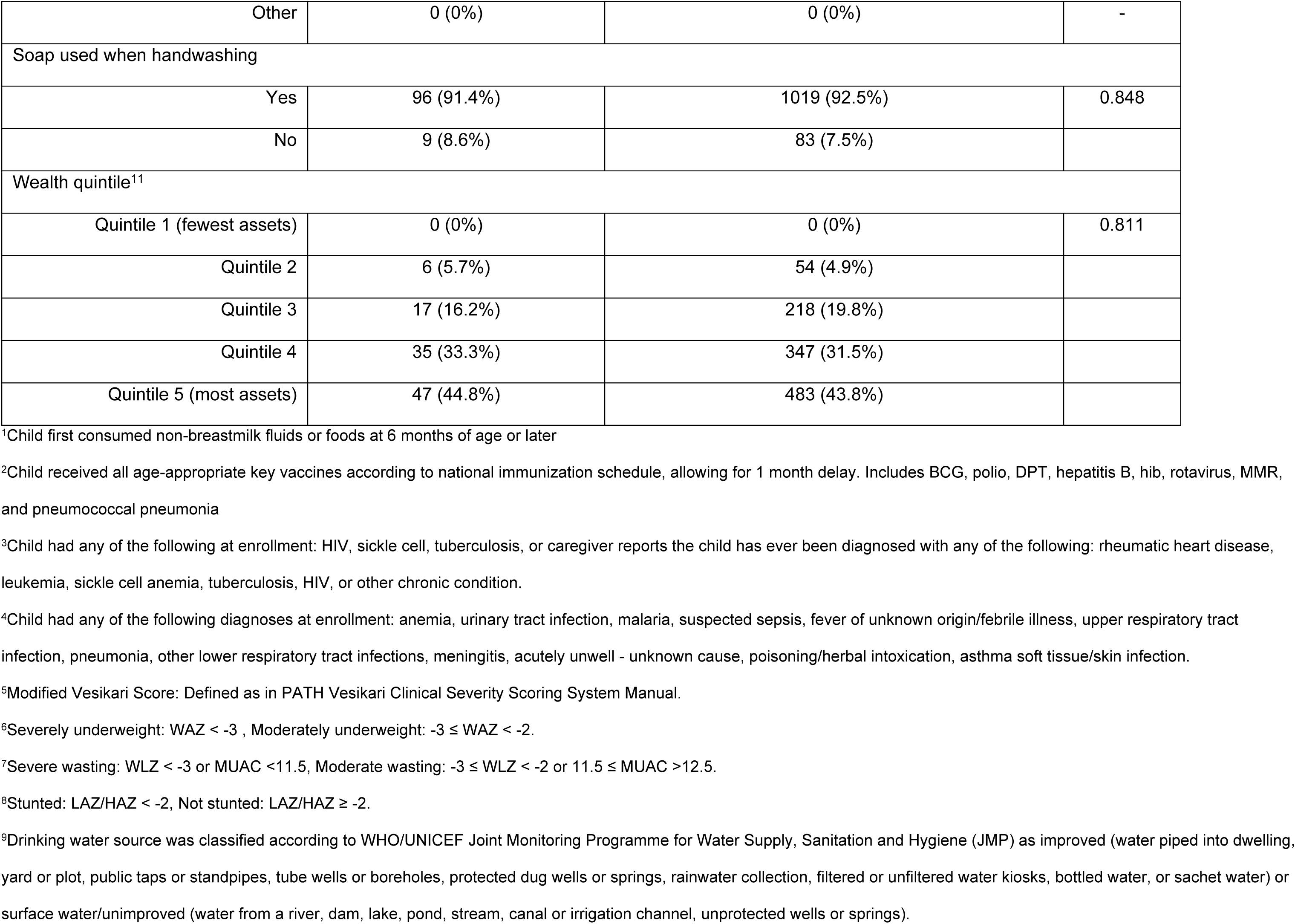
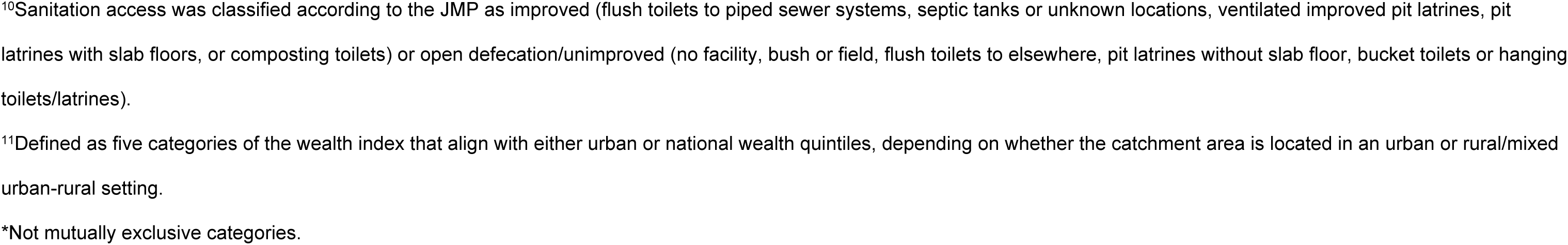
Host, clinical and sociodemographic factors of hospitalization among children aged 6-35 months in Bangladesh

**S2 Table.**
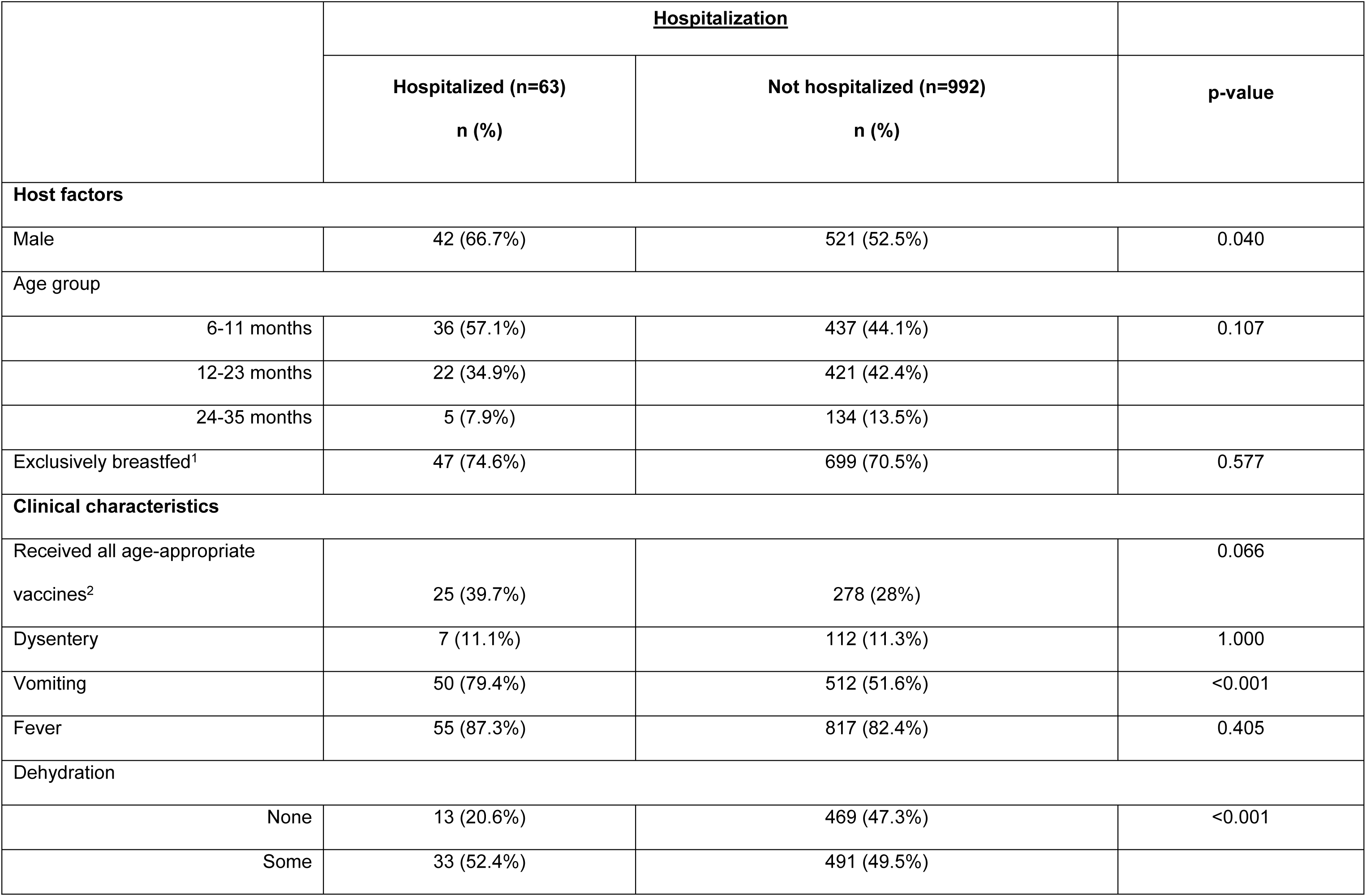
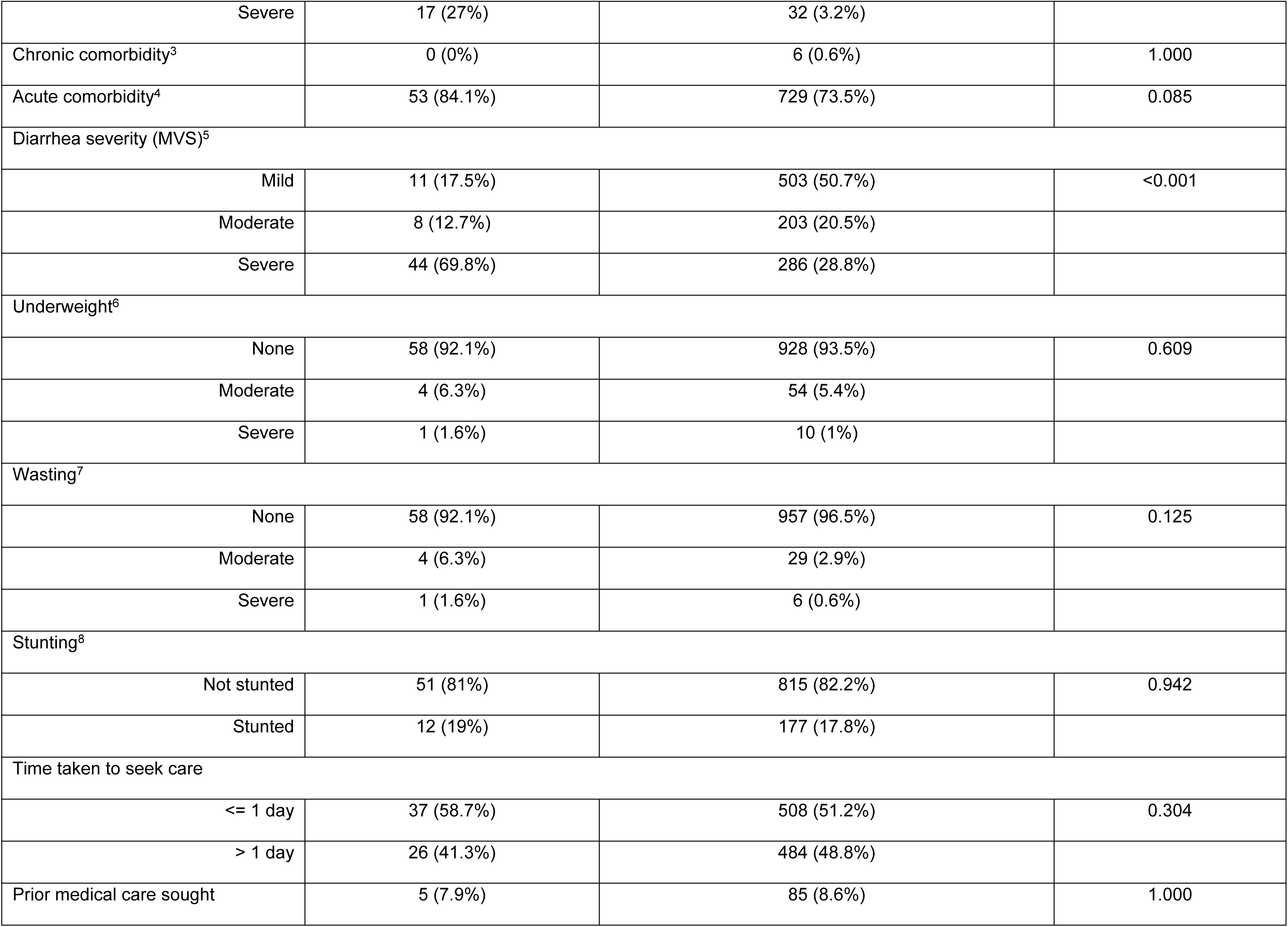
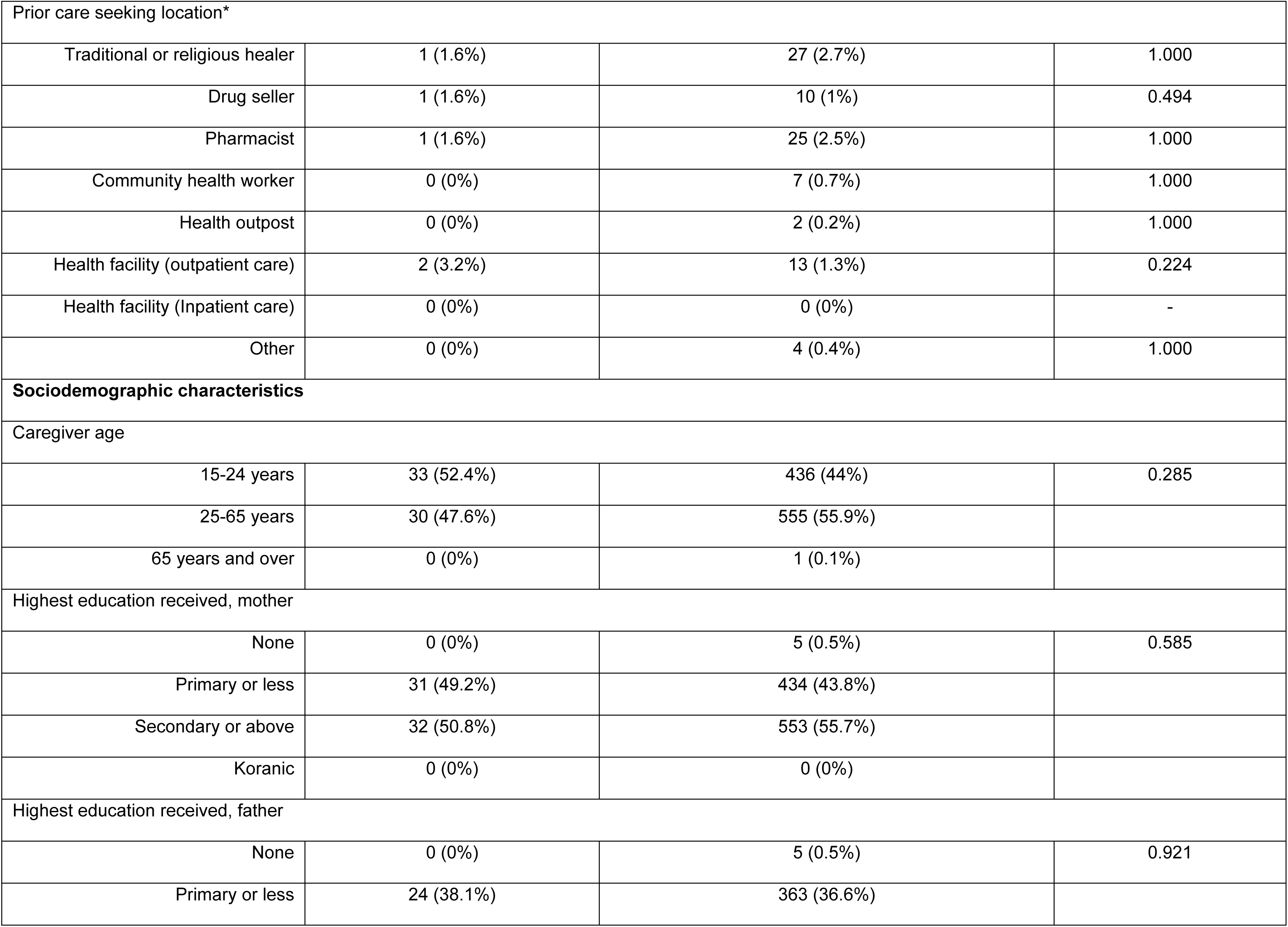
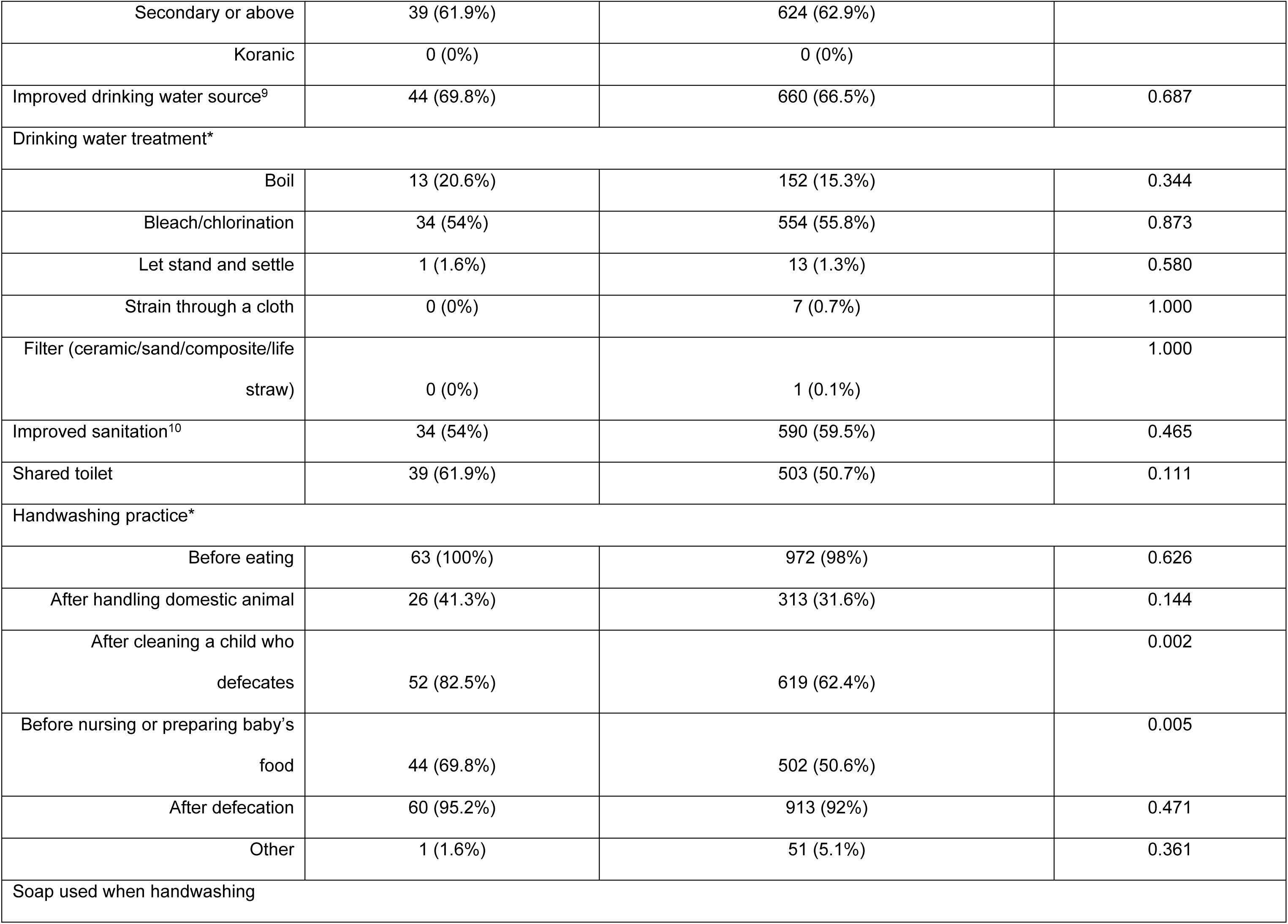
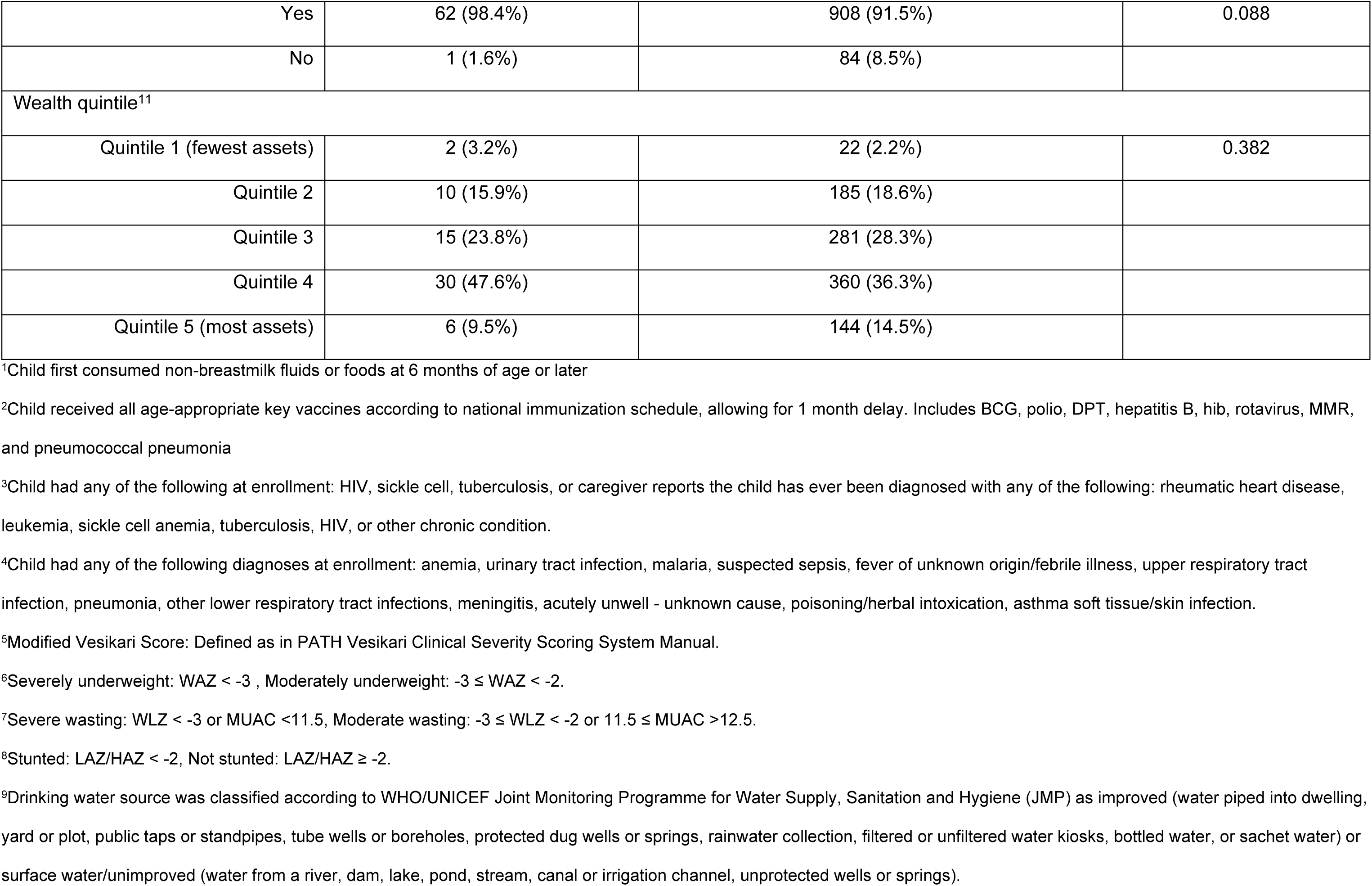
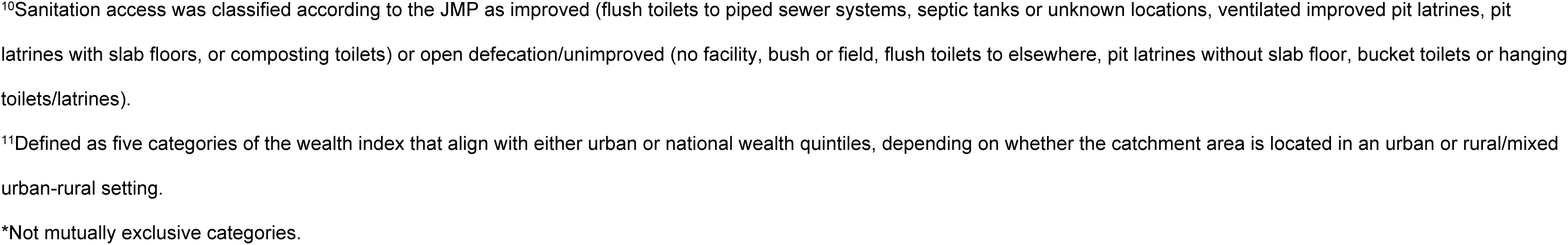
Host, clinical and sociodemographic factors of hospitalization among children aged 6-35 months in Kenya

|  | <u>Hospitalization</u> |  | p-value |
| --- | --- | --- | --- |
|  | Hospitalized (n=63)<br>n (%) | Not hospitalized (n=992)<br>n (%) |  |
| Host factors |  |  |  |
| Male | 42 (66.7%) | 521 (52.5%) | 0.040 |
| Age group |  |  |  |
| 6-11 months | 36 (57.1%) | 437 (44.1%) | 0.107 |
| 12-23 months | 22 (34.9%) | 421 (42.4%) |  |
| 24-35 months | 5 (7.9%) | 134 (13.5%) |  |
| Exclusively breastfed<sup>1</sup> | 47 (74.6%) | 699 (70.5%) | 0.577 |
| Clinical characteristics |  |  |  |
| Received all age-appropriate vaccines<sup>2</sup> | 25 (39.7%) | 278 (28%) | 0.066 |
| Dysentery | 7 (11.1%) | 112 (11.3%) | 1.000 |
| Vomiting | 50 (79.4%) | 512 (51.6%) | <0.001 |
| Fever | 55 (87.3%) | 817 (82.4%) | 0.405 |
| Dehydration |  |  |  |
| None | 13 (20.6%) | 469 (47.3%) | <0.001 |
| Some | 33 (52.4%) | 491 (49.5%) |  |
| Severe | 17 (27%) | 32 (3.2%) |  |
| Chronic comorbidity<sup>3</sup> | 0 (0%) | 6 (0.6%) | 1.000 |
| Acute comorbidity<sup>4</sup> | 53 (84.1%) | 729 (73.5%) | 0.085 |
| Diarrhea severity (MVS)<sup>5</sup> |  |  |  |
| Mild | 11 (17.5%) | 503 (50.7%) | <0.001 |
| Moderate | 8 (12.7%) | 203 (20.5%) |  |
| Severe | 44 (69.8%) | 286 (28.8%) |  |
| Underweight<sup>6</sup> |  |  |  |
| None | 58 (92.1%) | 928 (93.5%) | 0.609 |
| Moderate | 4 (6.3%) | 54 (5.4%) |  |
| Severe | 1 (1.6%) | 10 (1%) |  |
| Wasting<sup>7</sup> |  |  |  |
| None | 58 (92.1%) | 957 (96.5%) | 0.125 |
| Moderate | 4 (6.3%) | 29 (2.9%) |  |
| Severe | 1 (1.6%) | 6 (0.6%) |  |
| Stunting<sup>8</sup> |  |  |  |
| Not stunted | 51 (81%) | 815 (82.2%) | 0.942 |
| Stunted | 12 (19%) | 177 (17.8%) |  |
| Time taken to seek care |  |  |  |
| <= 1 day | 37 (58.7%) | 508 (51.2%) | 0.304 |
| > 1 day | 26 (41.3%) | 484 (48.8%) |  |
| Prior medical care sought | 5 (7.9%) | 85 (8.6%) | 1.000 |
| Prior care seeking location* |  |  |  |
| Traditional or religious healer | 1 (1.6%) | 27 (2.7%) | 1.000 |
| Drug seller | 1 (1.6%) | 10 (1%) | 0.494 |
| Pharmacist | 1 (1.6%) | 25 (2.5%) | 1.000 |
| Community health worker | 0 (0%) | 7 (0.7%) | 1.000 |
| Health outpost | 0 (0%) | 2 (0.2%) | 1.000 |
| Health facility (outpatient care) | 2 (3.2%) | 13 (1.3%) | 0.224 |
| Health facility (Inpatient care) | 0 (0%) | 0 (0%) | - |
| Other | 0 (0%) | 4 (0.4%) | 1.000 |
| **Sociodemographic characteristics** |  |  |  |
| Caregiver age |  |  |  |
| 15-24 years | 33 (52.4%) | 436 (44%) | 0.285 |
| 25-65 years | 30 (47.6%) | 555 (55.9%) |  |
| 65 years and over | 0 (0%) | 1 (0.1%) |  |
| Highest education received, mother |  |  |  |
| None | 0 (0%) | 5 (0.5%) | 0.585 |
| Primary or less | 31 (49.2%) | 434 (43.8%) |  |
| Secondary or above | 32 (50.8%) | 553 (55.7%) |  |
| Koranic | 0 (0%) | 0 (0%) |  |
| Highest education received, father |  |  |  |
| None | 0 (0%) | 5 (0.5%) | 0.921 |
| Primary or less | 24 (38.1%) | 363 (36.6%) |  |
| Secondary or above | 39 (61.9%) | 624 (62.9%) |  |
| Koranic | 0 (0%) | 0 (0%) |  |
| Improved drinking water source<sup>9</sup> | 44 (69.8%) | 660 (66.5%) | 0.687 |
| Drinking water treatment* |  |  |  |
| Boil | 13 (20.6%) | 152 (15.3%) | 0.344 |
| Bleach/chlorination | 34 (54%) | 554 (55.8%) | 0.873 |
| Let stand and settle | 1 (1.6%) | 13 (1.3%) | 0.580 |
| Strain through a cloth | 0 (0%) | 7 (0.7%) | 1.000 |
| Filter (ceramic/sand/composite/life straw) | 0 (0%) | 1 (0.1%) | 1.000 |
| Improved sanitation<sup>10</sup> | 34 (54%) | 590 (59.5%) | 0.465 |
| Shared toilet | 39 (61.9%) | 503 (50.7%) | 0.111 |
| Handwashing practice* |  |  |  |
| Before eating | 63 (100%) | 972 (98%) | 0.626 |
| After handling domestic animal | 26 (41.3%) | 313 (31.6%) | 0.144 |
| After cleaning a child who defecates | 52 (82.5%) | 619 (62.4%) | 0.002 |
| Before nursing or preparing baby's food | 44 (69.8%) | 502 (50.6%) | 0.005 |
| After defecation | 60 (95.2%) | 913 (92%) | 0.471 |
| Other | 1 (1.6%) | 51 (5.1%) | 0.361 |
| Soap used when handwashing |  |  |  |
| Yes | 62 (98.4%) | 908 (91.5%) | 0.088 |
| No | 1 (1.6%) | 84 (8.5%) |  |
| Wealth quintile<sup>11</sup> |  |  |  |
| Quintile 1 (fewest assets) | 2 (3.2%) | 22 (2.2%) | 0.382 |
| Quintile 2 | 10 (15.9%) | 185 (18.6%) |  |
| Quintile 3 | 15 (23.8%) | 281 (28.3%) |  |
| Quintile 4 | 30 (47.6%) | 360 (36.3%) |  |
| Quintile 5 (most assets) | 6 (9.5%) | 144 (14.5%) |  |

<sup>1</sup>Child first consumed non-breastmilk fluids or foods at 6 months of age or later
<sup>2</sup>Child received all age-appropriate key vaccines according to national immunization schedule, allowing for 1 month delay. Includes BCG, polio, DPT, hepatitis B, hib, rotavirus, MMR, and pneumococcal pneumonia
<sup>3</sup>Child had any of the following at enrollment: HIV, sickle cell, tuberculosis, or caregiver reports the child has ever been diagnosed with any of the following: rheumatic heart disease, leukemia, sickle cell anemia, tuberculosis, HIV, or other chronic condition.
<sup>4</sup>Child had any of the following diagnoses at enrollment: anemia, urinary tract infection, malaria, suspected sepsis, fever of unknown origin/febrile illness, upper respiratory tract infection, pneumonia, other lower respiratory tract infections, meningitis, acutely unwell - unknown cause, poisoning/herbal intoxication, asthma soft tissue/skin infection.
<sup>5</sup>Modified Vesikari Score: Defined as in PATH Vesikari Clinical Severity Scoring System Manual.
<sup>6</sup>Severely underweight: WAZ < -3 , Moderately underweight: -3 ≤ WAZ < -2.
<sup>7</sup>Severe wasting: WLZ < -3 or MUAC <11.5, Moderate wasting: -3 ≤ WLZ < -2 or 11.5 ≤ MUAC >12.5.
<sup>8</sup>Stunted: LAZ/HAZ < -2, Not stunted: LAZ/HAZ ≥ -2.
<sup>9</sup>Drinking water source was classified according to WHO/UNICEF Joint Monitoring Programme for Water Supply, Sanitation and Hygiene (JMP) as improved (water piped into dwelling, yard or plot, public taps or standpipes, tube wells or boreholes, protected dug wells or springs, rainwater collection, filtered or unfiltered water kiosks, bottled water, or sachet water) or surface water/unimproved (water from a river, dam, lake, pond, stream, canal or irrigation channel, unprotected wells or springs).
<sup>10</sup>Sanitation access was classified according to the JMP as improved (flush toilets to piped sewer systems, septic tanks or unknown locations, ventilated improved pit latrines, pit latrines with slab floors, or composting toilets) or open defecation/unimproved (no facility, bush or field, flush toilets to elsewhere, pit latrines without slab floor, bucket toilets or hanging toilets/latrines). <sup>11</sup>Defined as five categories of the wealth index that align with either urban or national wealth quintiles, depending on whether the catchment area is located in an urban or rural/mixed urban-rural setting. \*Not mutually exclusive categories.

**S3 Table.**
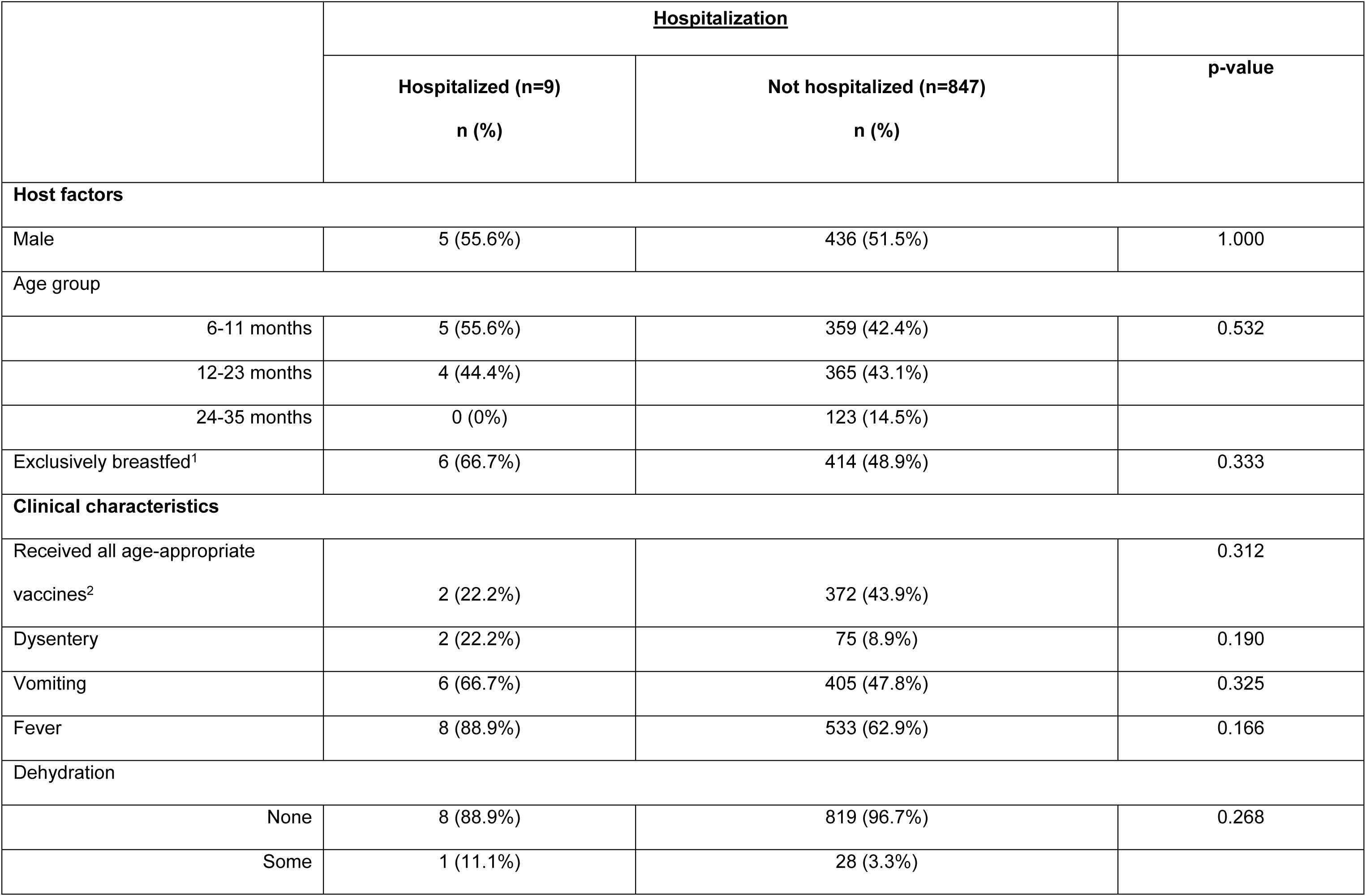
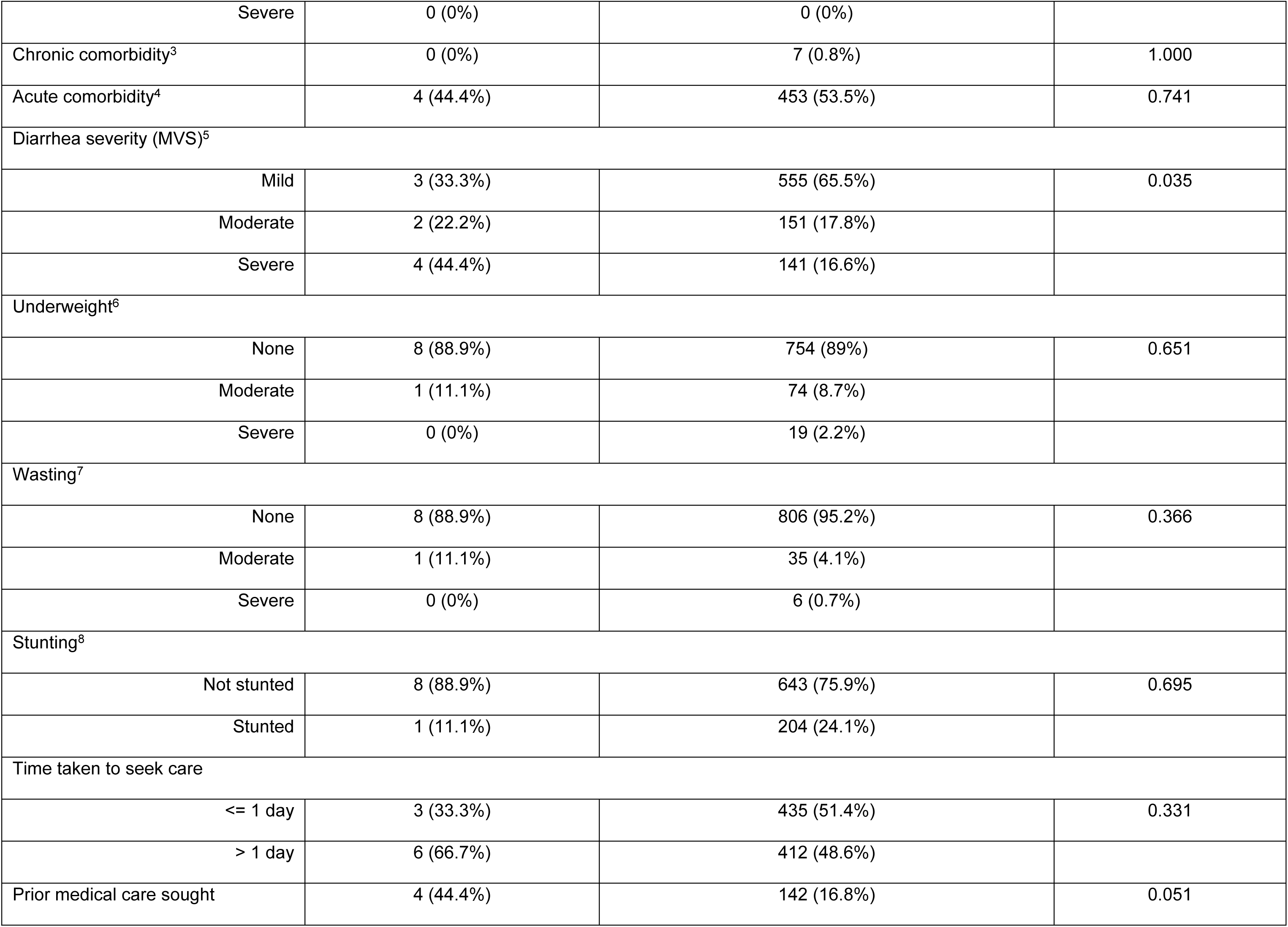
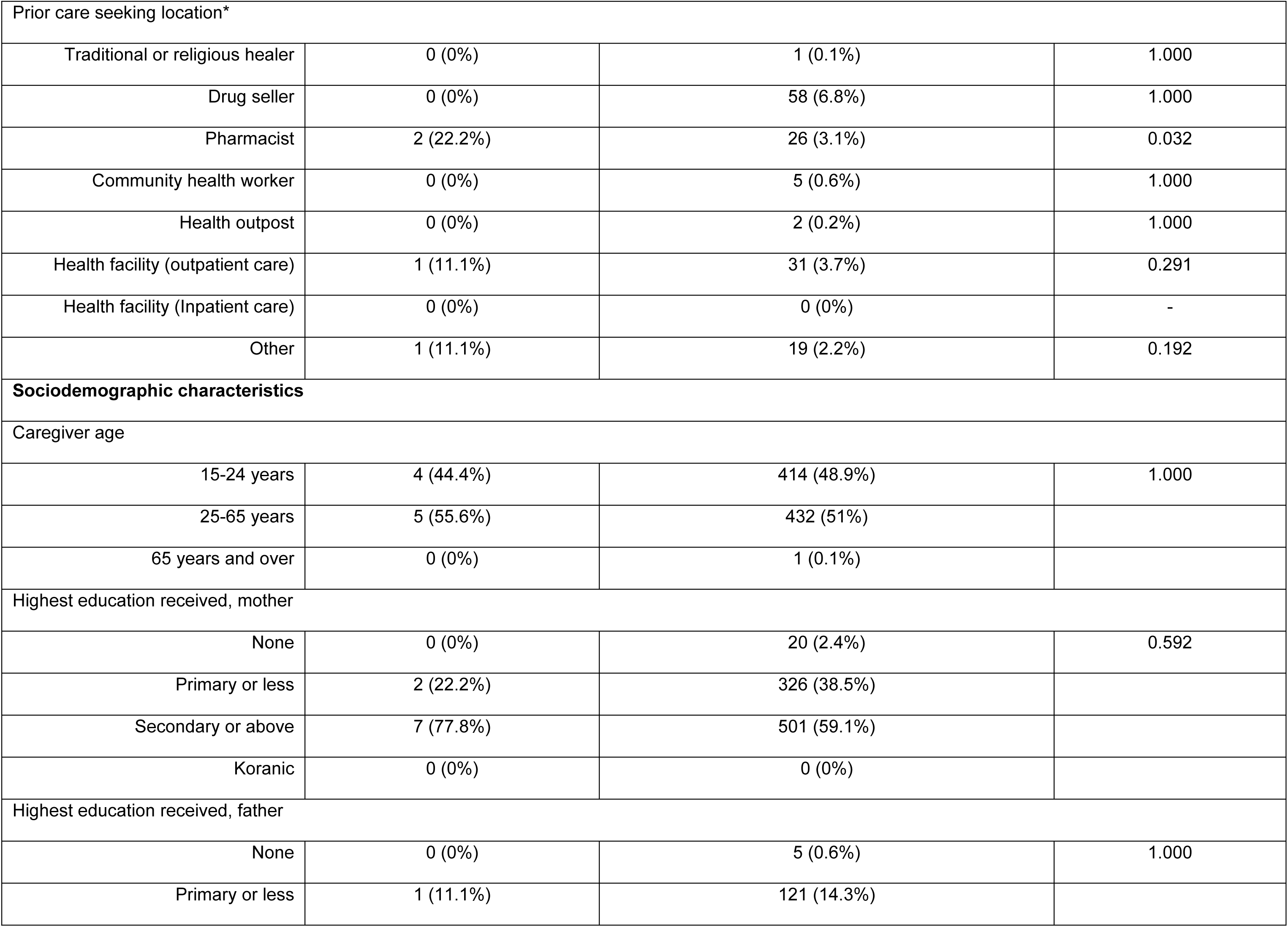
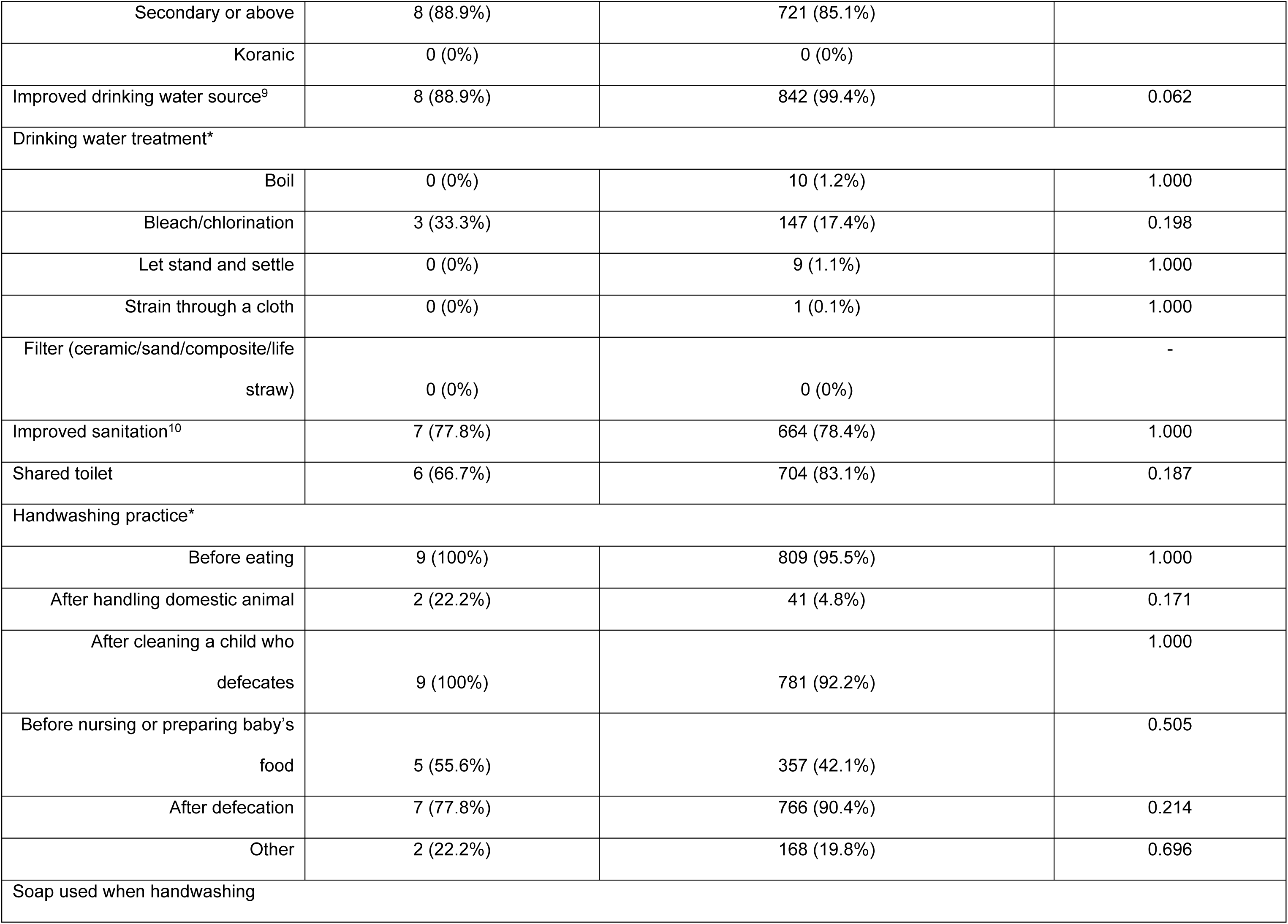
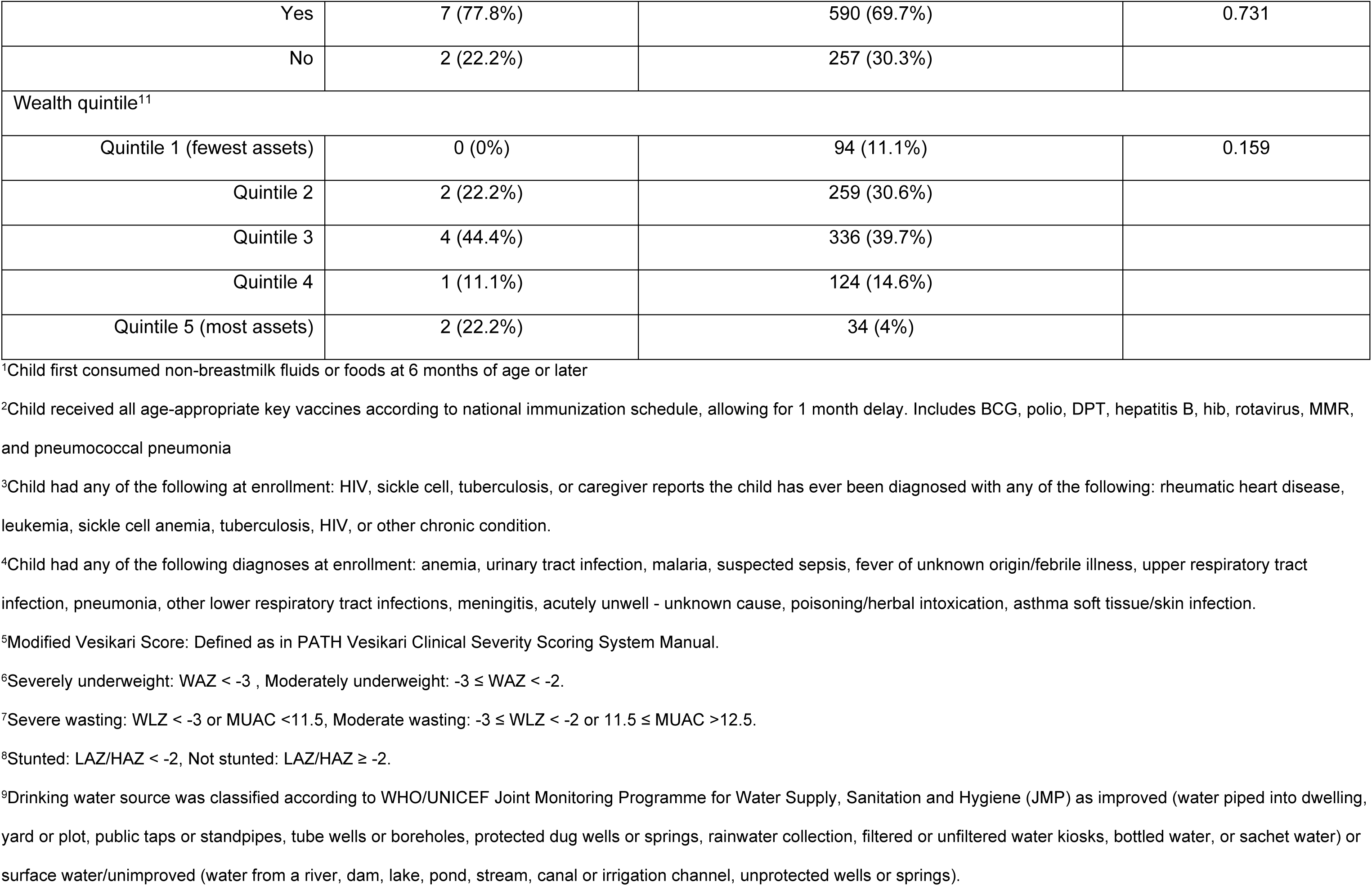
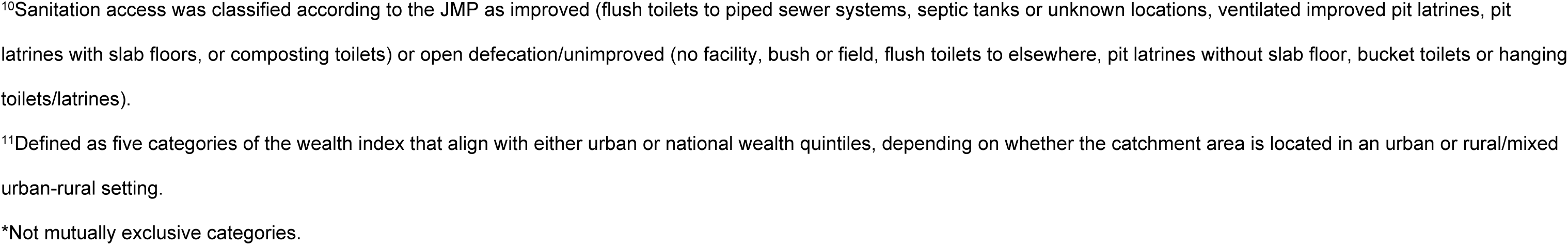
Host, clinical and sociodemographic factors of hospitalization among children aged 6-35 months in Malawi

|  | <u>Hospitalization</u> |  | p-value |
| --- | --- | --- | --- |
|  | Hospitalized (n=9)<br>n (%) | Not hospitalized (n=847)<br>n (%) |  |
| Host factors |  |  |  |
| Male | 5 (55.6%) | 436 (51.5%) | 1.000 |
| Age group |  |  |  |
| 6-11 months | 5 (55.6%) | 359 (42.4%) | 0.532 |
| 12-23 months | 4 (44.4%) | 365 (43.1%) |  |
| 24-35 months | 0 (0%) | 123 (14.5%) |  |
| Exclusively breastfed<sup>1</sup> | 6 (66.7%) | 414 (48.9%) | 0.333 |
| Clinical characteristics |  |  |  |
| Received all age-appropriate vaccines<sup>2</sup> | 2 (22.2%) | 372 (43.9%) | 0.312 |
| Dysentery | 2 (22.2%) | 75 (8.9%) | 0.190 |
| Vomiting | 6 (66.7%) | 405 (47.8%) | 0.325 |
| Fever | 8 (88.9%) | 533 (62.9%) | 0.166 |
| Dehydration |  |  |  |
| None | 8 (88.9%) | 819 (96.7%) | 0.268 |
| Some | 1 (11.1%) | 28 (3.3%) |  |
| Severe | 0 (0%) | 0 (0%) |  |
| Chronic comorbidity<sup>3</sup> | 0 (0%) | 7 (0.8%) | 1.000 |
| Acute comorbidity<sup>4</sup> | 4 (44.4%) | 453 (53.5%) | 0.741 |
| Diarrhea severity (MVS)<sup>5</sup> |  |  |  |
| Mild | 3 (33.3%) | 555 (65.5%) | 0.035 |
| Moderate | 2 (22.2%) | 151 (17.8%) |  |
| Severe | 4 (44.4%) | 141 (16.6%) |  |
| Underweight<sup>6</sup> |  |  |  |
| None | 8 (88.9%) | 754 (89%) | 0.651 |
| Moderate | 1 (11.1%) | 74 (8.7%) |  |
| Severe | 0 (0%) | 19 (2.2%) |  |
| Wasting<sup>7</sup> |  |  |  |
| None | 8 (88.9%) | 806 (95.2%) | 0.366 |
| Moderate | 1 (11.1%) | 35 (4.1%) |  |
| Severe | 0 (0%) | 6 (0.7%) |  |
| Stunting<sup>8</sup> |  |  |  |
| Not stunted | 8 (88.9%) | 643 (75.9%) | 0.695 |
| Stunted | 1 (11.1%) | 204 (24.1%) |  |
| Time taken to seek care |  |  |  |
| <= 1 day | 3 (33.3%) | 435 (51.4%) | 0.331 |
| > 1 day | 6 (66.7%) | 412 (48.6%) |  |
| Prior medical care sought | 4 (44.4%) | 142 (16.8%) | 0.051 |
| Prior care seeking location* |  |  |  |
| Traditional or religious healer | 0 (0%) | 1 (0.1%) | 1.000 |
| Drug seller | 0 (0%) | 58 (6.8%) | 1.000 |
| Pharmacist | 2 (22.2%) | 26 (3.1%) | 0.032 |
| Community health worker | 0 (0%) | 5 (0.6%) | 1.000 |
| Health outpost | 0 (0%) | 2 (0.2%) | 1.000 |
| Health facility (outpatient care) | 1 (11.1%) | 31 (3.7%) | 0.291 |
| Health facility (Inpatient care) | 0 (0%) | 0 (0%) | - |
| Other | 1 (11.1%) | 19 (2.2%) | 0.192 |
| **Sociodemographic characteristics** |  |  |  |
| Caregiver age |  |  |  |
| 15-24 years | 4 (44.4%) | 414 (48.9%) | 1.000 |
| 25-65 years | 5 (55.6%) | 432 (51%) |  |
| 65 years and over | 0 (0%) | 1 (0.1%) |  |
| Highest education received, mother |  |  |  |
| None | 0 (0%) | 20 (2.4%) | 0.592 |
| Primary or less | 2 (22.2%) | 326 (38.5%) |  |
| Secondary or above | 7 (77.8%) | 501 (59.1%) |  |
| Koranic | 0 (0%) | 0 (0%) |  |
| Highest education received, father |  |  |  |
| None | 0 (0%) | 5 (0.6%) | 1.000 |
| Primary or less | 1 (11.1%) | 121 (14.3%) |  |
| Secondary or above | 8 (88.9%) | 721 (85.1%) |  |
| Koranic | 0 (0%) | 0 (0%) |  |
| Improved drinking water source<sup>9</sup> | 8 (88.9%) | 842 (99.4%) | 0.062 |
| Drinking water treatment* |  |  |  |
| Boil | 0 (0%) | 10 (1.2%) | 1.000 |
| Bleach/chlorination | 3 (33.3%) | 147 (17.4%) | 0.198 |
| Let stand and settle | 0 (0%) | 9 (1.1%) | 1.000 |
| Strain through a cloth | 0 (0%) | 1 (0.1%) | 1.000 |
| Filter (ceramic/sand/composite/life straw) | 0 (0%) | 0 (0%) | - |
| Improved sanitation<sup>10</sup> | 7 (77.8%) | 664 (78.4%) | 1.000 |
| Shared toilet | 6 (66.7%) | 704 (83.1%) | 0.187 |
| Handwashing practice* |  |  |  |
| Before eating | 9 (100%) | 809 (95.5%) | 1.000 |
| After handling domestic animal | 2 (22.2%) | 41 (4.8%) | 0.171 |
| After cleaning a child who defecates | 9 (100%) | 781 (92.2%) | 1.000 |
| Before nursing or preparing baby's food | 5 (55.6%) | 357 (42.1%) | 0.505 |
| After defecation | 7 (77.8%) | 766 (90.4%) | 0.214 |
| Other | 2 (22.2%) | 168 (19.8%) | 0.696 |
| Soap used when handwashing |  |  |  |
| Yes | 7 (77.8%) | 590 (69.7%) | 0.731 |
| No | 2 (22.2%) | 257 (30.3%) |  |
| Wealth quintile<sup>11</sup> |  |  |  |
| Quintile 1 (fewest assets) | 0 (0%) | 94 (11.1%) | 0.159 |
| Quintile 2 | 2 (22.2%) | 259 (30.6%) |  |
| Quintile 3 | 4 (44.4%) | 336 (39.7%) |  |
| Quintile 4 | 1 (11.1%) | 124 (14.6%) |  |
| Quintile 5 (most assets) | 2 (22.2%) | 34 (4%) |  |

<sup>1</sup>Child first consumed non-breastmilk fluids or foods at 6 months of age or later
<sup>2</sup>Child received all age-appropriate key vaccines according to national immunization schedule, allowing for 1 month delay. Includes BCG, polio, DPT, hepatitis B, hib, rotavirus, MMR, and pneumococcal pneumonia
<sup>3</sup>Child had any of the following at enrollment: HIV, sickle cell, tuberculosis, or caregiver reports the child has ever been diagnosed with any of the following: rheumatic heart disease, leukemia, sickle cell anemia, tuberculosis, HIV, or other chronic condition.
<sup>4</sup>Child had any of the following diagnoses at enrollment: anemia, urinary tract infection, malaria, suspected sepsis, fever of unknown origin/febrile illness, upper respiratory tract infection, pneumonia, other lower respiratory tract infections, meningitis, acutely unwell - unknown cause, poisoning/herbal intoxication, asthma soft tissue/skin infection.
<sup>5</sup>Modified Vesikari Score: Defined as in PATH Vesikari Clinical Severity Scoring System Manual.
<sup>6</sup>Severely underweight: WAZ < -3 , Moderately underweight: -3 ≤ WAZ < -2.
<sup>7</sup>Severe wasting: WLZ < -3 or MUAC <11.5, Moderate wasting: -3 ≤ WLZ < -2 or 11.5 ≤ MUAC >12.5.
<sup>8</sup>Stunted: LAZ/HAZ < -2, Not stunted: LAZ/HAZ ≥ -2.
<sup>9</sup>Drinking water source was classified according to WHO/UNICEF Joint Monitoring Programme for Water Supply, Sanitation and Hygiene (JMP) as improved (water piped into dwelling, yard or plot, public taps or standpipes, tube wells or boreholes, protected dug wells or springs, rainwater collection, filtered or unfiltered water kiosks, bottled water, or sachet water) or surface water/unimproved (water from a river, dam, lake, pond, stream, canal or irrigation channel, unprotected wells or springs).
<sup>10</sup>Sanitation access was classified according to the JMP as improved (flush toilets to piped sewer systems, septic tanks or unknown locations, ventilated improved pit latrines, pit latrines with slab floors, or composting toilets) or open defecation/unimproved (no facility, bush or field, flush toilets to elsewhere, pit latrines without slab floor, bucket toilets or hanging toilets/latrines). <sup>11</sup>Defined as five categories of the wealth index that align with either urban or national wealth quintiles, depending on whether the catchment area is located in an urban or rural/mixed urban-rural setting. \*Not mutually exclusive categories.

**S4 Table.**
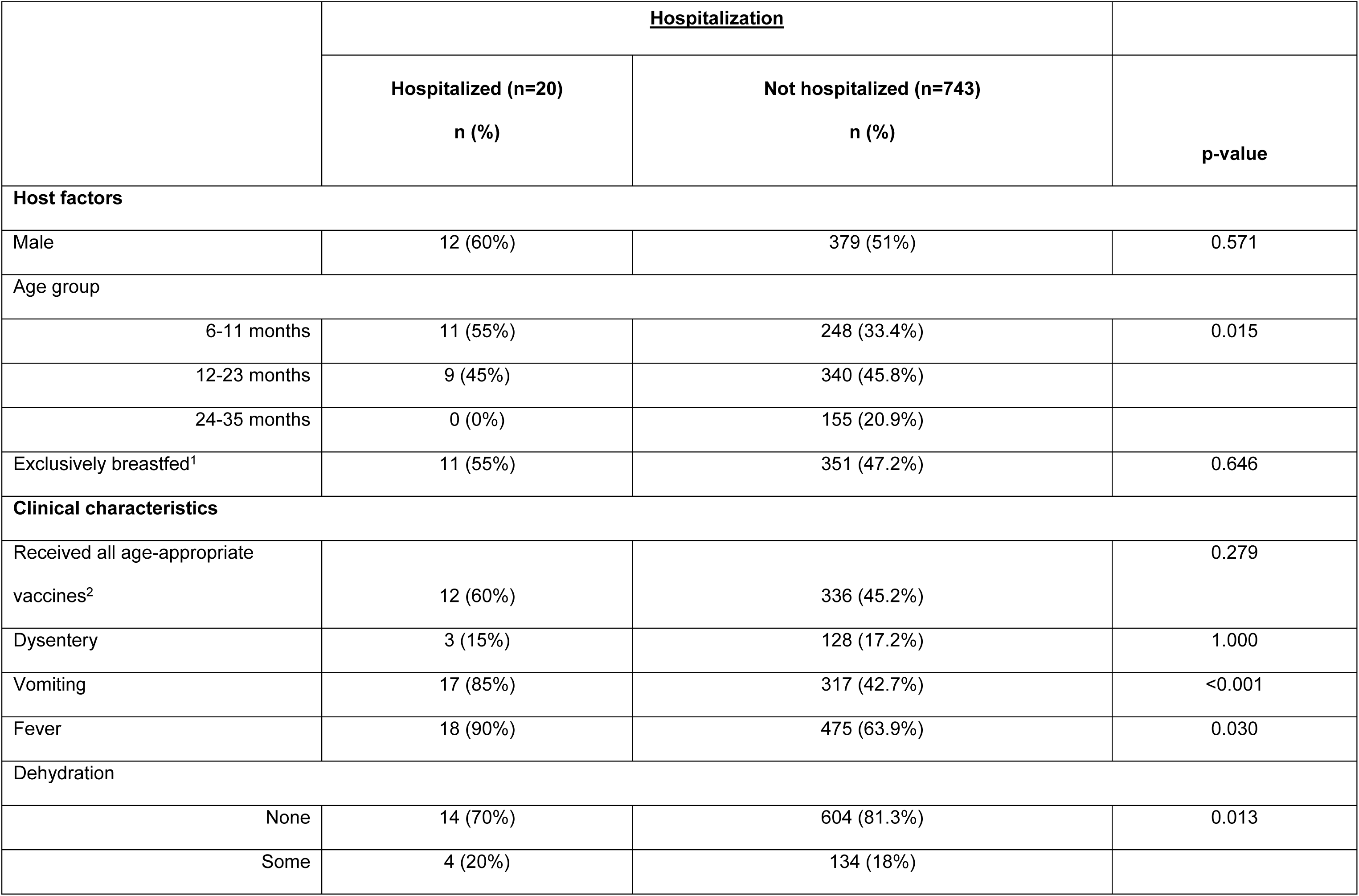
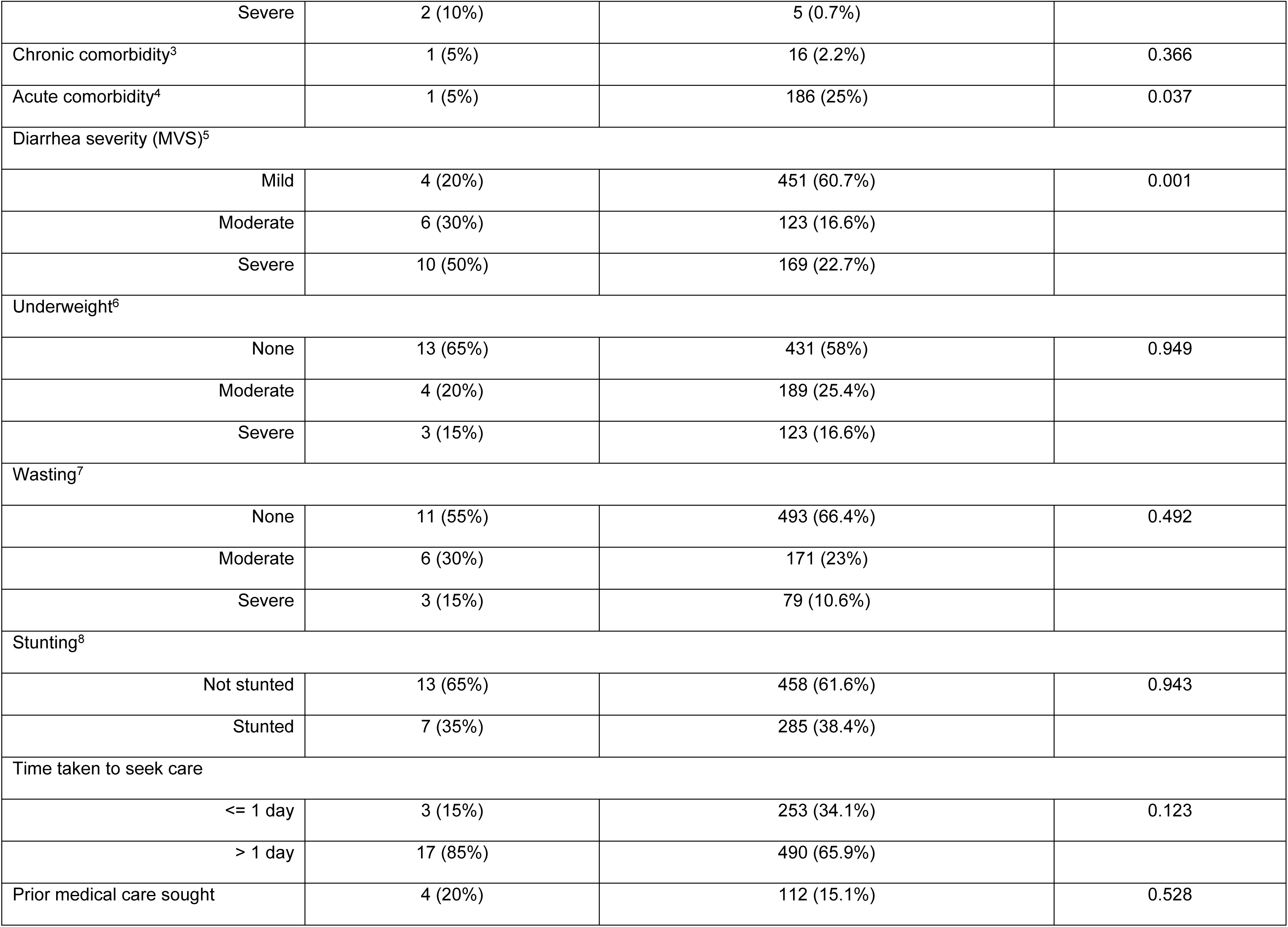
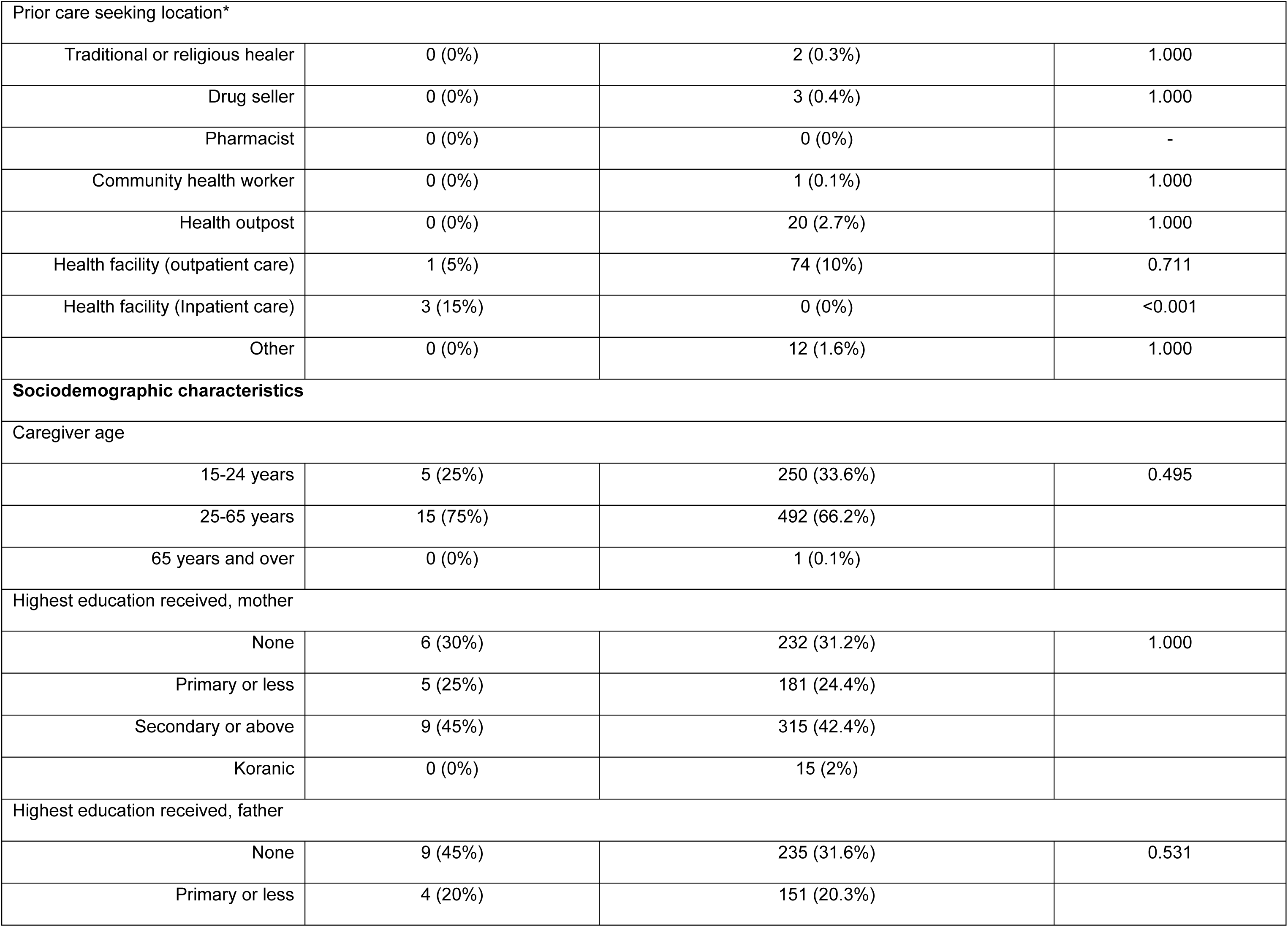
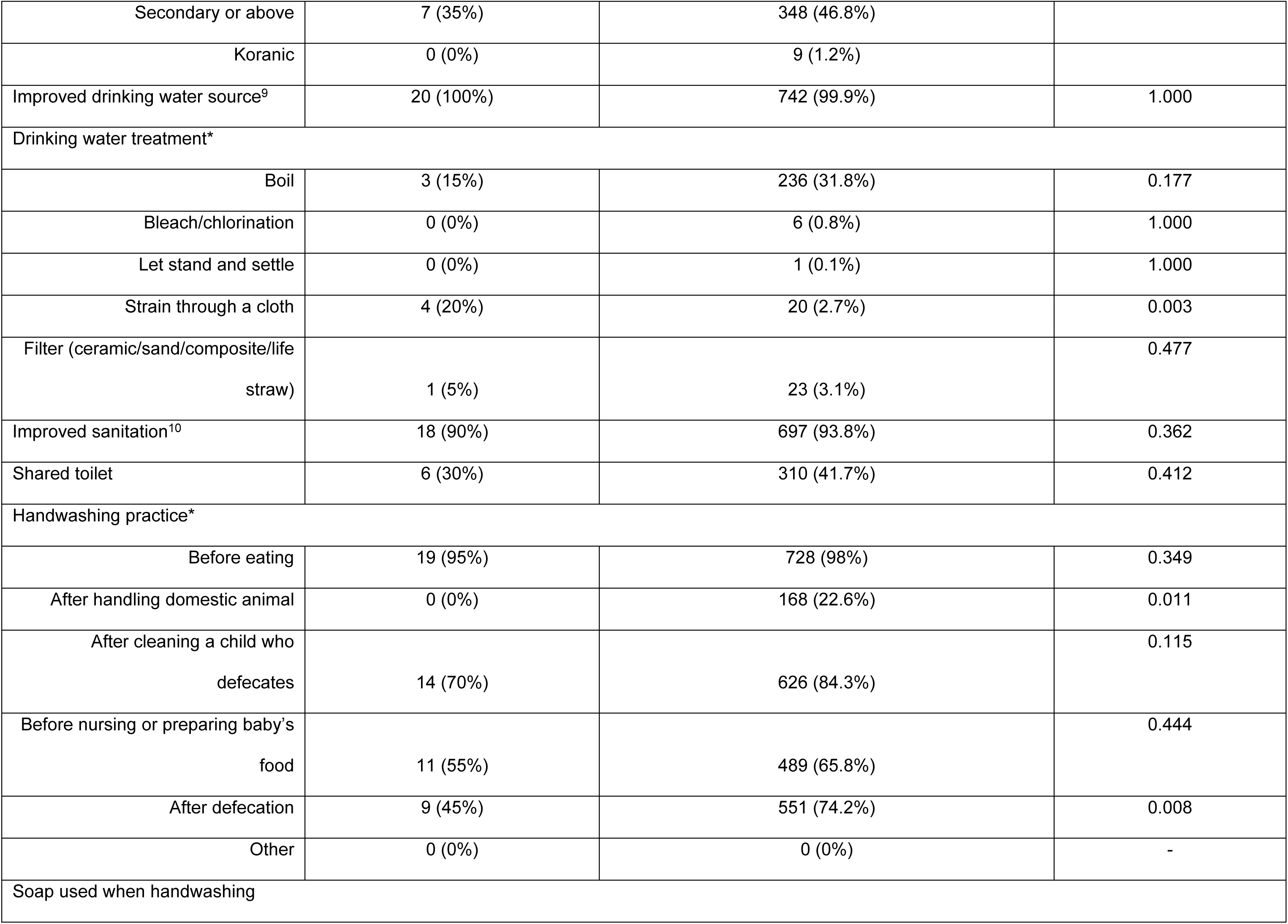
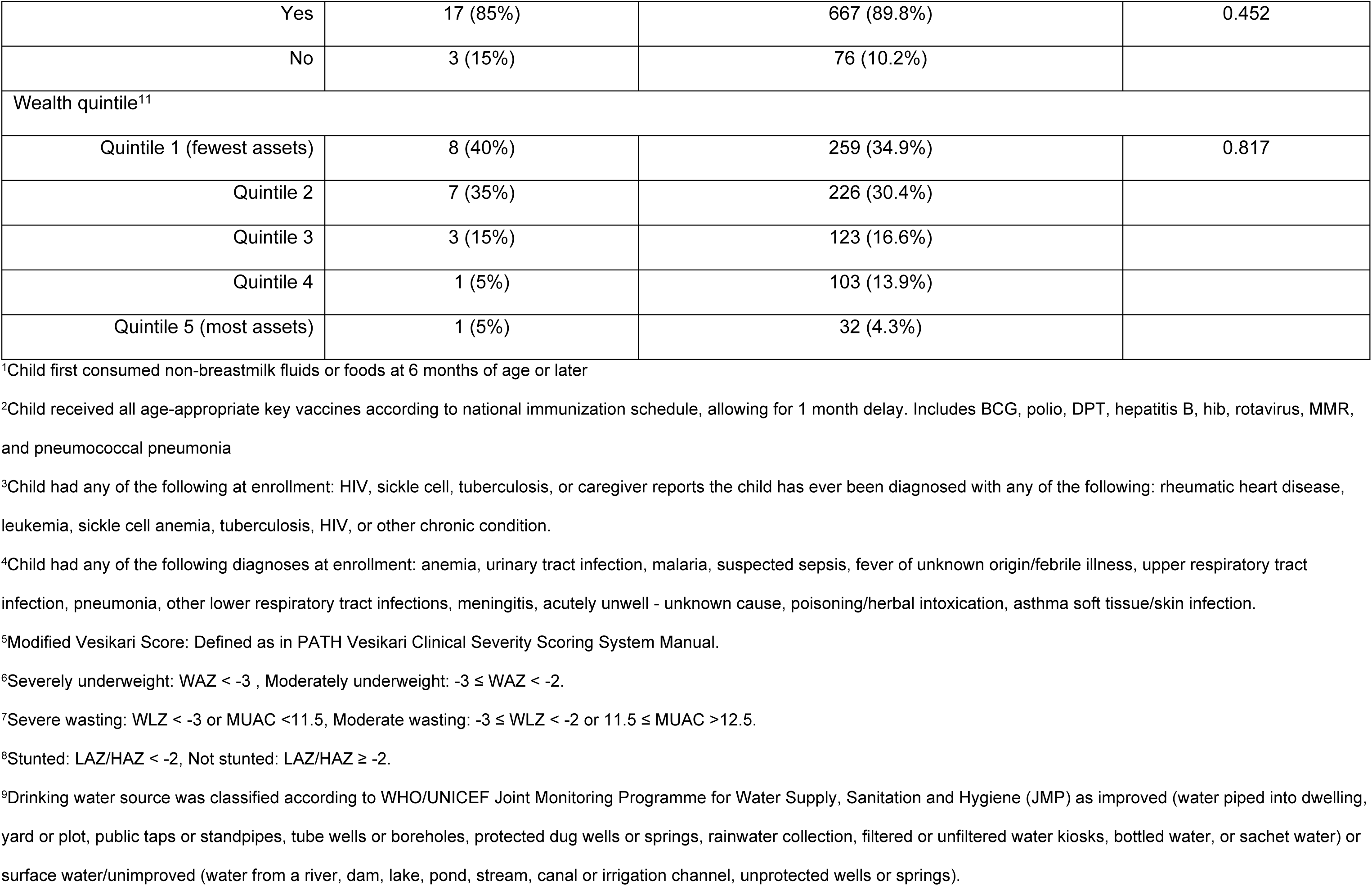
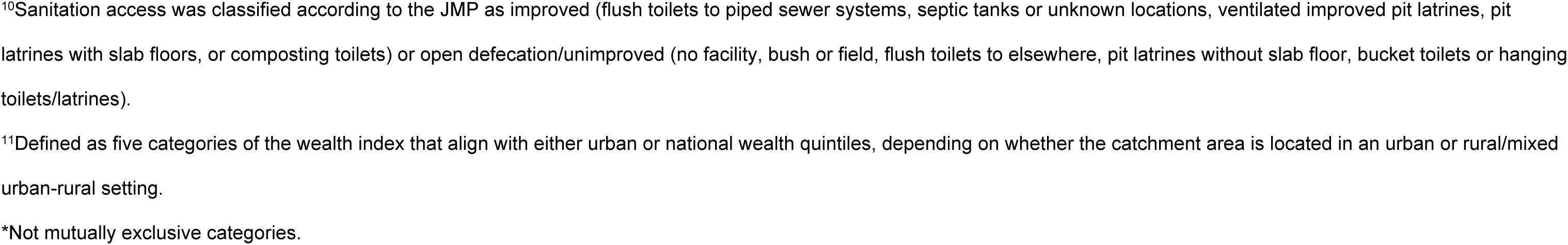
Host, clinical and sociodemographic factors of hospitalization among children aged 6-35 months in Pakistan

|  | <u>Hospitalization</u> |  | p-value |
| --- | --- | --- | --- |
|  | Hospitalized (n=20)<br>n (%) | Not hospitalized (n=743)<br>n (%) |  |
| Host factors |  |  |  |
| Male | 12 (60%) | 379 (51%) | 0.571 |
| Age group |  |  |  |
| 6-11 months | 11 (55%) | 248 (33.4%) | 0.015 |
| 12-23 months | 9 (45%) | 340 (45.8%) |  |
| 24-35 months | 0 (0%) | 155 (20.9%) |  |
| Exclusively breastfed<sup>1</sup> | 11 (55%) | 351 (47.2%) | 0.646 |
| Clinical characteristics |  |  |  |
| Received all age-appropriate vaccines<sup>2</sup> | 12 (60%) | 336 (45.2%) | 0.279 |
| Dysentery | 3 (15%) | 128 (17.2%) | 1.000 |
| Vomiting | 17 (85%) | 317 (42.7%) | <0.001 |
| Fever | 18 (90%) | 475 (63.9%) | 0.030 |
| Dehydration |  |  |  |
| None | 14 (70%) | 604 (81.3%) | 0.013 |
| Some | 4 (20%) | 134 (18%) |  |
| Severe | 2 (10%) | 5 (0.7%) |  |
| Chronic comorbidity<sup>3</sup> | 1 (5%) | 16 (2.2%) | 0.366 |
| Acute comorbidity<sup>4</sup> | 1 (5%) | 186 (25%) | 0.037 |
| Diarrhea severity (MVS)<sup>5</sup> |  |  |  |
| Mild | 4 (20%) | 451 (60.7%) | 0.001 |
| Moderate | 6 (30%) | 123 (16.6%) |  |
| Severe | 10 (50%) | 169 (22.7%) |  |
| Underweight<sup>6</sup> |  |  |  |
| None | 13 (65%) | 431 (58%) | 0.949 |
| Moderate | 4 (20%) | 189 (25.4%) |  |
| Severe | 3 (15%) | 123 (16.6%) |  |
| Wasting<sup>7</sup> |  |  |  |
| None | 11 (55%) | 493 (66.4%) | 0.492 |
| Moderate | 6 (30%) | 171 (23%) |  |
| Severe | 3 (15%) | 79 (10.6%) |  |
| Stunting<sup>8</sup> |  |  |  |
| Not stunted | 13 (65%) | 458 (61.6%) | 0.943 |
| Stunted | 7 (35%) | 285 (38.4%) |  |
| Time taken to seek care |  |  |  |
| <= 1 day | 3 (15%) | 253 (34.1%) | 0.123 |
| > 1 day | 17 (85%) | 490 (65.9%) |  |
| Prior medical care sought | 4 (20%) | 112 (15.1%) | 0.528 |
| Prior care seeking location* |  |  |  |
| Traditional or religious healer | 0 (0%) | 2 (0.3%) | 1.000 |
| Drug seller | 0 (0%) | 3 (0.4%) | 1.000 |
| Pharmacist | 0 (0%) | 0 (0%) | - |
| Community health worker | 0 (0%) | 1 (0.1%) | 1.000 |
| Health outpost | 0 (0%) | 20 (2.7%) | 1.000 |
| Health facility (outpatient care) | 1 (5%) | 74 (10%) | 0.711 |
| Health facility (Inpatient care) | 3 (15%) | 0 (0%) | <0.001 |
| Other | 0 (0%) | 12 (1.6%) | 1.000 |
| **Sociodemographic characteristics** |  |  |  |
| Caregiver age |  |  |  |
| 15-24 years | 5 (25%) | 250 (33.6%) | 0.495 |
| 25-65 years | 15 (75%) | 492 (66.2%) |  |
| 65 years and over | 0 (0%) | 1 (0.1%) |  |
| Highest education received, mother |  |  |  |
| None | 6 (30%) | 232 (31.2%) | 1.000 |
| Primary or less | 5 (25%) | 181 (24.4%) |  |
| Secondary or above | 9 (45%) | 315 (42.4%) |  |
| Koranic | 0 (0%) | 15 (2%) |  |
| Highest education received, father |  |  |  |
| None | 9 (45%) | 235 (31.6%) | 0.531 |
| Primary or less | 4 (20%) | 151 (20.3%) |  |
| Secondary or above | 7 (35%) | 348 (46.8%) |  |
| Koranic | 0 (0%) | 9 (1.2%) |  |
| Improved drinking water source<sup>9</sup> | 20 (100%) | 742 (99.9%) | 1.000 |
| Drinking water treatment* |  |  |  |
| Boil | 3 (15%) | 236 (31.8%) | 0.177 |
| Bleach/chlorination | 0 (0%) | 6 (0.8%) | 1.000 |
| Let stand and settle | 0 (0%) | 1 (0.1%) | 1.000 |
| Strain through a cloth | 4 (20%) | 20 (2.7%) | 0.003 |
| Filter (ceramic/sand/composite/life straw) | 1 (5%) | 23 (3.1%) | 0.477 |
| Improved sanitation<sup>10</sup> | 18 (90%) | 697 (93.8%) | 0.362 |
| Shared toilet | 6 (30%) | 310 (41.7%) | 0.412 |
| Handwashing practice* |  |  |  |
| Before eating | 19 (95%) | 728 (98%) | 0.349 |
| After handling domestic animal | 0 (0%) | 168 (22.6%) | 0.011 |
| After cleaning a child who defecates | 14 (70%) | 626 (84.3%) | 0.115 |
| Before nursing or preparing baby's food | 11 (55%) | 489 (65.8%) | 0.444 |
| After defecation | 9 (45%) | 551 (74.2%) | 0.008 |
| Other | 0 (0%) | 0 (0%) | - |
| Soap used when handwashing |  |  |  |
| Yes | 17 (85%) | 667 (89.8%) | 0.452 |
| No | 3 (15%) | 76 (10.2%) |  |
| Wealth quintile<sup>11</sup> |  |  |  |
| Quintile 1 (fewest assets) | 8 (40%) | 259 (34.9%) | 0.817 |
| Quintile 2 | 7 (35%) | 226 (30.4%) |  |
| Quintile 3 | 3 (15%) | 123 (16.6%) |  |
| Quintile 4 | 1 (5%) | 103 (13.9%) |  |
| Quintile 5 (most assets) | 1 (5%) | 32 (4.3%) |  |

<sup>1</sup>Child first consumed non-breastmilk fluids or foods at 6 months of age or later
<sup>2</sup>Child received all age-appropriate key vaccines according to national immunization schedule, allowing for 1 month delay. Includes BCG, polio, DPT, hepatitis B, hib, rotavirus, MMR, and pneumococcal pneumonia
<sup>3</sup>Child had any of the following at enrollment: HIV, sickle cell, tuberculosis, or caregiver reports the child has ever been diagnosed with any of the following: rheumatic heart disease, leukemia, sickle cell anemia, tuberculosis, HIV, or other chronic condition.
<sup>4</sup>Child had any of the following diagnoses at enrollment: anemia, urinary tract infection, malaria, suspected sepsis, fever of unknown origin/febrile illness, upper respiratory tract infection, pneumonia, other lower respiratory tract infections, meningitis, acutely unwell - unknown cause, poisoning/herbal intoxication, asthma soft tissue/skin infection.
<sup>5</sup>Modified Vesikari Score: Defined as in PATH Vesikari Clinical Severity Scoring System Manual.
<sup>6</sup>Severely underweight: WAZ < -3 , Moderately underweight: -3 ≤ WAZ < -2.
<sup>7</sup>Severe wasting: WLZ < -3 or MUAC <11.5, Moderate wasting: -3 ≤ WLZ < -2 or 11.5 ≤ MUAC >12.5.
<sup>8</sup>Stunted: LAZ/HAZ < -2, Not stunted: LAZ/HAZ ≥ -2.
<sup>9</sup>Drinking water source was classified according to WHO/UNICEF Joint Monitoring Programme for Water Supply, Sanitation and Hygiene (JMP) as improved (water piped into dwelling, yard or plot, public taps or standpipes, tube wells or boreholes, protected dug wells or springs, rainwater collection, filtered or unfiltered water kiosks, bottled water, or sachet water) or surface water/unimproved (water from a river, dam, lake, pond, stream, canal or irrigation channel, unprotected wells or springs).
<sup>10</sup>Sanitation access was classified according to the JMP as improved (flush toilets to piped sewer systems, septic tanks or unknown locations, ventilated improved pit latrines, pit latrines with slab floors, or composting toilets) or open defecation/unimproved (no facility, bush or field, flush toilets to elsewhere, pit latrines without slab floor, bucket toilets or hanging toilets/latrines). <sup>11</sup>Defined as five categories of the wealth index that align with either urban or national wealth quintiles, depending on whether the catchment area is located in an urban or rural/mixed urban-rural setting. \*Not mutually exclusive categories.

**S5 Table.**
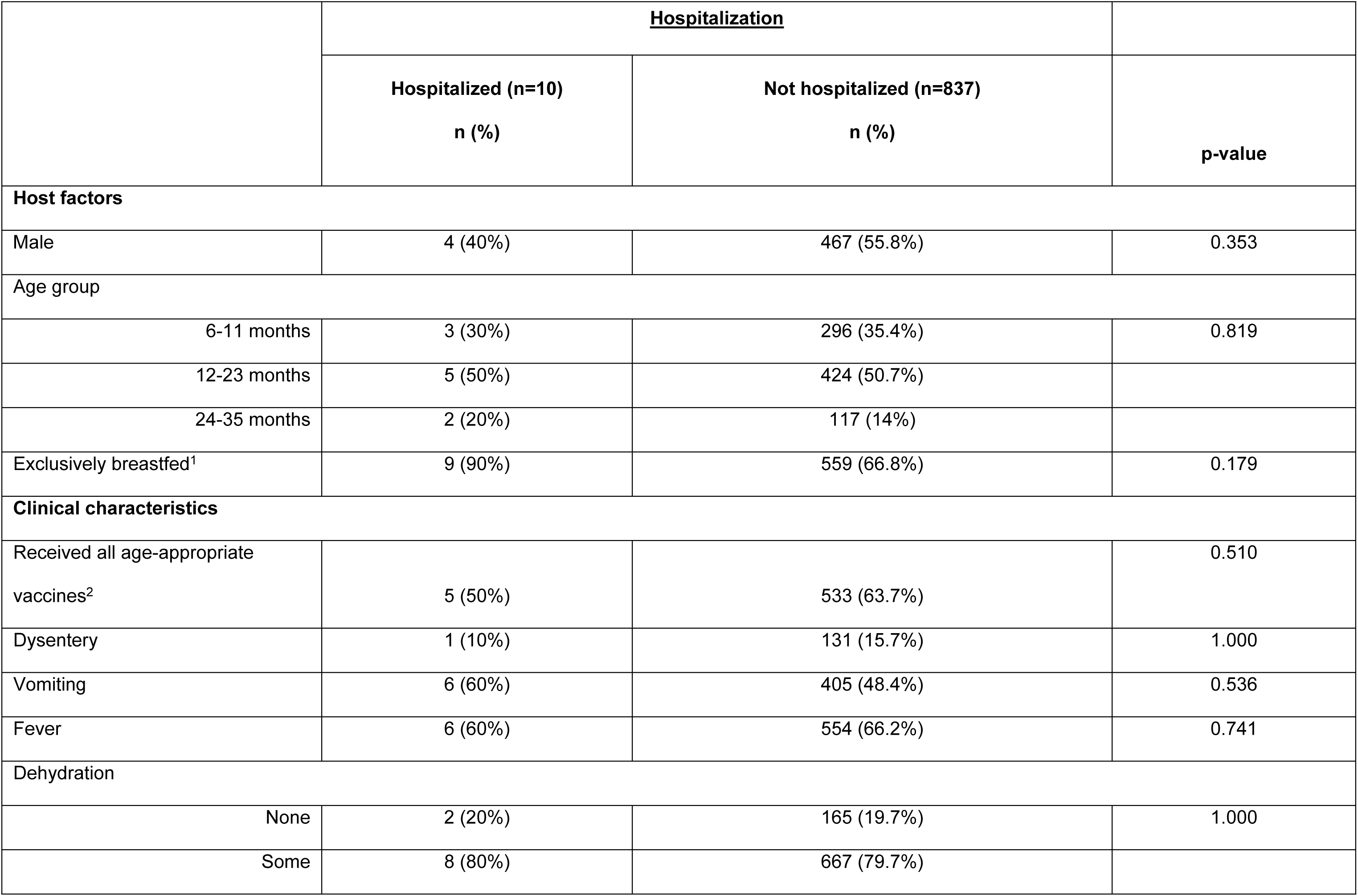
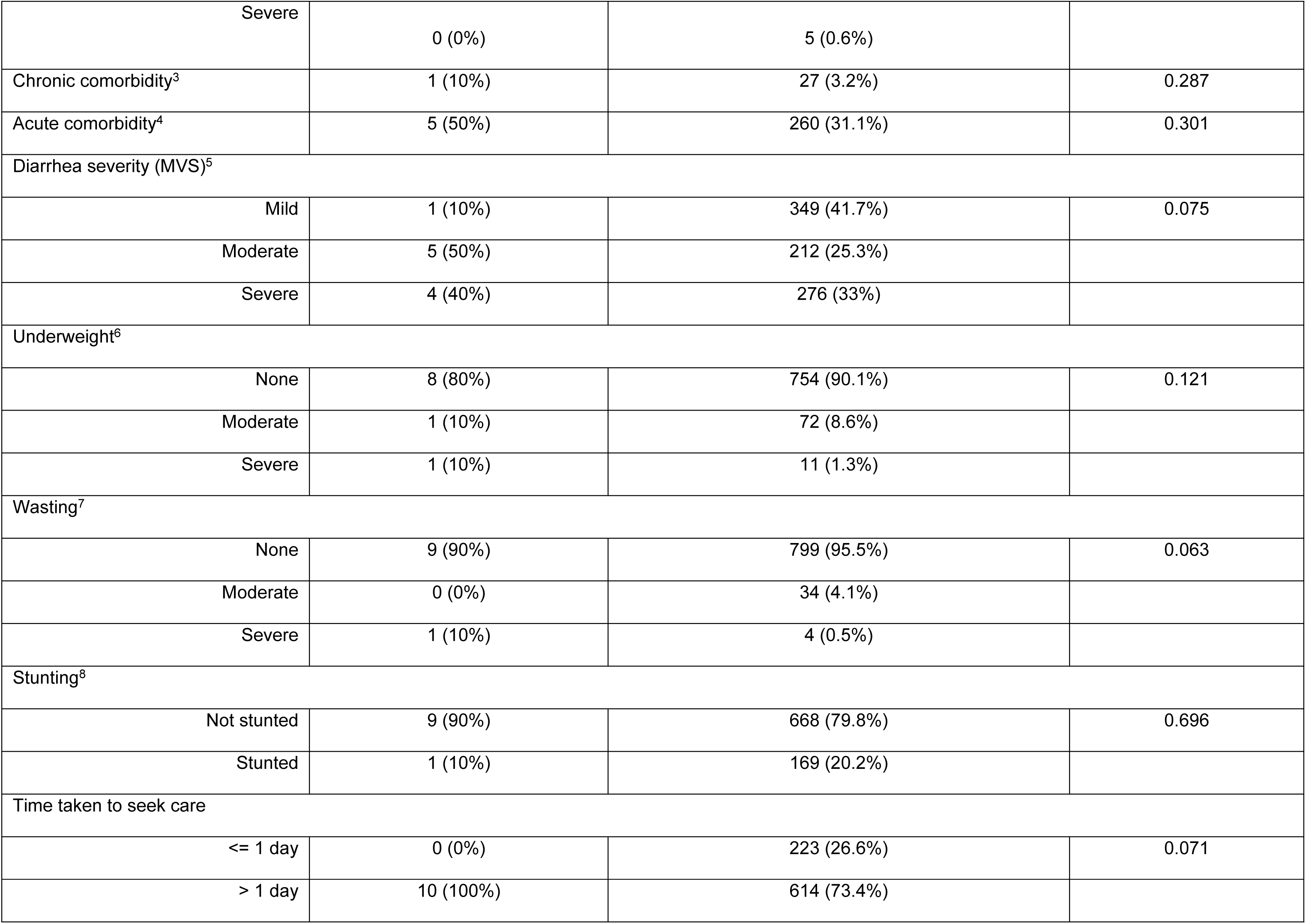
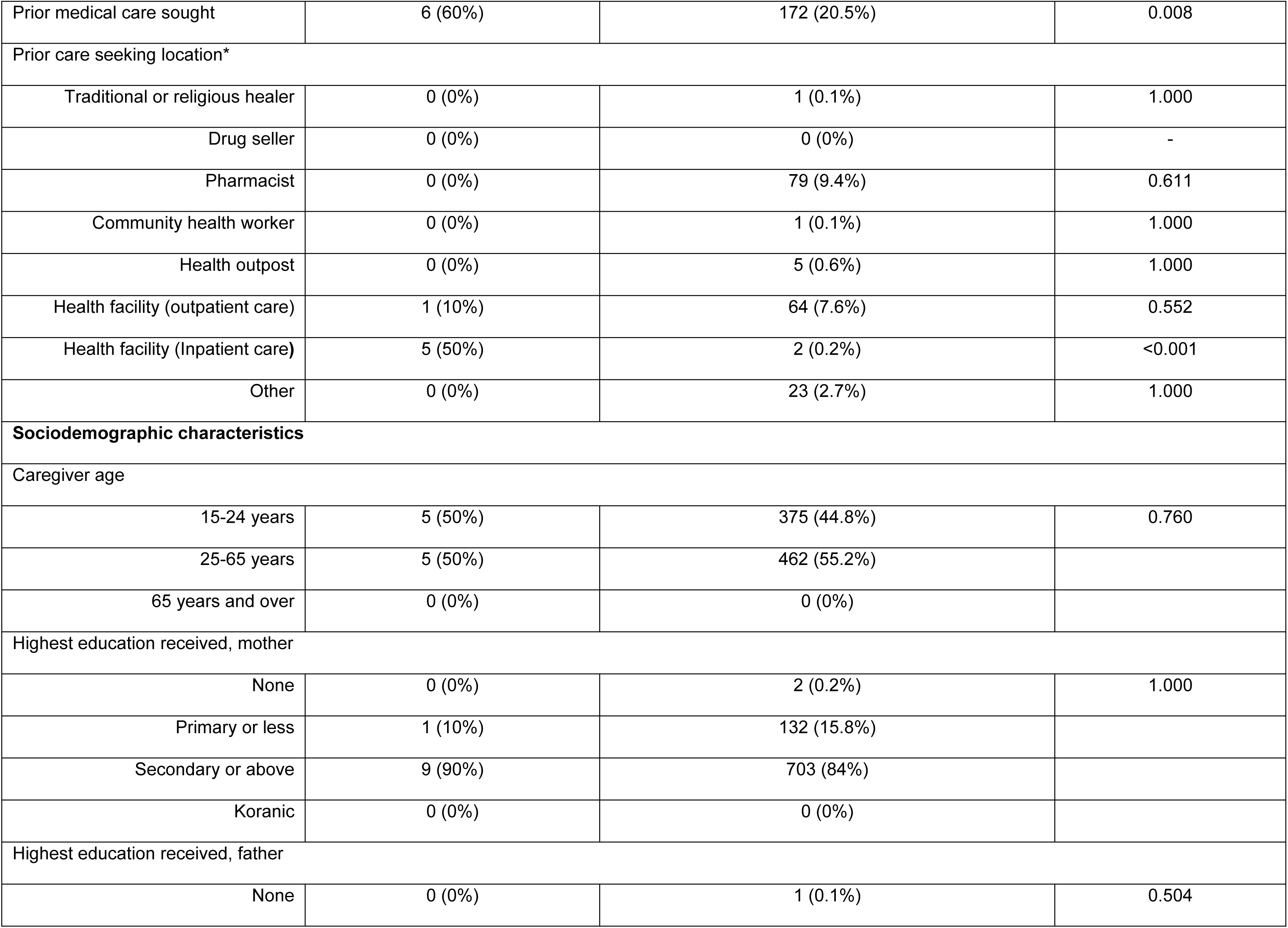
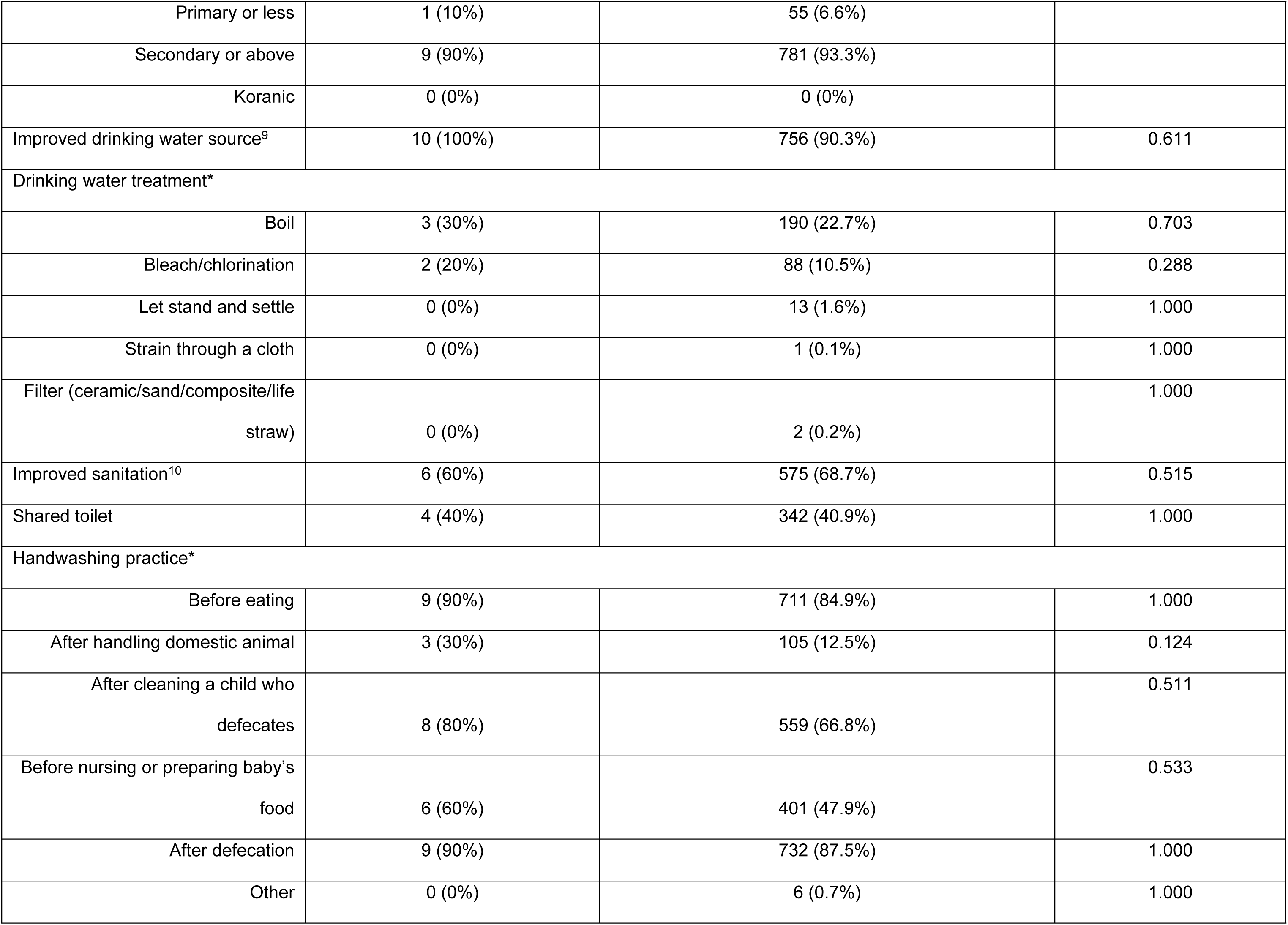
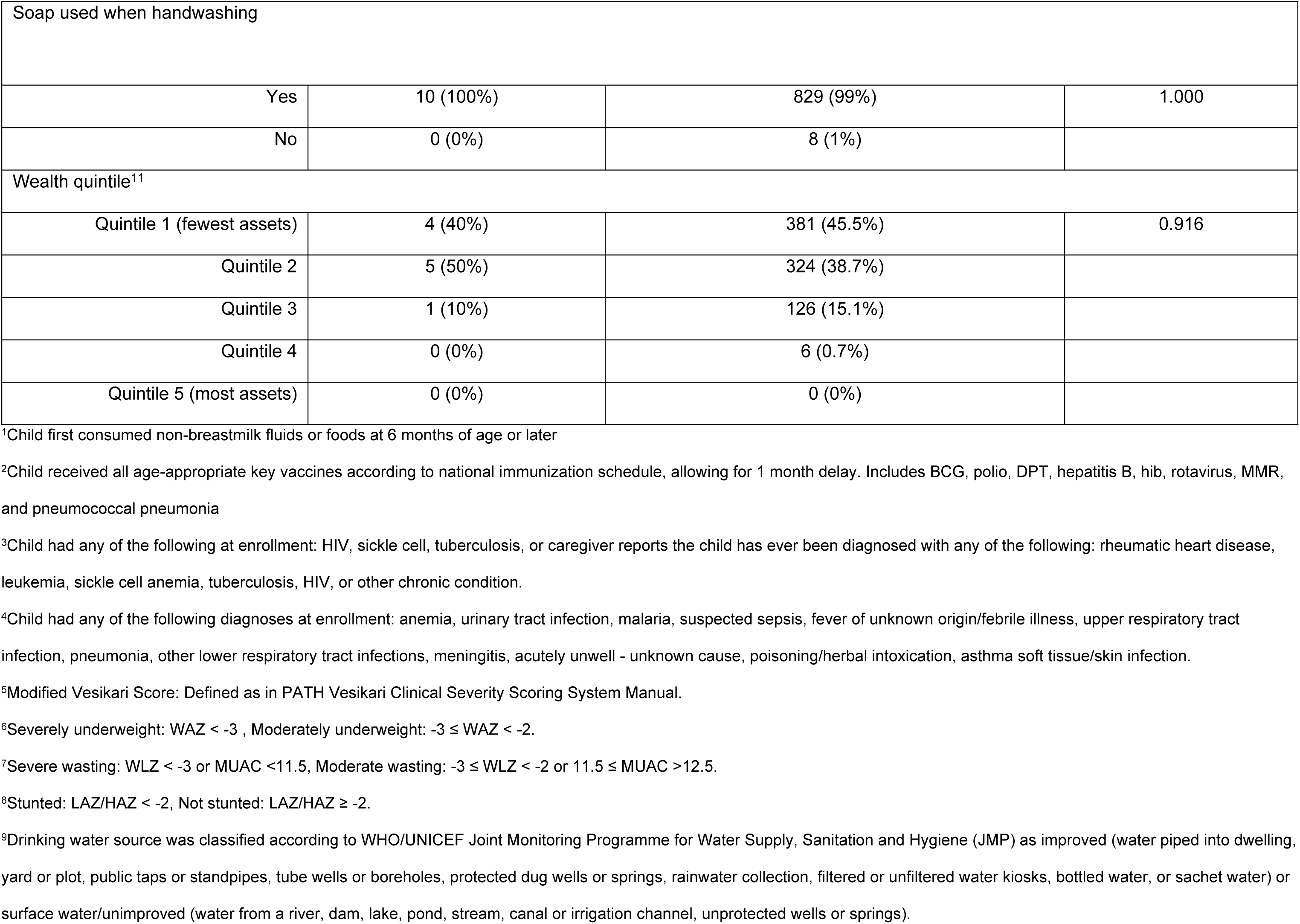
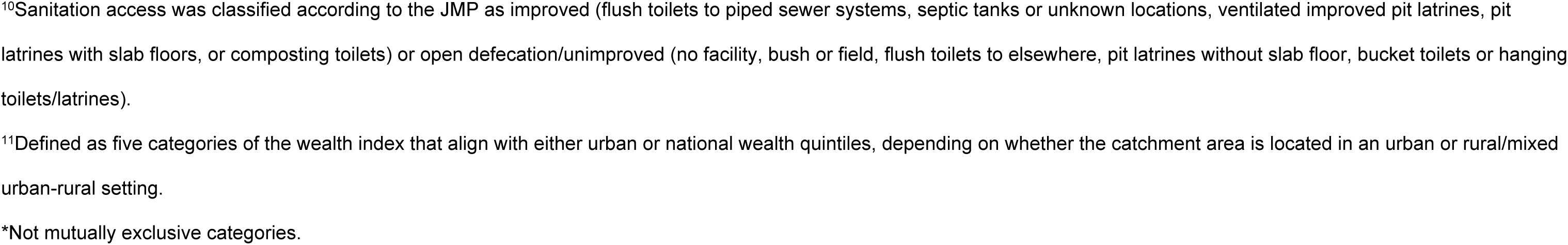
Host, clinical and sociodemographic factors of hospitalization among children aged 6-35 months in Peru

**S6 Table.**
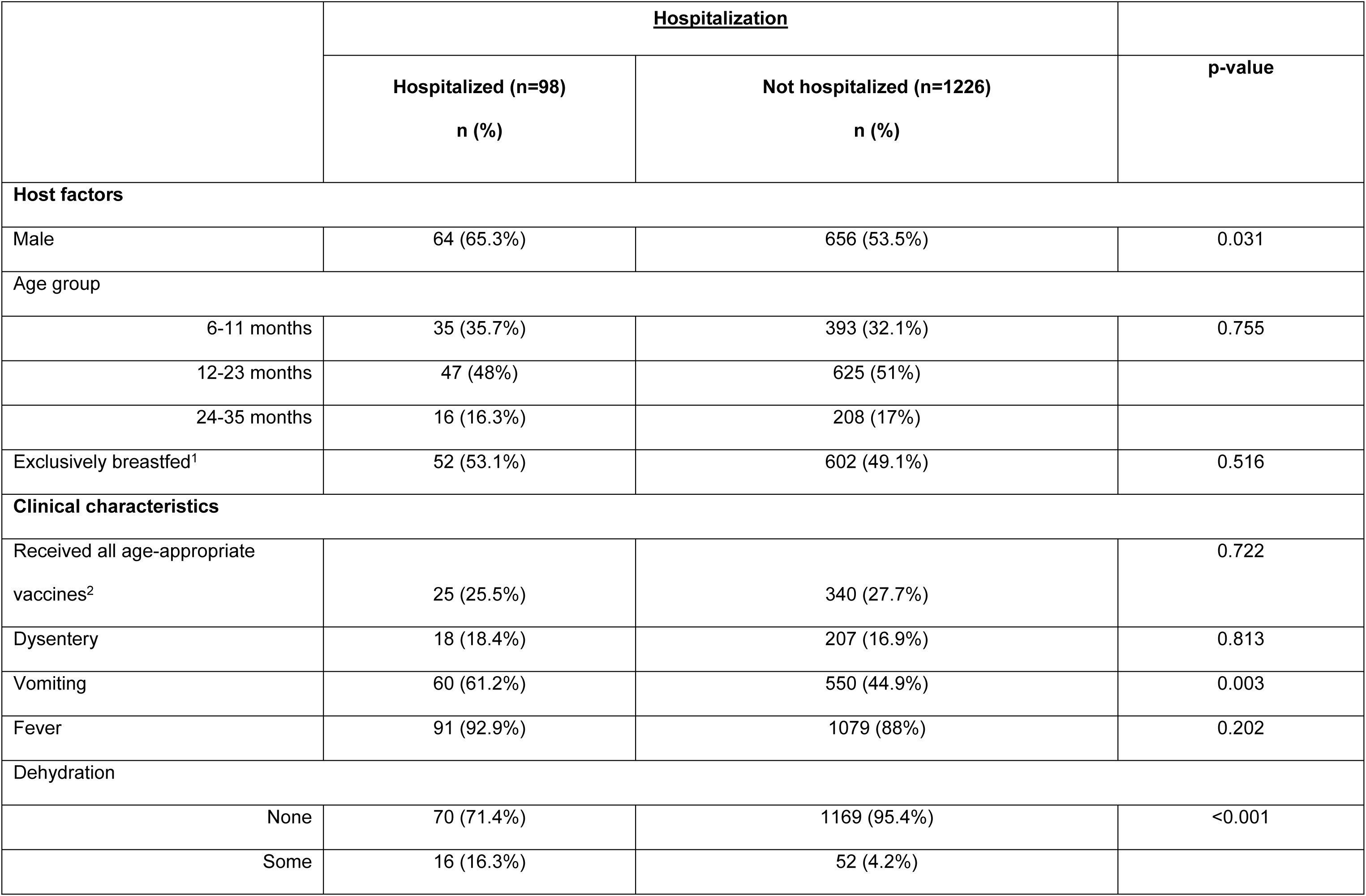
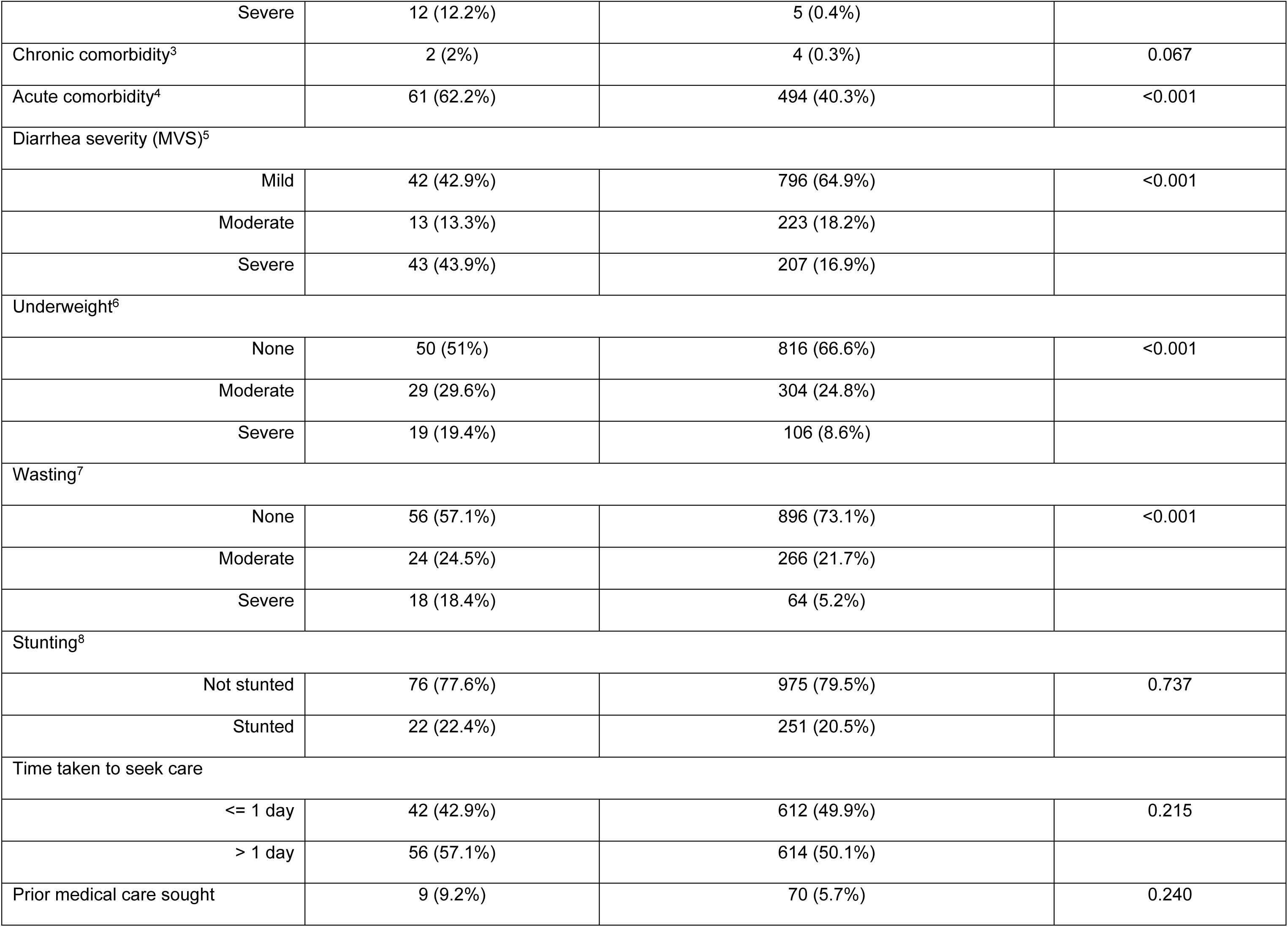
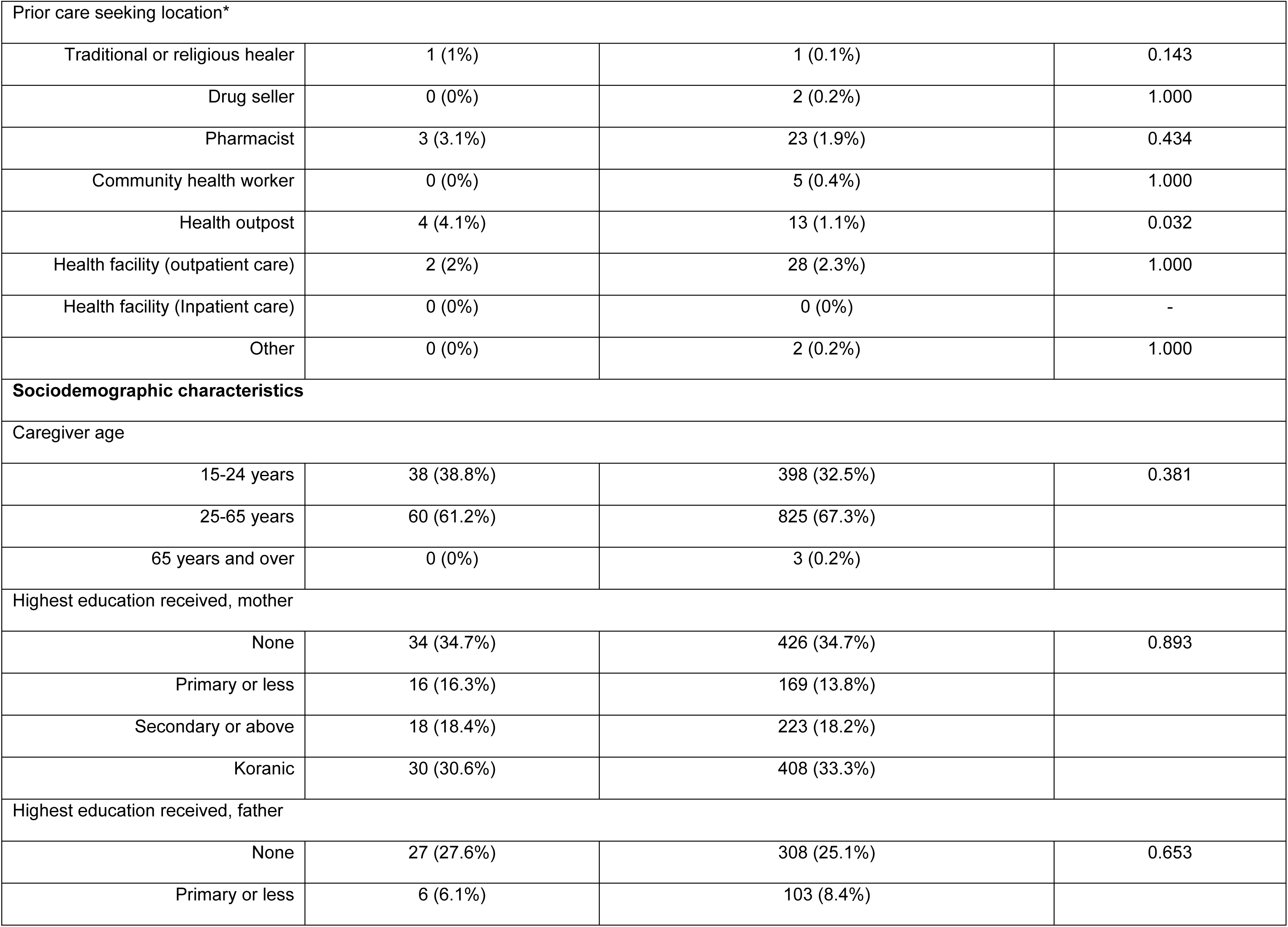
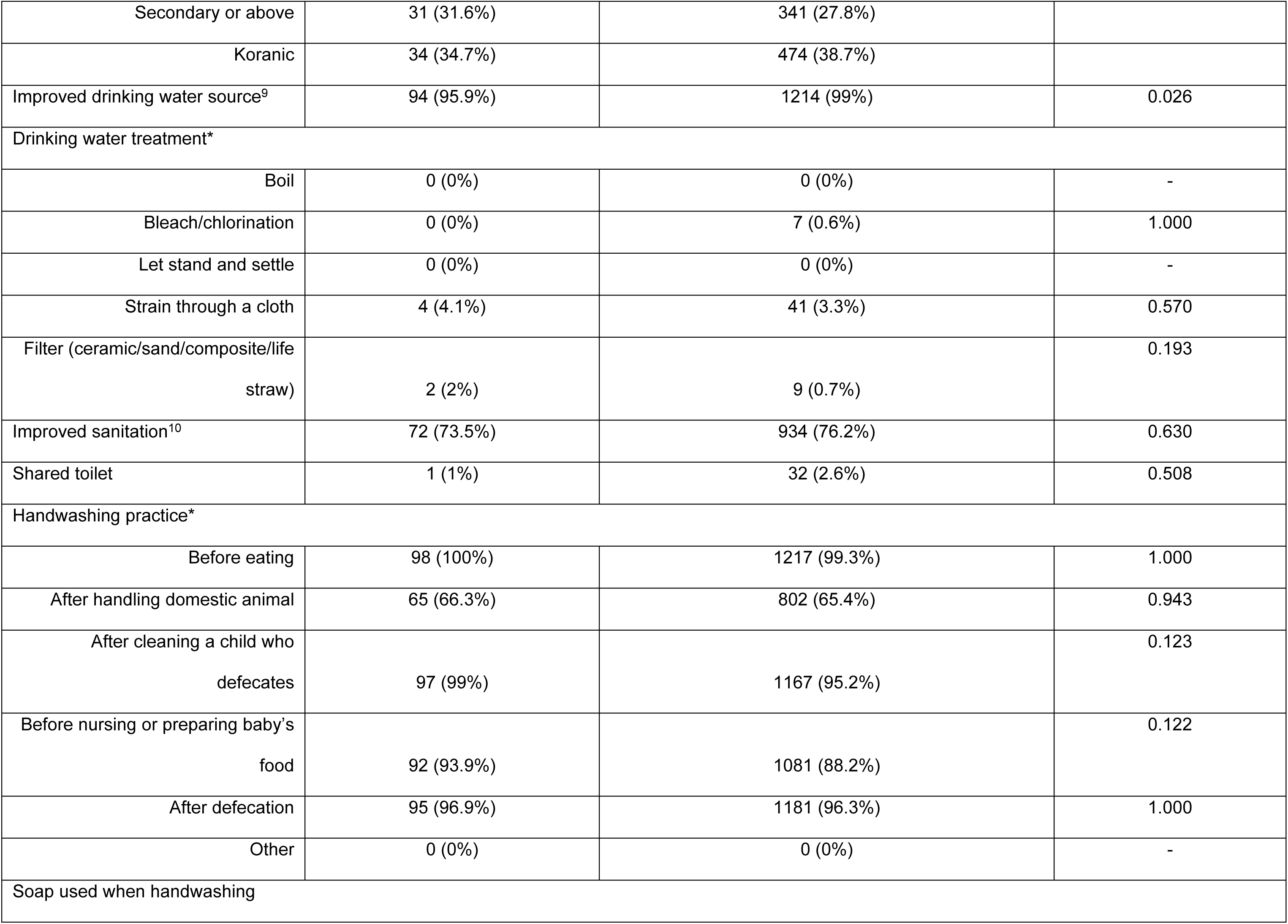
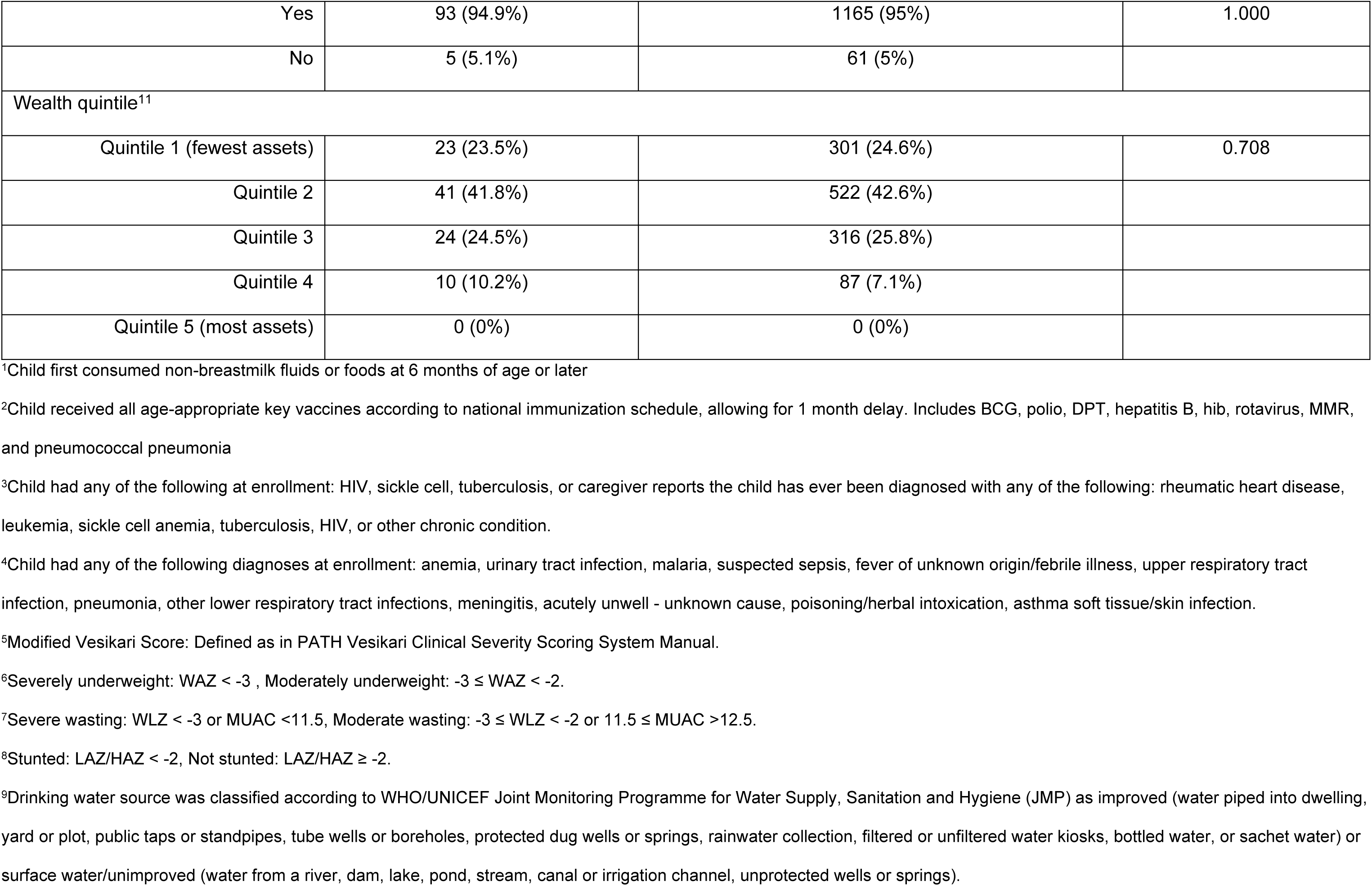
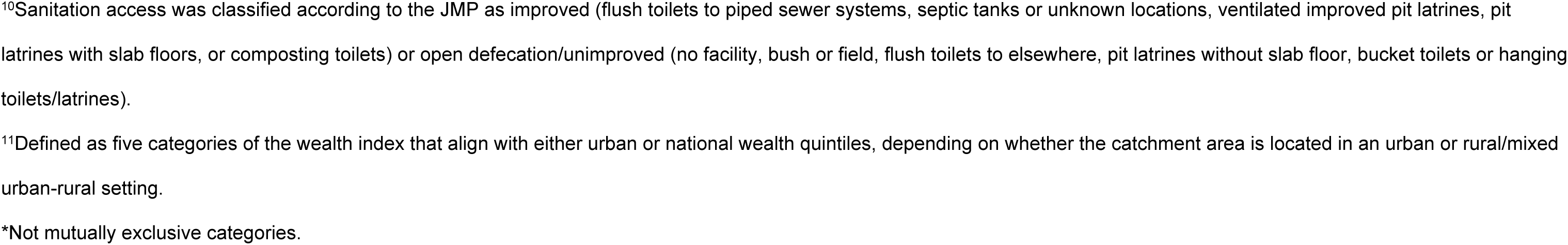
Host, clinical and sociodemographic factors of hospitalization among children aged 6-35 months in The Gambia

## Notes

### Competing Interest Statement

The authors have declared no competing interest.

### Clinical Protocols

https://academic.oup.com/ofid/issue/11/Supplement_1

### Author Declarations

This secondary analysis did not require additional ethical clearance beyond the approvals of the parent study at each country site, details of which can be found elsewhere (cited as reference 27 in the manuscript)

